# SYSTEMS AND NETWORK BIOLOGY ANALYSIS COMBINED WITH MACHINE LEARNING IDENTIFIES KEY IMMUNE RESPONSE PROFILES AND POTENTIAL CORRELATES OF PROTECTION FOR THE M72/AS01E TUBERCULOSIS VACCINE

**DOI:** 10.1101/2025.01.13.25320455

**Authors:** Oluwaseun oluwatosin Taofeek, Solomon Osarumwense Alile, Evans Mauta Elcanah, Louis Odinakaose Ezediuno, Ifeoluwa Adeniyi George, Olawale Moses Oyewole, Peter Ngo’la Owiti, Lateef Adegboyega Sulaimon

**Affiliations:** Department of Chemical Sciences, Crescent University, Abeokuta, Nigeria; Department of Computer Science, Pan-Atlantic University, Ibeju Lekki, Lagos, Nigeria; Department of Biochemistry, Jomo Kenyatta university of Agriculture and Technology, Nairobi, Kenya; Department of Microbiology, University of Benin, Benin, Nigeria; Department of Pure and Applied Zoology, Federal University of Agriculture, Abeokuta, Nigeria; Department of Biochemistry, Federal University of Agriculture, Abeokuta, Nigeria; C/O GAVI Steering Committee for Vaccines and Immunization, Nairobi, Kenya

**Keywords:** Tuberculosis Vaccine, M72/AS01E, Gene Expression Analysis, Network Biology, immune Response

## Abstract

Tuberculosis claims around 1.5 million lives annually. The M72/AS01E vaccine candidate is an innovative effort demonstrating a 50% reduction in the incidence of active TB in adults. However, optimization and effective immunization strategies against TB depends heavily on precise identification of specific molecular signatures active in vaccine protection. In this study, we employed weighted gene co-expression network analysis, machine learning and network biology to investigate the gene expression patterns of peripheral blood mononuclear cells, identifying transcriptomic markers of vaccine protection. Our comprehensive exploration of publicly available gene expression dataset comprising samples from subjects vaccinated twice with 10μg of M72/AS01E vaccine one day post-second dose (D31) and one week post-second dose (D37) in a Phase IIA clinical trial revealed intense induction of multiple gene modules, indicative of acute/immediate immune response at D31 that subsided by D37. Thirty-one hub genes with significant elevation/correlation with immune protection were identified significantly mediating key events in immunity to TB. The more selective profile at D37 involved additional adaptive immunity pathways including Th1/Th2/Th17 differentiation, T cell receptor and cytokine signaling. The functional relevance of these biomarkers in predicting vaccine response was further analyzed using the random forest classifier demonstrating high accuracy in distinguishing between vaccinated and non-vaccinated samples. Additionally, the study pinpointed a miRNAs-transcription factors (TF)-target regulatory network excavating key TF, miRNA, mRNAs mediating vaccine protection. Our results provided new insights into immune protection of M72/AS01E vaccine meriting further study aiming to advance its optimization and informing development of future TB vaccines.

## Introduction

Tuberculosis (TB) reclaimed its position as the number one infectious disease killer worldwide in 2023, overtaking COVID-19 and causing an estimated 10.8 million cases and 1.25 million deaths [1]. Historically, vaccines have played crucial roles in combating infectious diseases saving millions of lives annually by preventing the onset of diseases. It is emphasized that vaccination programs in populations with high (TB) incidence could lower disease rates by 1.6-fold or more [2]. Until now, the Bacillus Calmette-Guérin (BCG) TB vaccine, introduced in 1921 remains the only licensed TB vaccine which already has been administered over 4 billion times worldwide with varying efficacy against pulmonary TB in adolescents and adults [2,3]. Development of effective vaccines therefore for TB is a critical focus in the global fight against TB particularly in low-and-middle-income countries where the disease burden is highest [3,4]. The recent decade has witnessed increased research interest into TB vaccines covering novel candidates including live attenuated, killed mycobacterial, subunit, viral vector, and mRNA vaccines with 16 vaccine candidates currently in various stages of clinical trials [5]. Among these, promising candidates have included M72/AS01E and VPM1002 showing potential for improved immune responses and efficacy against TB [3,5].

The M72/AS01E vaccine candidate is an innovative immunization effort that combines the M72 antigen with the AS01E adjuvant to enhance the immune response against Mycobacterium tuberculosis [6]. It was reported to demonstrate a 50% reduction in the incidence of active TB disease in adults with latent TB with acceptable safety, reactogenicity, and immunogenicity profiles [7–9]. While this is encouraging, the World Health Organization (WHO) has highlighted critical knowledge gaps regarding the vaccine’s mechanism of protection and implores basic research to better understand the immune correlates involved [6]. Although previous studies have characterized the vaccine’s ability to elicit broad immune responses via the CD4+ T-cell dynamics and antibody production, supported by activation of Th1/Th17 pathways and cytokine release [2,7], significant gap exists in understanding the detailed molecular networks orchestrating these effects. To address these gaps, this study integrated advanced systems biology tools and machine learning techniques to perform multi-layered analysis of a public RNAseq data from a Phase IIA clinical trial of the M72/AS01E vaccine to identify key genes involved in the immune response to Mycobacterium tuberculosis (M. tb) vaccination and their modular coexpression interactions underlying immune response kinetics. We further employed multiple bioinformatics resources to predict the miRNAs and transcription factors (TFs) mediating regulation of identified hubs genes critical for host immune response to vaccination against M. tb infection. The miRNAs are small non-coding RNA molecules (18–22 nucleotides) that regulate the expression of approximately one-third of human genes, including those involved in innate and adaptive immune responses, by executing post-transcriptional gene regulation. [9,10]. The TFs on the other hand play a pivotal role in activation or repression of the transcription rate at the pre-transcriptional stage [11]. Integrating these analyses, we constructed a miRNA-target-TF regulatory network, identifying hub drivers of vaccine-induced immunity. These findings provide a more potential granular understanding of the molecular underpinnings of M72/AS01E-induced protection, offering valuable insights for optimizing current vaccine strategies and guiding the development of next-generation tuberculosis vaccines.

## Materials and Methods

## Gene Expression Dataset

The gene expression omnibus (GEO) [https://www.ncbi.nlm.nih.gov/geo/], an open source of high-throughput gene expression and functional genomics data (accessed on August 3, 2024) was utilized to extract RNAseq data (accession number is GSE102574 generated on Illumina HiSeq 2000 platform) [12] recently deposited (December 30, 2023) following a Phase IIA clinical trial (NCT01669096) of the GSK M72/AS01E adjuvant vaccine. Briefly, this trial project involved sample collection from 18 according-to-protocol subjects that were vaccinated twice with 10μg M72/AS01E. Responses were measured at three time points in peripheral blood mononuclear cells with the majority of measurements being made early after the 2nd dose of the vaccine. RNA samples were collected from all eighteen subjects at pre-vaccination (D0), one day after second vaccination (D31), and one week after vaccination (D37) time points making a total 54 samples analyzed.

## Identification of differentially expressed genes

To understand the expression pattern of host genes between different time points upon vaccination, we employed the Bioconductor package, DESeq2 in RStudio (version 4.4.1) [(13)] to identify the differentially expressed genes (DEGs) at D0 vs D31, D0 vs D37, and D31 vs D37. Before performing differential expression analysis, genes with low counts across all samples were filtered out, retaining only those genes that had at least 15 counts in the smallest group. The significant DEGs were filtered based on a threshold of adjusted p-value < 0.05, absolute log2 fold change > 1.5, and Benjamini & Hochberg (False discovery rate) <0.05. The DEGs were visualized using volcano plots. The volcano plot assessed the relationship between log2 fold changes and adjusted p-values, highlighting genes with large fold changes and significant p-values.

## Construction of Gene Co-Expression Network

RNA-seq count data and metadata were retrieved for the GSE102574 dataset, and counts were filtered to exclude genes with expression counts <20 in more than 75% of the dataset. The resulting data was normalized using the variance stabilizing transformation (VST) followed by quality control using hierarchical clustering, principal component analysis (PCA), Mahalanobis, and Euclidean distance calculations to detect and remove outlier samples. A signed co-expression network was constructed using Weighted gene co-expression network analysis (WGCNA) [(14)] via the WGCNA package of the Comprehensive R Archive Network (CRAN). A soft-thresholding optimal power of 14 was selected based on scale-free topology. Modules were then identified using the blockwiseModules function with dynamic tree cutting (maximum block size: 16,000; merge cut height: 0.25). After determining the gene modules using dynamic cutting, the eigenvector value of each module was calculated, and modules were merged based on similarity into new, consolidated modules. These modules were then summarized by eigengenes, and their correlations with timepoint traits (D0, D31, and D37) were analyzed using Pearson correlation. Associations or correlations were visualized in heatmaps. Traits were binarized, and categorical levels were transformed into binary matrices for enhanced module-trait analysis. Module sizes and eigengene-trait correlations were archived while gene contents were exported into Cytoscape where their interaction network was visualized and Cytoscape’s MCODE [15] plugin was utilized for clustering the network to generate the hub genes mediating vaccine response. By leveraging large-scale gene expression data to uncover co-expression networks and hub genes, WGCNA provided a robust framework for identifying key molecular players and biomarkers involved in immunological responses to vaccination.

## Enrichment and Pathway Analysis

We performed downstream analysis, including functional annotation and pathway enrichment analysis to provide biological interpretation of the roles of the hub genes in host immnune response to vaccine. The analysis of canonical pathways and function associations was performed using Enrichr analysis tool [16] on differentially expressed hub genes. Canonical pathway analysis in Enrichr utilized combined score metric to rank enriched terms, balancing p-value significance and Z-score of gene set overlaps. The z-score algorithm predict pathway activation or inhibition and function analysis associated DEGs with known biological functions and diseases. Visualization of enrichment results was facilitated via bubble plot combined with Sankey diagram demonstrating statistically significant pathways (adjusted p-value < 0.05), with annotations derived from the KEGG datatbase. These analyses provided insights into biological pathways and functions enriched in the DEG dataset, enhancing the understanding of their roles in the studied context.

## Machine learning

To test the predictive potential of identified modules and genes, validating their functional relevance in the context of the M72/AS01E vaccine response, we applied a machine learning approach to analyze the gene expression data. Using the MLSeq R package [17]. The dataset was split into training (70%) and testing (30%) subsets using stratified sampling to maintain the distribution of timepoints across both sets. To address the scale of gene expression data, normalization was performed using DESeq2’s VST, which minimizes the bias in data. For feature selection, the top 500 variable genes were selected based on variance across the samples and used for subsequent analysis. Several classifiers were trained and evaluated on the filtered data to predict the timepoint labels. For the Random Forest (RF) model, the number of trees was set to 500, and the number of features considered at each split was set to the square root of the total number of features. Model performance was evaluated using the accuracy metric derived from the confusion matrix on the test set. In addition to the RF model, other classifiers—Support Vector Machine (SVM), K-Nearest Neighbors (KNN), and the voom-based Linear Discriminant Analysis (LDA) were trained and evaluated using similar cross-validation techniques. Performance metrics such as accuracy, sensitivity, and specificity were computed from confusion matrices for each model. For the best-performing classifiers, feature importance scores were calculated, and the top 10% of ranking genes were extracted. The biological relevance of these genes was verified using the Enrichr gene set enrichment analysis tool [16]. The classifier that demonstrated the most biologically relevant results, as determined by the functional annotation of its top-ranking genes, was selected as the best-performing model. Genes identified from this model were highlighted for their high predictive potential and strong correlation with vaccine response.

## Identification of miRNAs targeting differentially expressed genes

The R package multiMiR [18] was employed to retrieve the potential miRNAs targeting the DEGs. It comprises a wide collection of validated and predicted miRNA–target interactions and also details their associations with drugs and diseases. The multiMiR package is also composed of murine and human datasets from 14 external databases—eight predicted, three validated, and three drug- or disease-related miRNA–target databases. In this study, we only considered three databases (miRecords, miRTarBase, and TarBase) for analysis of the predicted target miRNAs of the DEGs [19].

## Construction of miRNA-mRNA network

In this study, we focused on validated miRNAs interactions with the DEGs and interaction scores were calculated and filtered for moderate interaction scores (≥4). The resulting data were structured into a node attribute table, distinguishing between miRNAs and DEGs, and prepared for visualization using the igraph package [20] to construct a miRNA-target interaction network with directed edges representing the interactions. Node attributes were assigned based on type (miRNA or DEG), and labels were set to match node names. The miRNA-target interaction network was visualized using Cytoscape version (3.10.2) [15] and their functional relevance investigated using the DIANA-miRPath v4.0 tool [21]

## Identification of transcription factors (TFs) regulating DEGs

The iRegulon (version 1.3) computation tool [22] was utilized in Cytoscape to identify TFs in the gene sets of DEGs. iRegulon is a database containing approximately 10,000 TF motifs used to detect enriched TF motifs in the regulatory regions of each gene. Each candidate TF is linked to enriched TF motifs, which are then used to identify the subset of corresponding direct target genes. The tool integrates multiple motif sources, including TRANSFAfC, JASPAR, SwissRegulon, HOMER, and ENCODE, to robustly link TF motifs to their corresponding TFs and direct target genes. By incorporating ChIP-seq profiles, particularly from the ENCODE project, iRegulon enhances the identification of key regulatory networks, providing a comprehensive view of motif-to-TF associations and regulatory mechanisms.

## Construction of a miRNA-Target-TF regulatory network

TF-target (DEGs) interaction networks with a Normalized Enrichment Score (NES) greater than 9 were selected for downstream analysis. The miRNA-Target-TF regulatory network was then constructed for visualization in Cytoscape. To understand the hub miRNA-Target-TF regulatory network central to immune responses upon vaccination, the MCODE Cytoscape plugin [15] was employed to identify highly interconnected clusters (modules) within the network. These modules represent key regulatory interactions that may play crucial roles in modulating immune responses. The miRNA-Target-TF regulatory network, enriched with information from DEGs, miRNAs, and TFs, provides a comprehensive view of the potential regulatory mechanisms underlying the immune responses to the M72/AS01E vaccine.

## Results

### Transcriptional landscape of peripheral blood mononuclear cells in response to the M72/AS01E tuberculosis vaccine

We analyzed RNA-seq data from PBMCs collected from 18 according-to-protocol subjects vaccinated twice with 10μg M72/AS01E tuberculosis vaccine to elucidate the influence of the vaccine on the transcriptional landscape of PBMCs, providing insights into its immunological effects. Our initial exploration via PCA of the rlog-transformed counts to assess the overall variability in the dataset demonstrated a clear group separation (Fig. 1A). The samples (D0, D31, D37), represented by red, green, and blue points, respectively, appear to cluster relatively well by their group labels along the principal components (PC1 and PC2). PC1 and PC2 accounted for 41.64% and 11.95% of the variance, respectively. This suggests that the primary variance captured in the data is related to the experimental conditions (time points), not external batch effects. Volcano plots in Fig. 1B-D captured the differentially expressed genes between D0 and D31, D0 and D37, and D31 and D37 respectively highlighting the significant changes in gene expression. A total of 468 DEGs (73 upregulated and 395 downregulated) were identified between D0 and D31 (Fig. 1B, Table S1). Between D0 and D37, 166 downregulated DEGs were identified (Fig. 1C, Table S2), while 818 DEGs (304 upregulated and 514 downregulated) (Fig. 1D) were identified between D31 and D37 (Table S3).

**Fig. 1:**
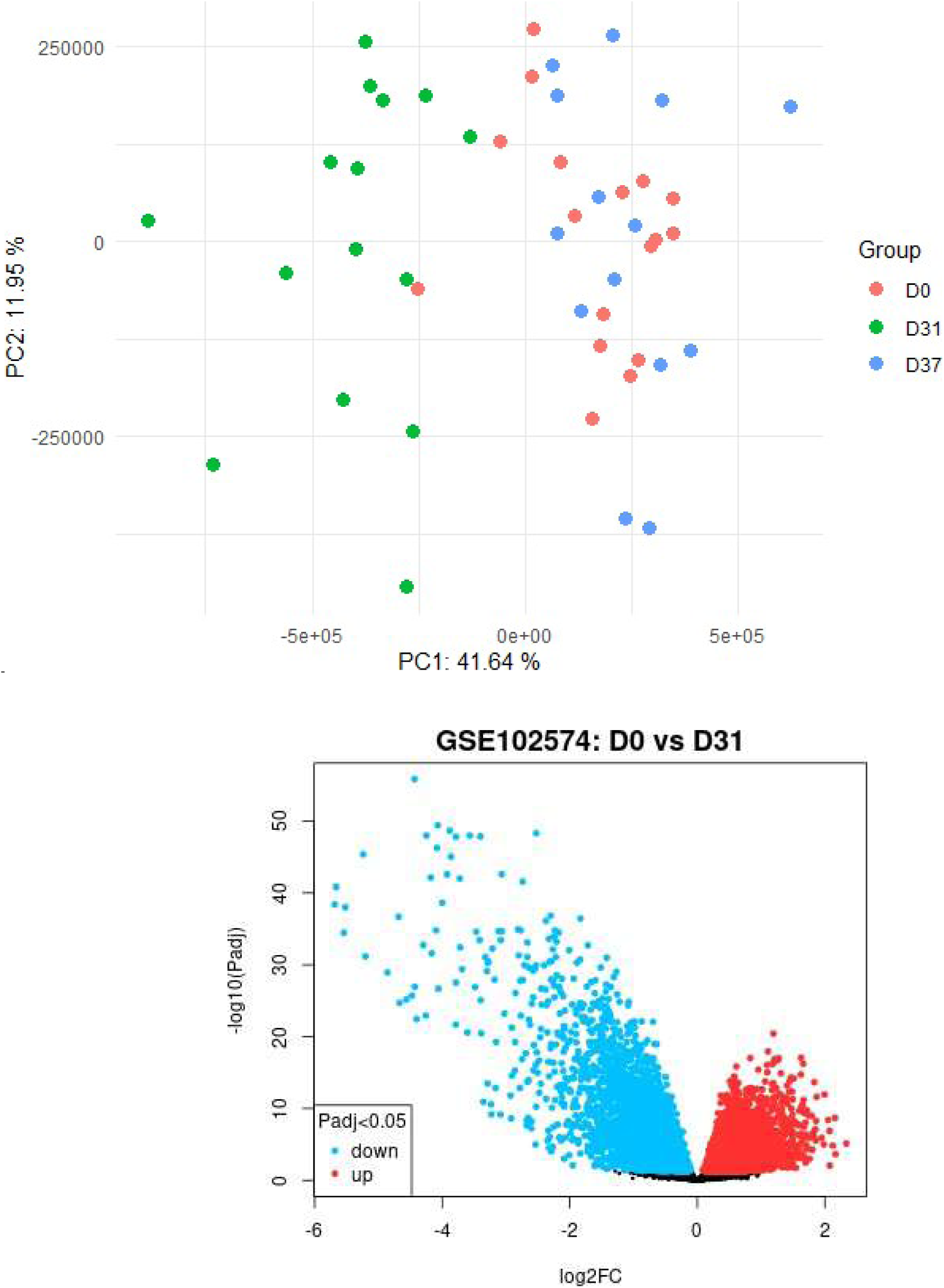
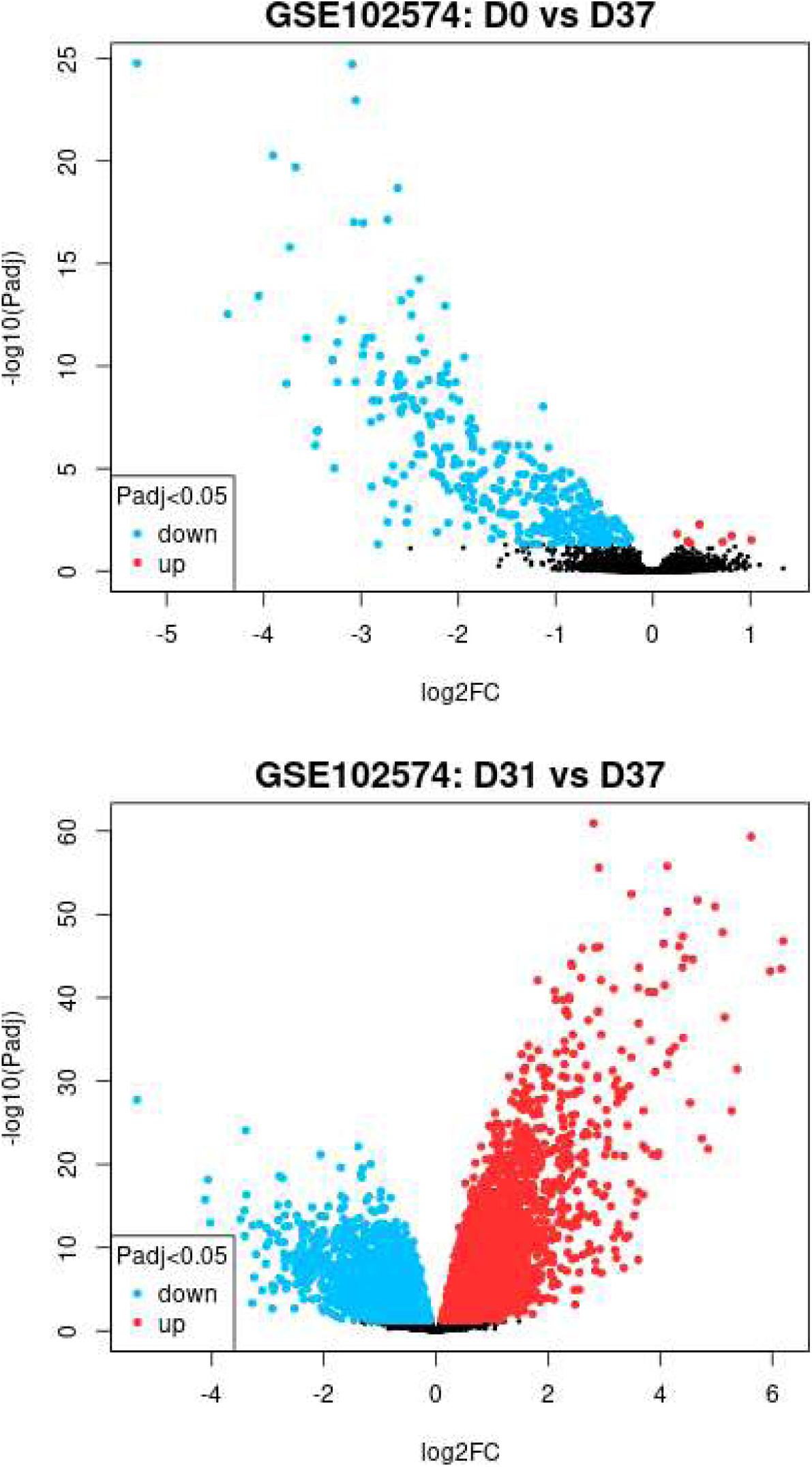
Transcriptional landscape of PBMCs of vaccinated and non-vaccinated subjects. (A) Principal component analysis (PCA) of rlog-transformed counts showing clear separation of timepoints: prevaccination, D0 (red), one day post second dose, D31 (green), and one week post second dose, D37 (blue). (B) Volcano plot representing differentially expressed genes between D0 and D31 (C) Volcano plot representing differentially expressed genes between D0 and D37 (D) Volcano plot representing differentially expressed genes between D31 and D337.

### WGCA to identify key biomarkers of Host response to vaccination

To identify key potential biomarker targets of host immune response to vaccination, we employed a weighted gene co-expression network analysis (WGCNA). Clustering analysis was conducted using the average linkage method and Pearson correlation to assess relationships and group similarities among all samples. A weighted gene co-expression network was constructed using a soft-thresholding power of 14 to achieve scale-free topology (Fig. 2A). Co-expression modules were identified using the blockwiseModules function with a signed topological overlap matrix and a merge cut height of 0.25 to merge similar modules. The resulting module eigengenes, which summarize the expression profiles of each module, were computed and exported. Relationships between modules and genes were visualized using a dendogram with color annotations for unmerged and merged modules (Fig. 2B). The number of genes in each module was also summarized and recorded for downstream analysis. Our examination revealed that M72/AS01E vaccine induced broad, intense initial expression of multiple gene modules, indicative of an acute/immediate immune response at D31 that subsided by D37 (Fig. 2C). We performed intramodular analysis within the WGCNA framework to identify driver genes that are highly correlated with immune response to vaccination. Among the various modules identified and analyzed, the blue (2655 genes, Corr. 0.89, p < 0.001 at D31, Corr. -0.42, p < 0.01 at D37), green (1032 genes, Corr. -0.78, p < 0.001 at D31, Corr. 0.44, p < 0.01 at D37), and red (275 genes, Corr. -0.36, p < 0.05 at D31, Corr. 0.74, p < 0.001 at D37) modules were selected for detailed examination due to their strong association with vaccination response at D31 and D37 (Fig. 2C). Statistical correlation of genes belonging to these modules to vaccine response at different timepoints were first calculated by applying the Student’s t-distribution and the significance of the correlations for each gene was ranked using the resulting p-values (p < 0.05). Using the StringApp and MCODE plugins in Cytoscape [15], we identified the driver hub genes of vaccine response which included downregulated CXCL10, DDX58, DHX58, GBP1, GBP2, GBP4, GBP5, IFI16, IFIH1, IRF1, IRF7, IRF9, OAS1, OAS2, OAS3, STAT1, and STAT2 in the blue module (MYB) (Fig. 2D.). Hub genes identified in the green module (MYG) included CCL5, CD244, CD27, CTLA4, CXCR6, FASLG, GZMB, IL2RB, KLRD1, LCK, NCR3, PRF1, TBX21, ZAP70 (Fig. 2E). For the red module, identified hub genes included CDCA3, CDCA7, CDK1, CENPE, CENPF, CHEK1, DEPDC1B, FEN1, MCM2, NCAPH, NUF2, ORC1, PCLAF, PRC1, TYMS, UHRF1, ZWINT (Fig. 2F) These hub genes obtained suggests that these modules contain genes potentially pivotal to the immune response to vaccination, highlighting their promise as candidates for monitoring vaccine efficacy or predicting response.

**Fig. 2:**
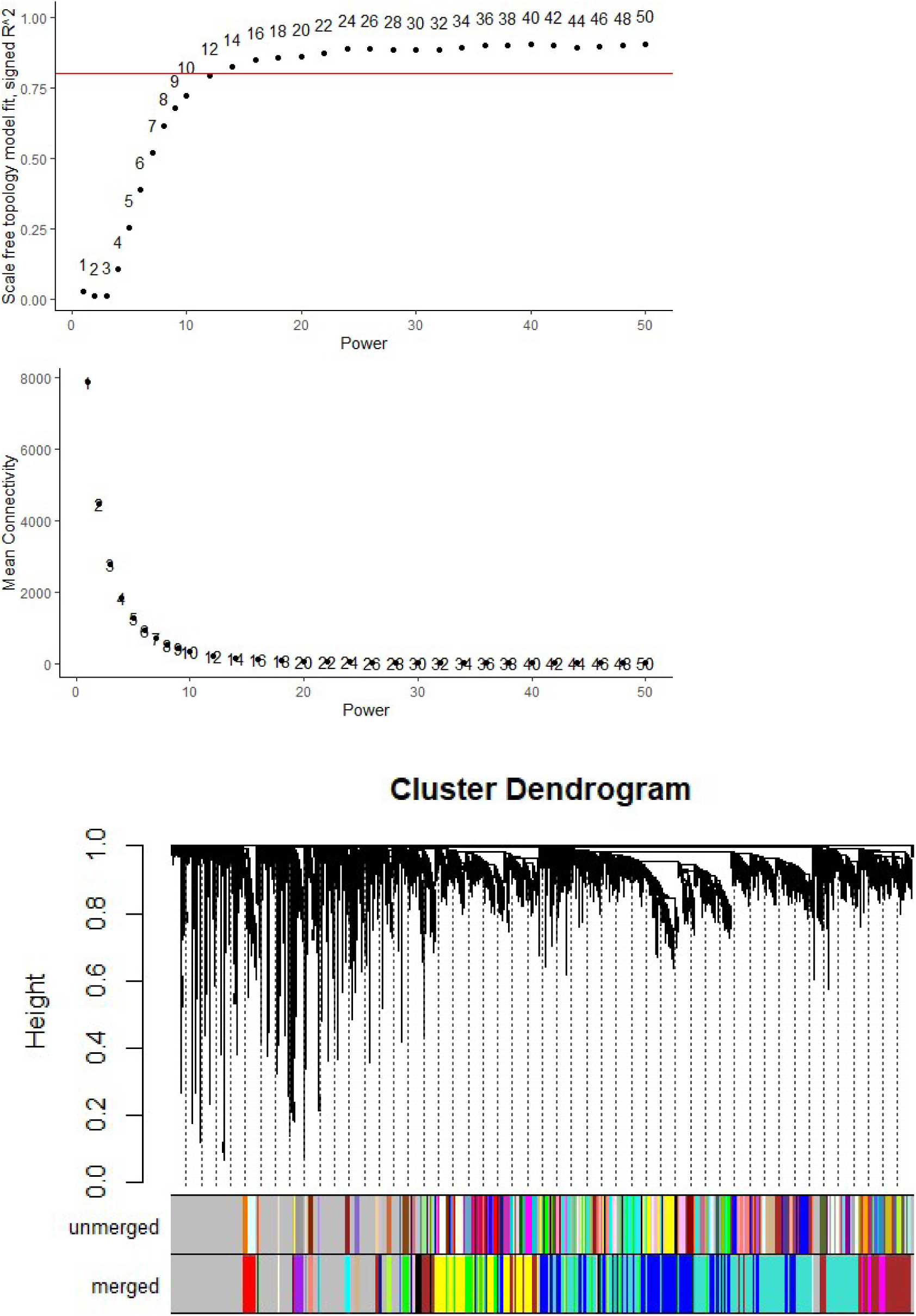
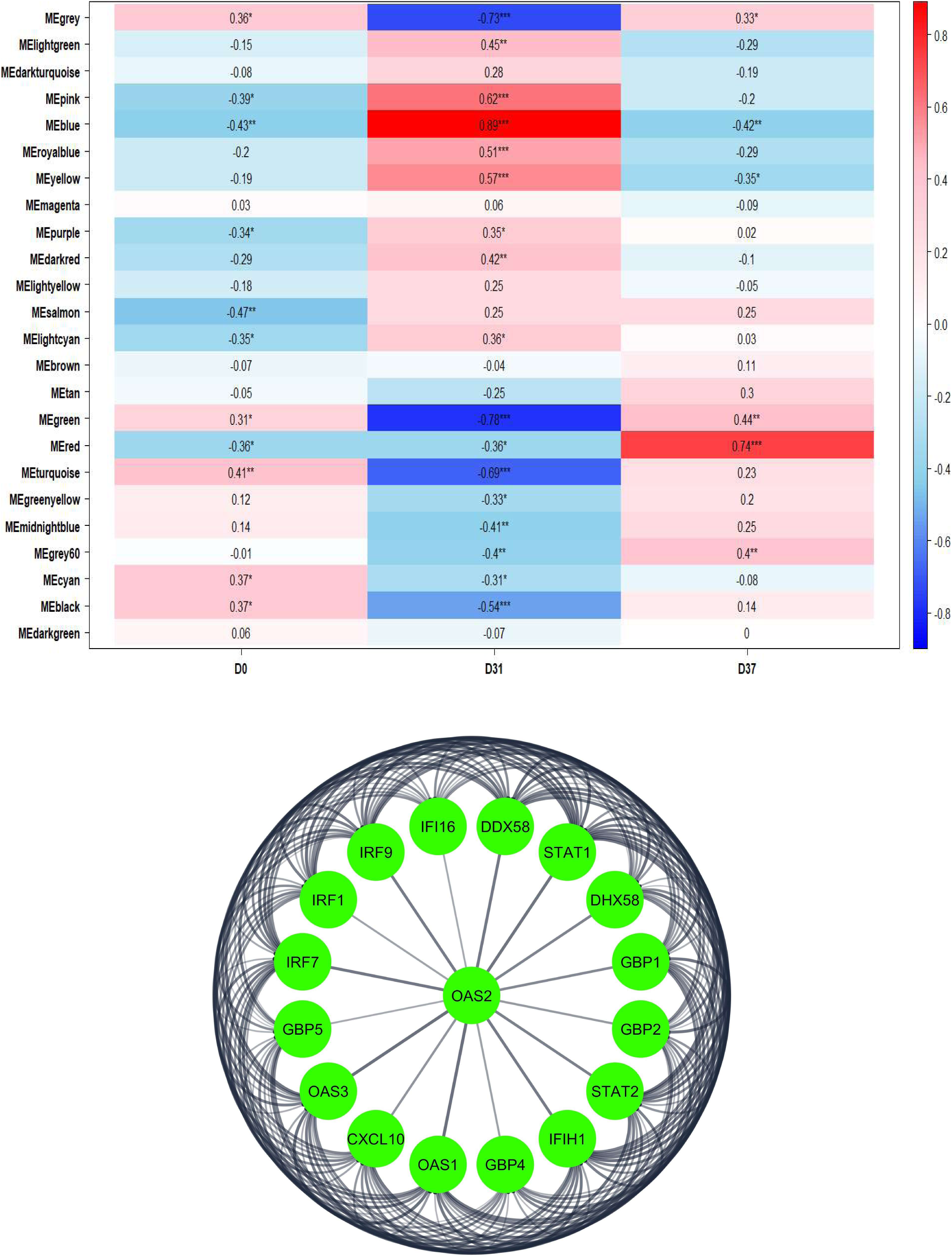
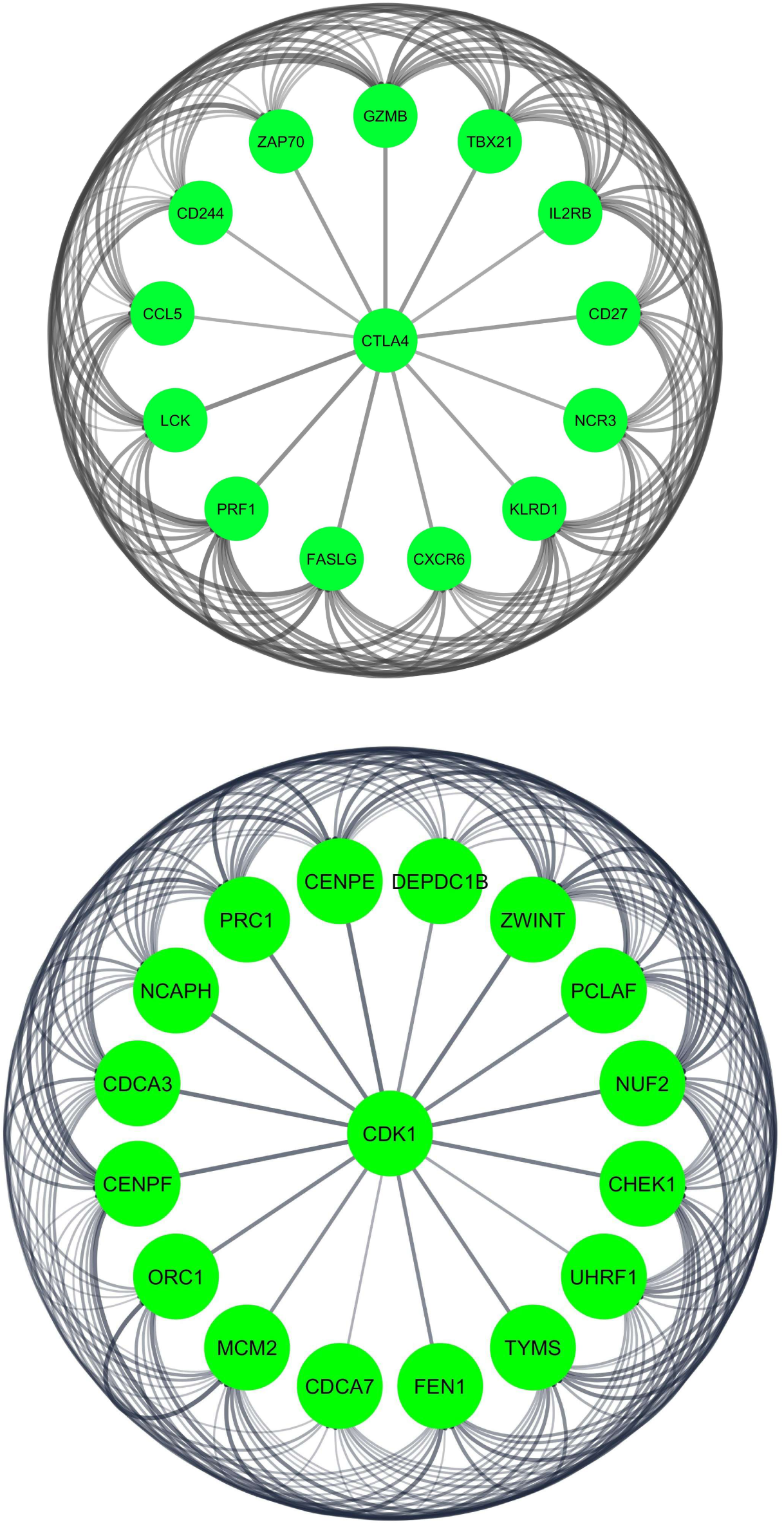
Identification of key biomarkers of Host response to vaccination using WGCA (A) Network construction with a soft-thresholding power of 14. (B) Clustering analysis using the average linkage method and Pearson correlation to assess relationships and group similarities among all samples. (C) Heatmap representing the correlation of vaccine response with different modules. (D) Hub genes mediating vaccine response identified in the blue module (E) Hub genes mediating vaccine response identified in the green module (F) identified hub genes in the red module.

### Functional enrichment and pathway analysis

Hub genes identified in MYB exhibited a high correlation with vaccine response and significant changes in expression impacting key pathways such as NOD-like receptor, RIG-I-like receptor, Toll-like receptor, Chemokine, TNF, and the JAK-STAT signaling pathways, key events in immunity to TB (Fig. 3A). The more selective profile at D37 involved additional adaptive immunity pathways such as Th1, Th2, and Th17 differentiation, T cell receptor signaling, and cytokine interactions mediated by identified hub genes in the MYG (Fig. 3B). Enrichments obtained for hub genes of the red module significantly impacted cell cycle, DNA replication and p53 signaling pathways (Fig. 3C). Significant correlations of pathways impacted by genes in MYB and MYG suggest that the hub genes drive a significant response to vaccination and are therefore prioritized for subsequent analyses.

**Fig. 3:**
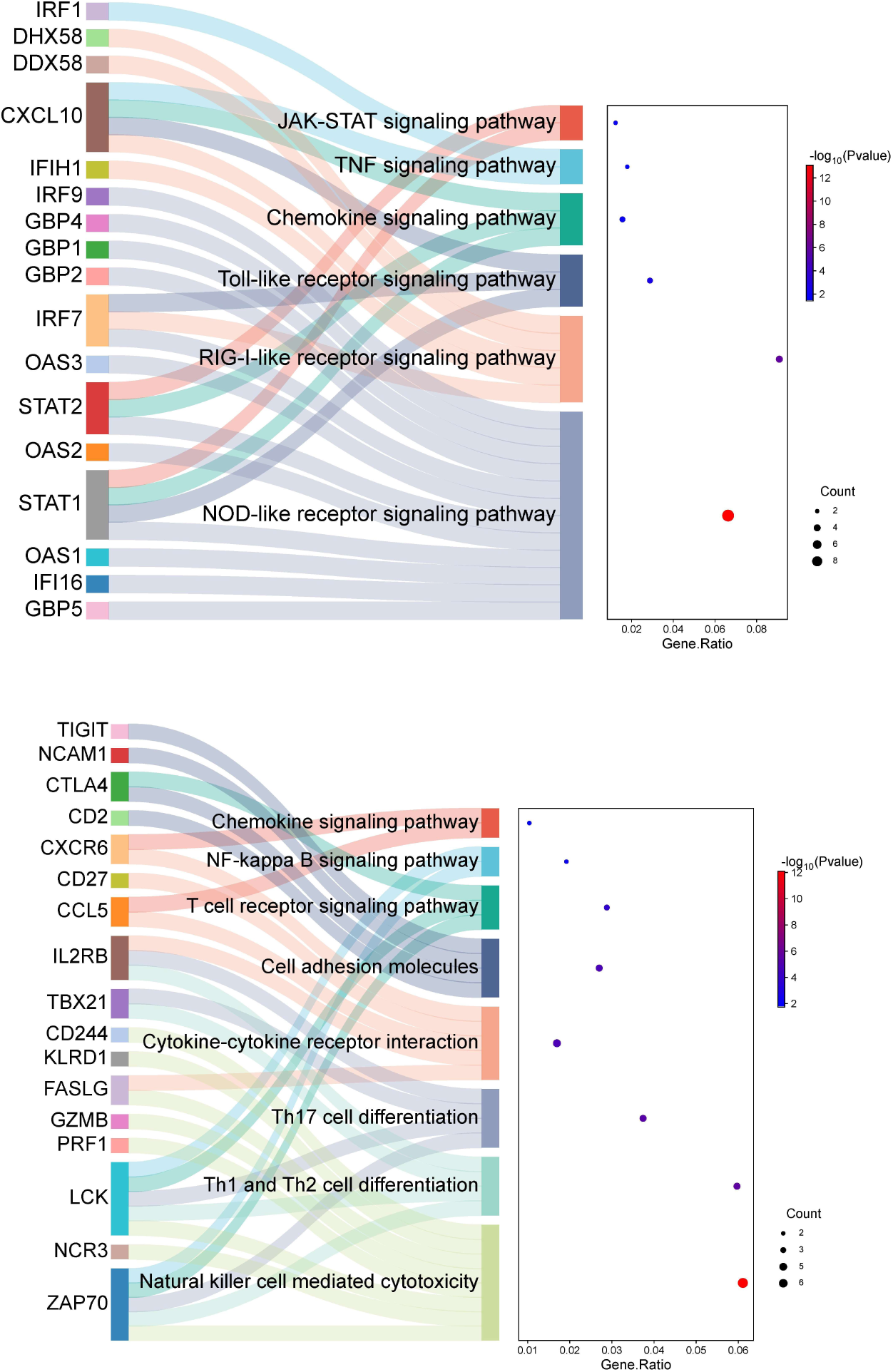
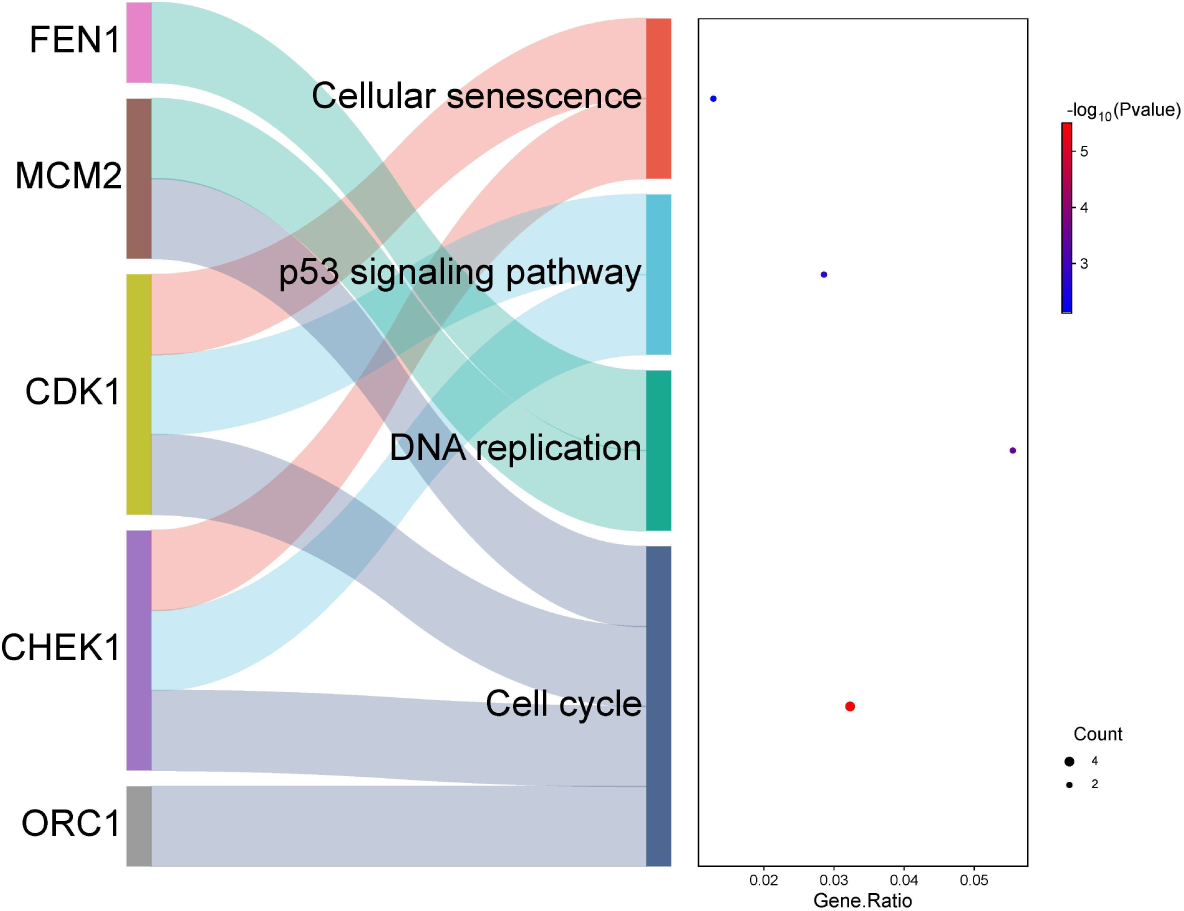
Pathway and functional analysis of identified hub genes in PBMCs from M72/AS01E tuberculosis vaccine recipients. (A) Top enriched canonical pathways identified for hub genes in the blue module. (B). Enriched pathways identified for hub genes in the green module (C). Significant correlations of pathways impacted by hub genes in the red module.

### Machine leaning approach to identify predictive potential of identified hub genes

To test the vaccine response predictive potential of the identified hub genes and their functional relevance in the context of the M72/AS01E vaccine response, we applied MLSeq framework with various machine learning classifiers including RF, SVM, KNN, and the voom-based LDA method. Among these classifiers, the RF model emerged as the best performer, achieving an overall accuracy of 90.91% (95% CI), as summarized in Table 1. It demonstrated robust sensitivity across multiple classes, achieving perfect specificity (100%) for classes D31 and D37 and perfect sensitivity (100%) for both classes D0 and D31. However, class D37 exhibited a slightly reduced sensitivity of 66.67%, likely due to inherent variability in the response profiles or data sparsity for that class. This result was achieved after hyperparameter optimization using a grid search approach with 10-fold cross-validation repeated 10 times. Notably, the enrichment factor (EF) for class D37 was the highest at 9.57, suggesting the RF model’s superior ability to prioritize true positives at the top of the ranked list. In contrast, the performance of other classifiers was suboptimal. The SVM model, for instance, showed poor sensitivity for D31 and D37 (0%), despite achieving 100% specificity for these classes. The KNN classifier exhibited moderate performance with an overall accuracy of 72.73%, but its sensitivity for class D37 was markedly lower (33.33%). The LDA model performed comparably to RF in terms of accuracy, sensitivity, and specificity. However, it achieved lower enrichment factor (EF) values, with the highest being 3.667 for class D31. A confusion matrix was plotted (Figure 4A) for the RF classifier to evaluate the false positive and false negative predictions across all classes. It showed minimal false predictions, aligning with its high precision and recall metrics. Additionally, the receiver operating characteristic (ROC) curves and the corresponding area under the curve (AUC) values further substantiated the RF classifier’s robust predictive performance, with AUC values consistently above 0.95 across all classes (Table 1; Fig. 4B). The RF model’s overall performance demonstrates its utility in classifying vaccine response at these timepoints. This model consistently ranked GBP1, GBP2, STAT1, IFIH1, CXCL10, IRF1, GBP5, OAS3, IRF7, OAS1, STAT2, GBP4, identified in MYB within the top 0.54% of the entire dataset based on importance scores, demonstrating their significance in vaccine response. Enrichment analysis of these genes showed their role in key immune processes such as signaling by IFN-γ, IFN-α, IFN-β, IL-20, IL-9, IL-21, IL-27, IL-6, IL-35, IL-6, IL-2, IL-10, IL-12, and chemokine receptors (Fig. 4C). These metrics underscore the classifier’s robustness in distinguishing vaccinated responders from non-responders, highlighting its potential for advancing vaccine efficacy evaluation and immunological research.

**Fig. 4:**
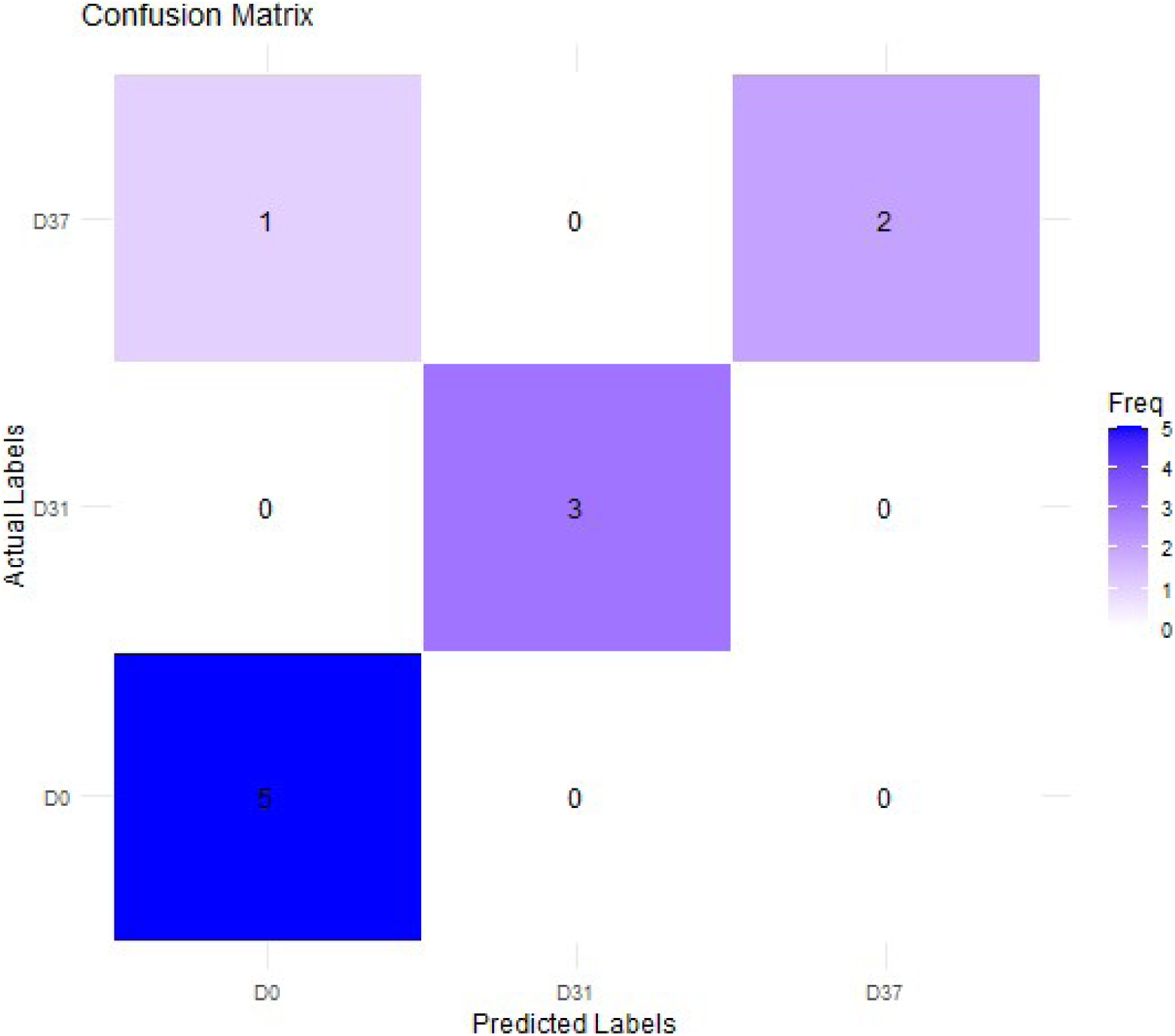
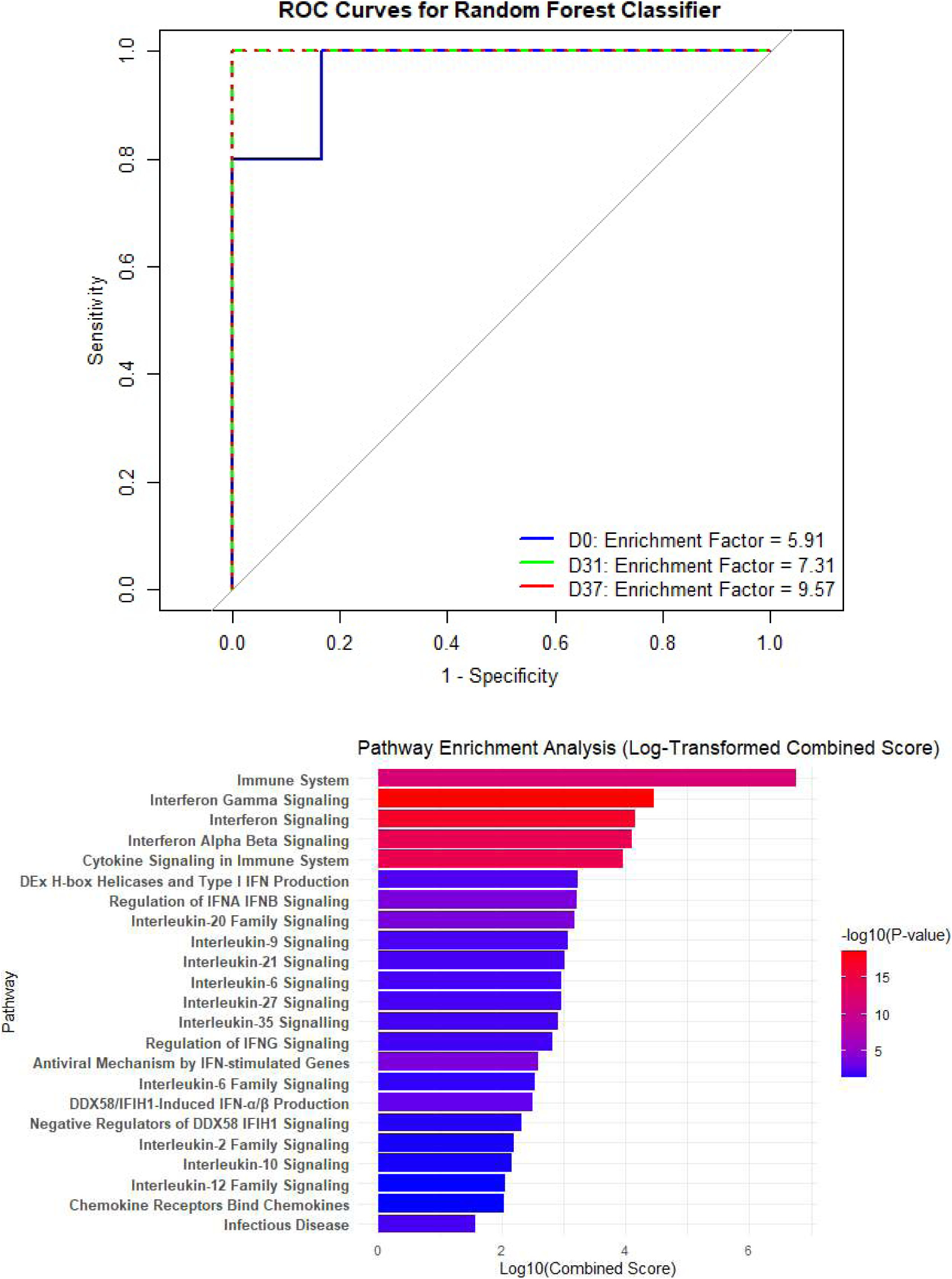
Machine learning approach to identify biomarkers with predictive potential for vaccine response. (A) Confusion matrix evaluating false positive and negative predictions of the RF classifier. (B) ROC curves for the RF classifier. (C) Functional annotation of top predictive genes identified by RF classifier.

**Table 1:**
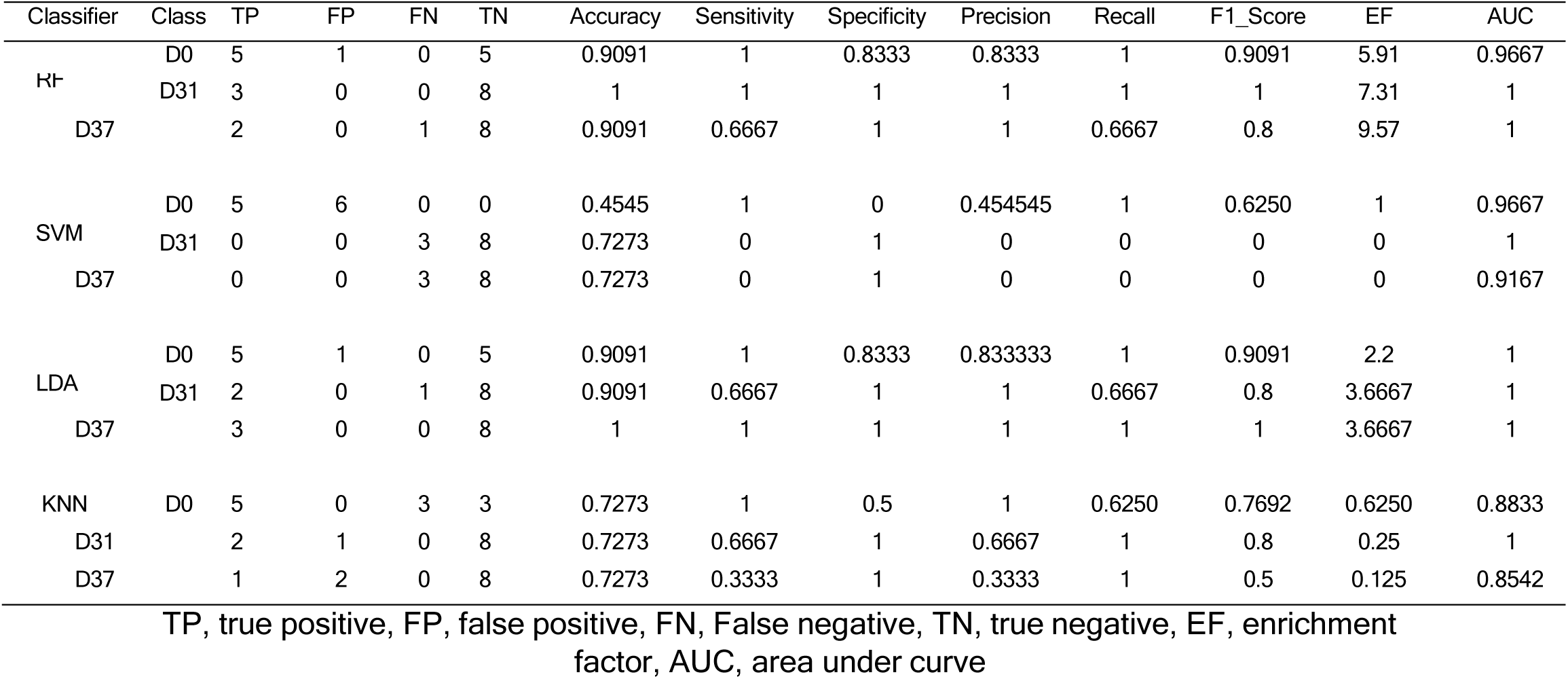
Performance metrics comparison across models.

### Identification of miRNAs-TFs-target regulatory network driving vaccination response

The multiMiR package identified 36 unique validated miRNA interactions for 11 downregulated hub genes in MYB and 28 miRNAs for 8 hub genes in the MYG based on miRecords, miRTarBase, and TarBase validated interaction scores (Tables 2 and 3). A threshold interaction score of 4 was applied to prioritize the most significant miRNAs. The miRNA-mRNA networks are shown in Fig. 5A and Fig. 5B. The functional analysis of these miRNAs reveals these miRNAs target pathways involved in cell cycle, autophagy, FoxO signaling, bacterial invasion, p53, MAPK, AMPK, HIF-1, TGF-beta, mTOR, TNF, chemokine signaling, apoptosis, Fc gamma R-mediated phagocytosis, T cell receptor signaling, and necroptosis (Fig. 5C and Fig. 5D). Through databases including TRANSFAC, FactorBook, SwissRegulon, HOMER, and JASPAR in the iRegulion tool [22], we have identified 19 TFs of the hub genes of MYB based on NES score > 9 and 58 for those of the MYG (Tables 4 and 5). To identify the interactions of the transcriptional factors and regulatory elements of miRNA, we constructed the miRNAs-TFs-target regulatory network using Cytoscape (Fig. 5E and Fig. 5F). For MYB, the network was characterized by 63 nodes and 906 edges while that of MYG was characterized by 94 nodes and 721 edges. To understand the core regulatory interaction, we generated two subnetworks each for the regulatory networks. For MYB, the first subnetwork involved the interaction of TFs including PURA, PRDM1, and SPI1 with miRNAs such as hsa-let-7i-5p, hsa-miR-34a-5p, hsa-miR-125a-5p, hsa-miR-146a-5p, hsa-miR-454-3p, and hsa-miR-130a-3p (Fig.5G). The second subnetwork showed the interaction of OAS2 and OAS3 genes with TF including E2F1, ZEB1, MYB, IRF3, IRF4, IRF5, IRF2, IRF6, IRF7, IRF8, IRF9, and ZNF683 (Fig. 5H). For MYG, the first subnetwork involved interaction of ELF1 and YY1 TFs with hsa-miR-6504-3p, hsa-miR-4438, hsa-miR-6506-5p, hsa-miR-7151-3p, hsa-miR-5089-5p, hsa-miR-98-5p, hsa-miR-5095, and hsa-miR-619-5p while the second subnetwork involved interaction of SOX15, SOX21, SOX18, SRY, SOX2, and SOX14 with hsa-miR-130b-3p hsa-miR-130a-3p (Fig. 5I and 5J).

**Fig. 5:**
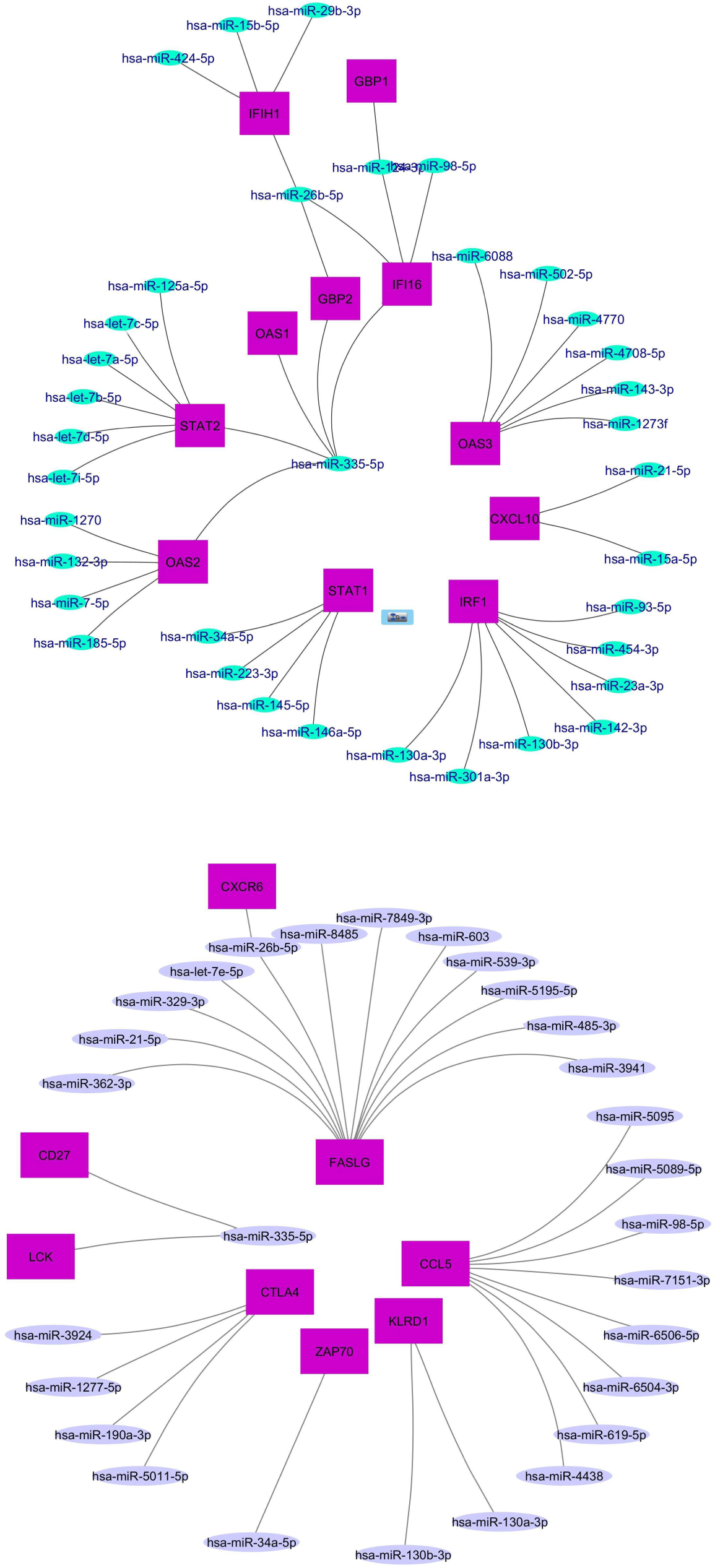
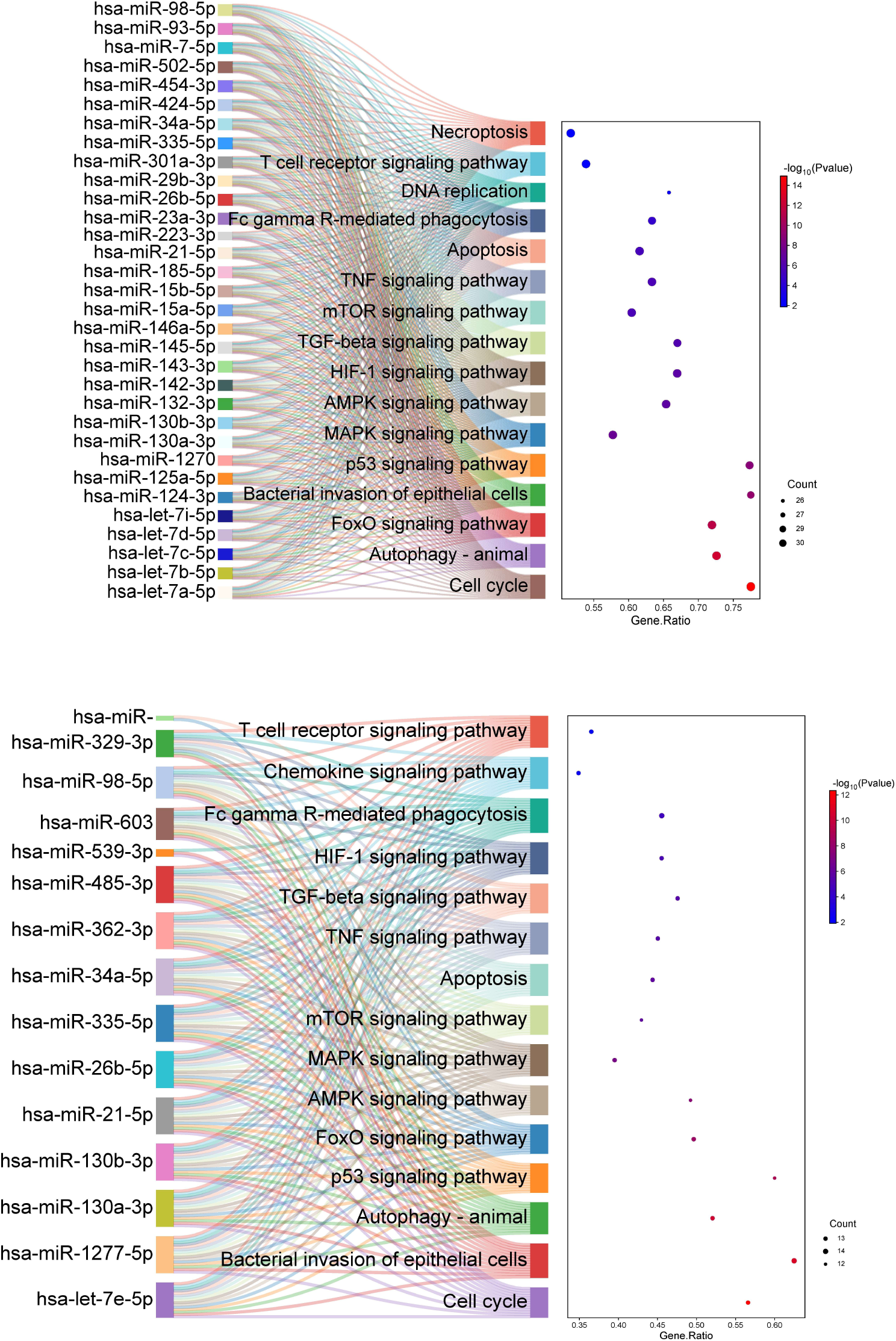
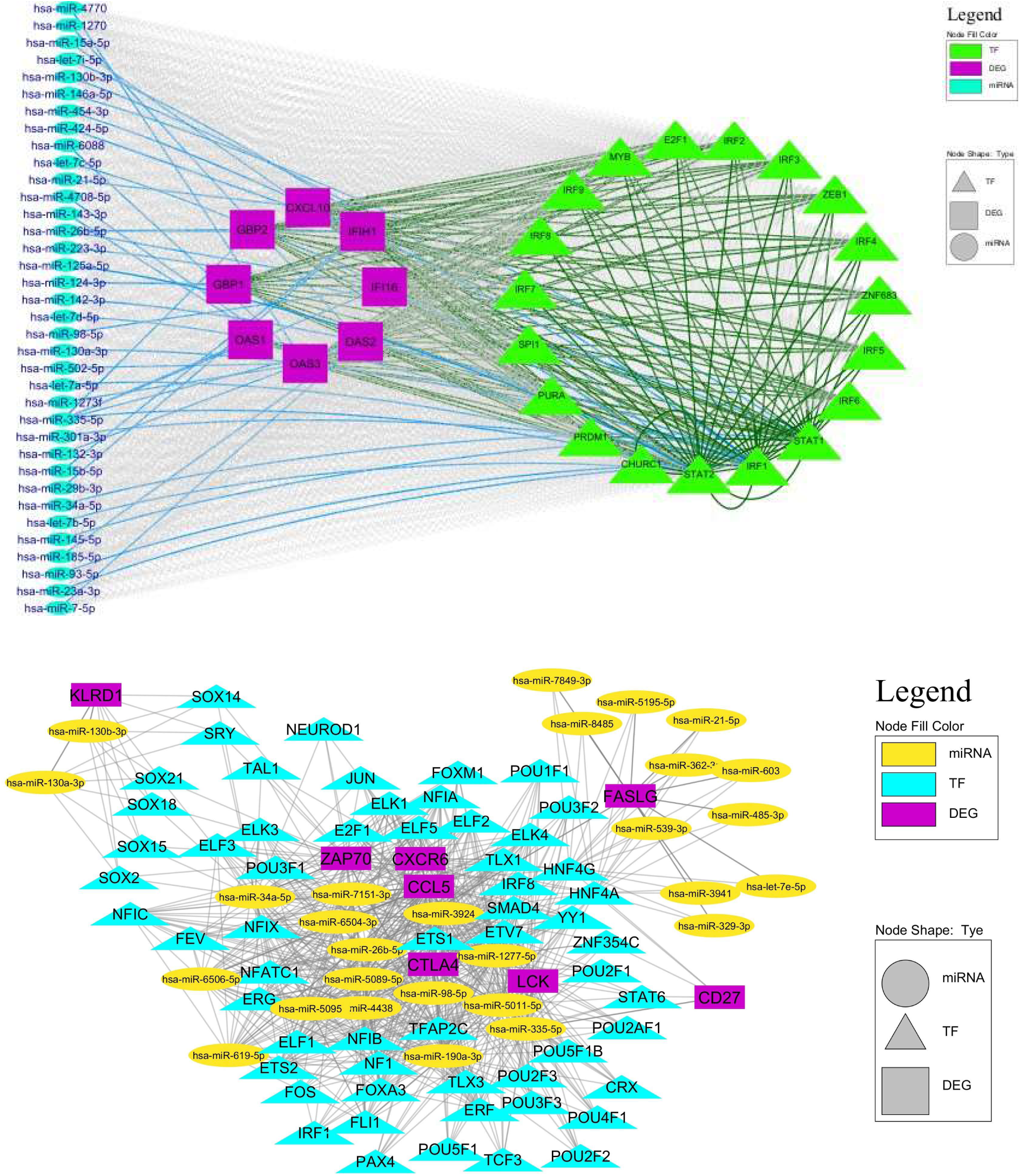
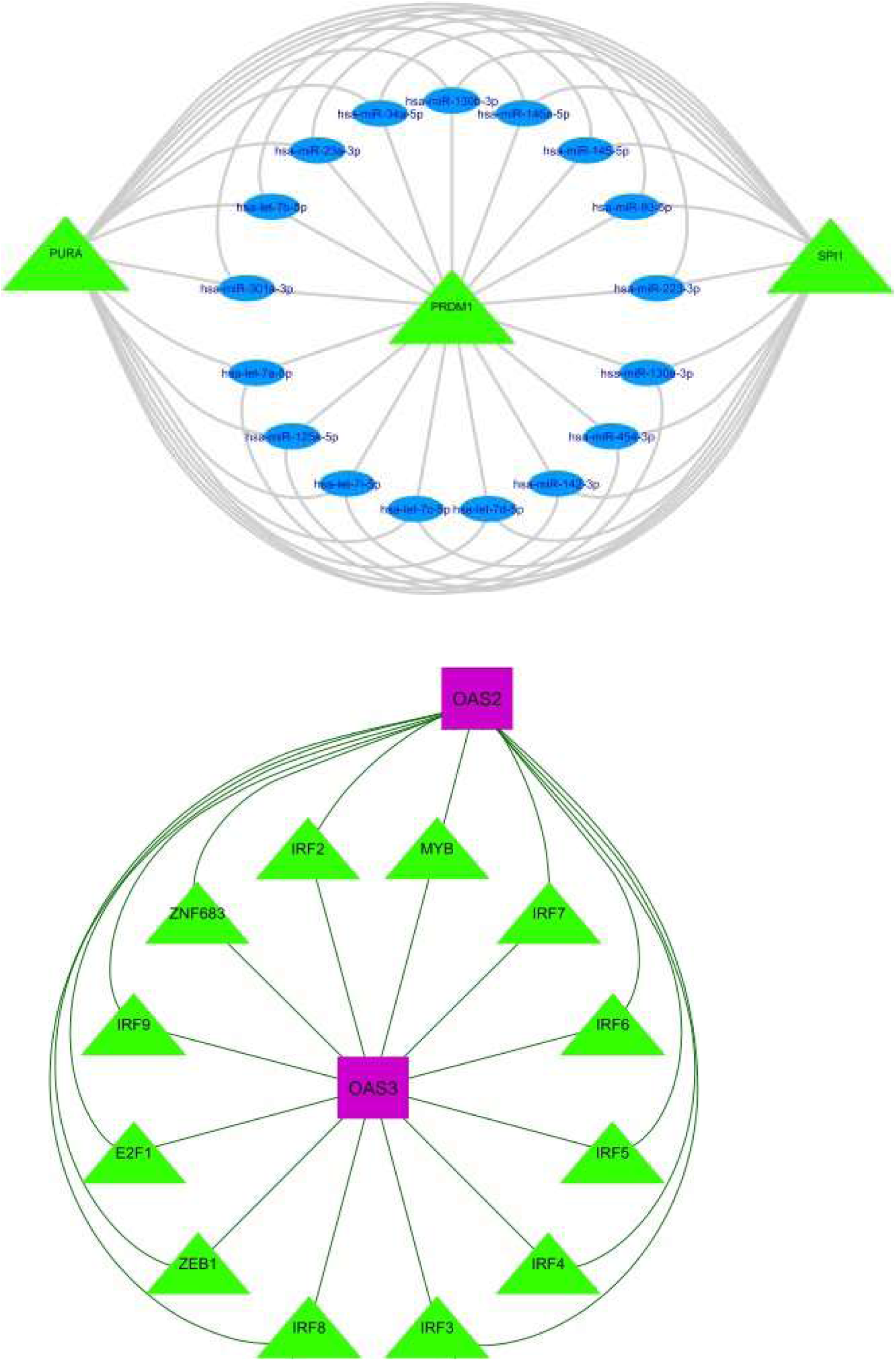
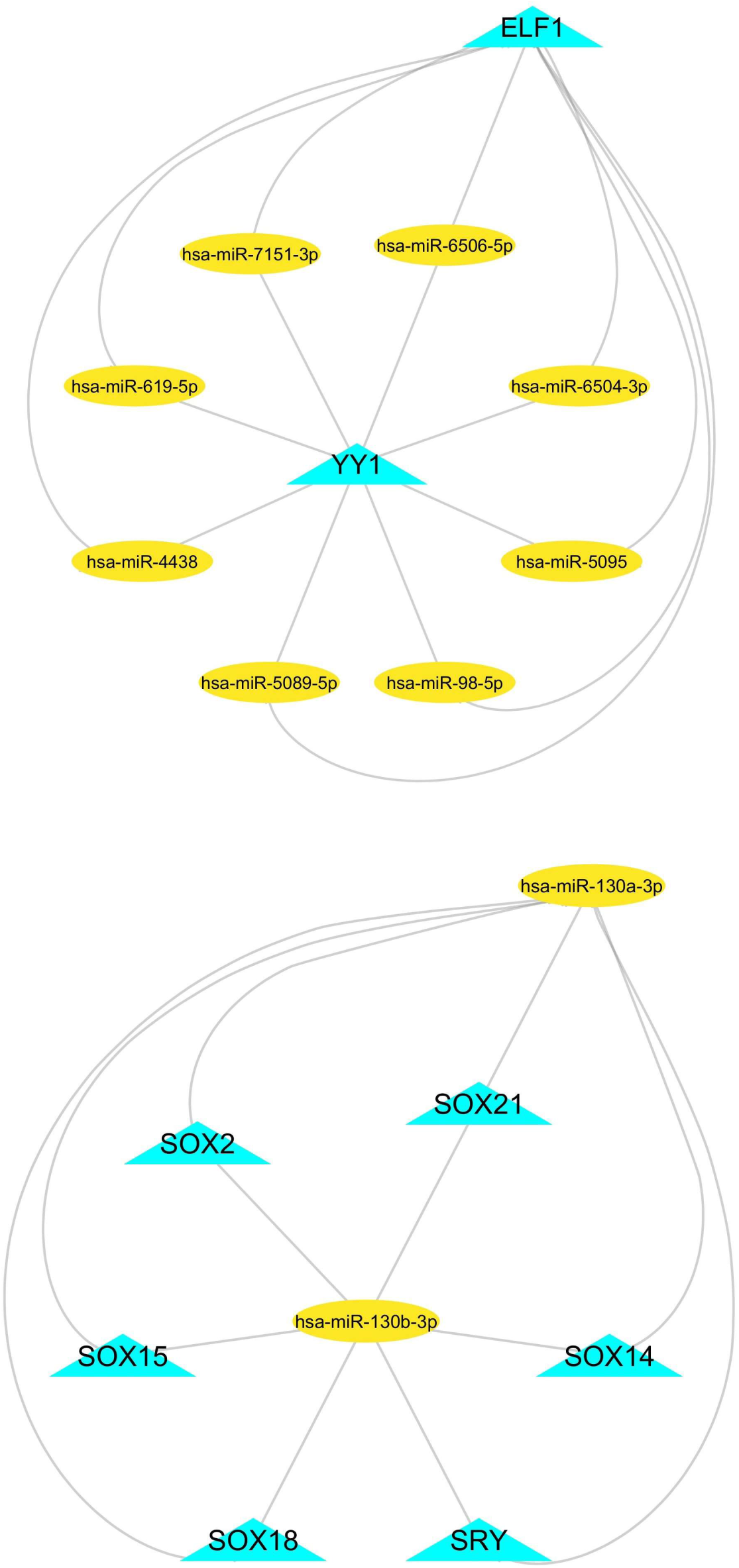
miRNAs-TFs-target regulatory network driving vaccination response (A) miRNA-target network of miRNAs targeting hub genes of the blue module. (B) miRNA-target network of the miRNAs regulating hub genes of the green module (C) pathway enrichment profile of the miRNAs identified to target hub genes of the blue module (D) pathway enrichment profile of the miRNAs identified to target hub genes of the green module (E) miRNAs-TFs-target regulatory network of the blue module (F) miRNAs-TFs-target regulatory network of the green module (G) First subnetwrok of the miRNAs-TFs-target regulatory network of the blue of the blue module (H) Second subnetwrok of the miRNAs-TFs-target regulatory network of the blue module (I) First subnetwrok of the miRNAs-TFs-target regulatory network of the green module (J) Second subnetwrok of the miRNAs-TFs-target regulatory network of the green module.

**Table 2:**
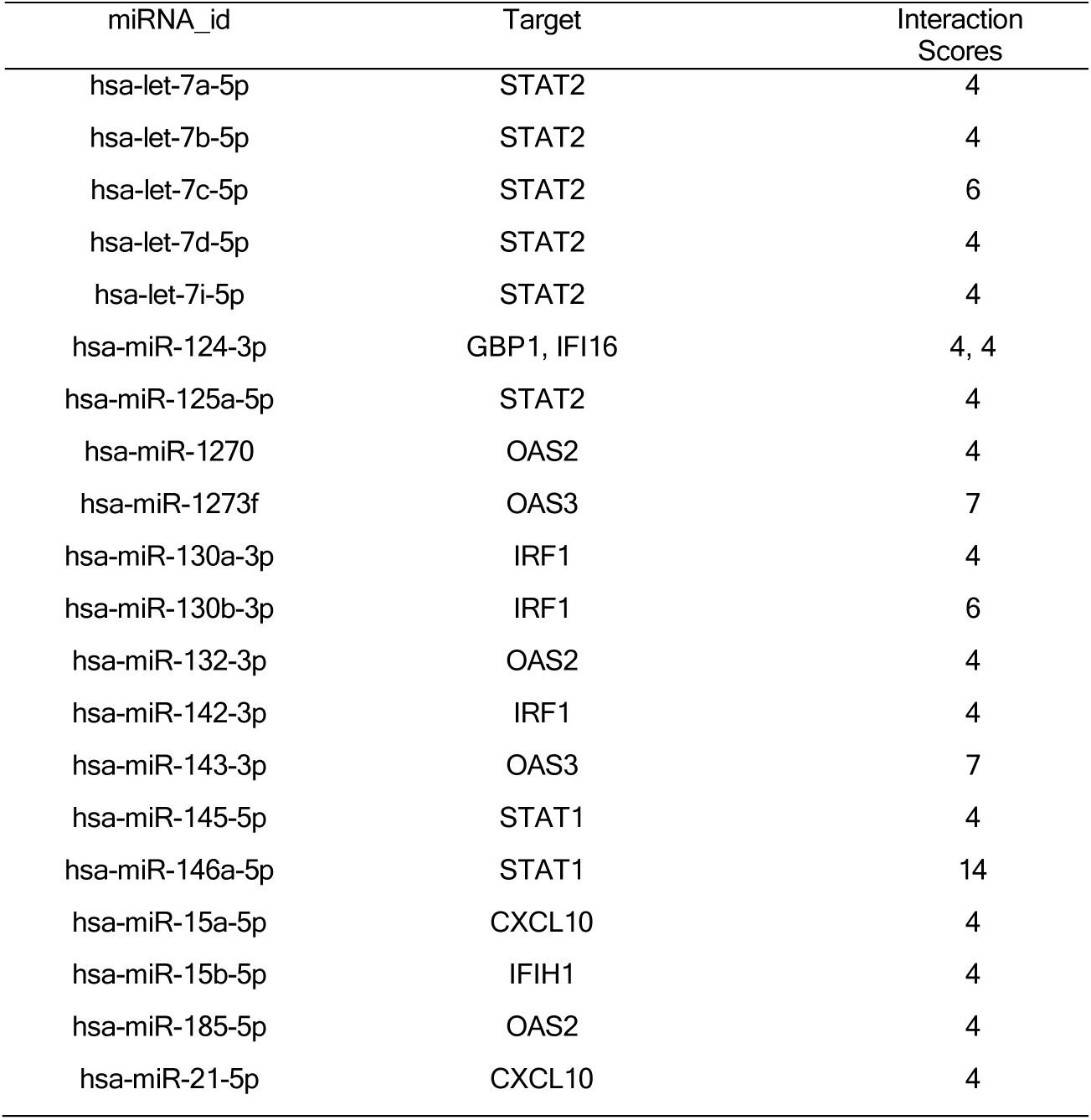
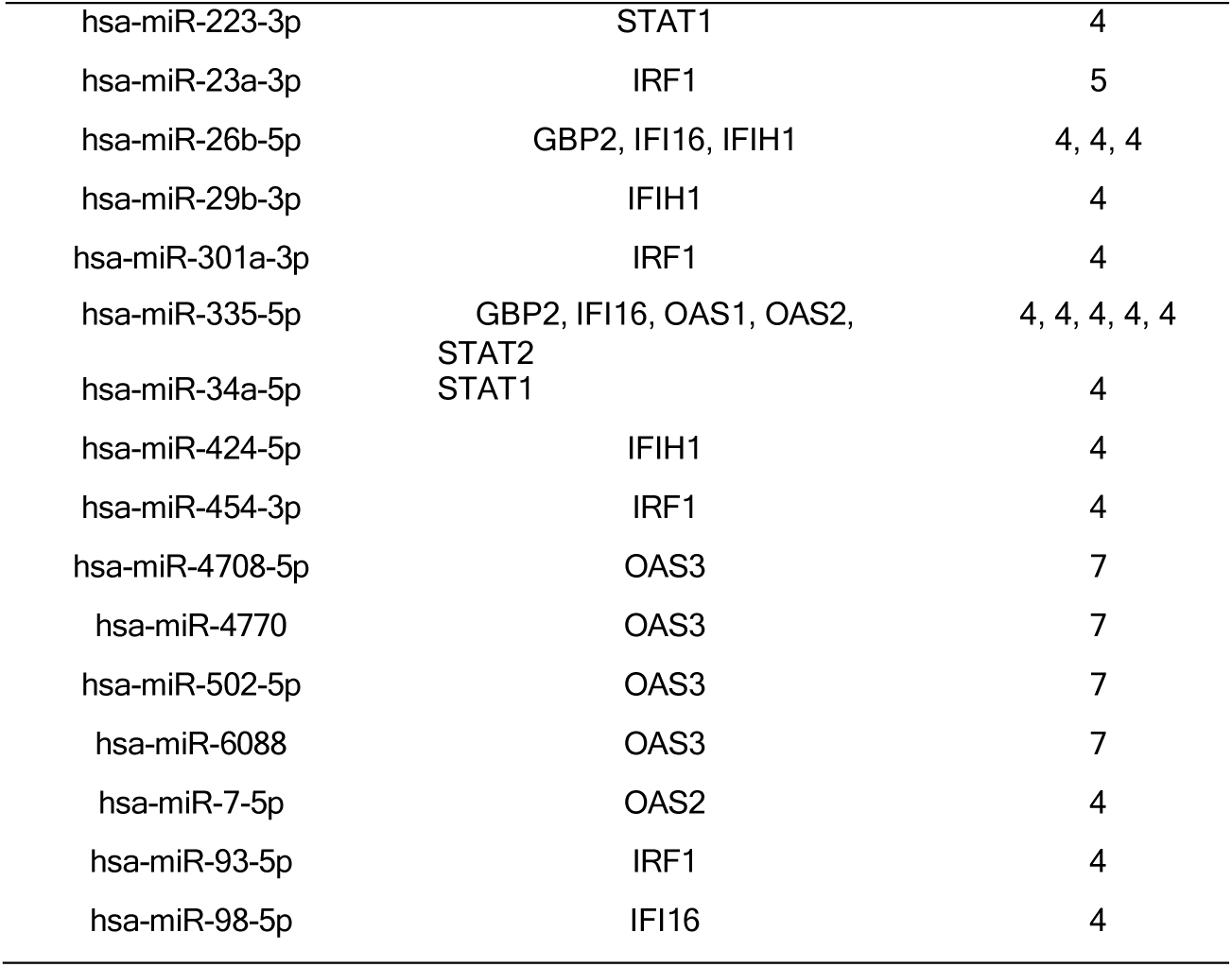
The top miRNAs identified for the hub genes of the blue module based mirecords, mirtarbase and tarbase validated interaction scores.

**Table 3:** The top miRNAs identified for the hub genes in the green module based mirecords, mirtarbase and tarbase validated interaction scores.

| mature_mirna_id | DEGs | Interaction_Scores |
| --- | --- | --- |
| hsa-let-7e-5p | FASLG | 4 |
| hsa-miR-1277-5p | CTLA4 | 9 |
| hsa-miR-130a-3p | KLRD1 | 5 |
| hsa-miR-130b-3p | KLRD1 | 5 |
| hsa-miR-190a-3p | CTLA4 | 9 |
| hsa-miR-21-5p | FASLG | 15 |
| hsa-miR-26b-5p | CXCR6, FASLG | 5, 5 |
| hsa-miR-329-3p | FASLG | 4 |
| hsa-miR-335-5p | CD27, LCK | 5, 5 |
| hsa-miR-34a-5p | ZAP70 | 5 |
| hsa-miR-362-3p | FASLG | 4 |
| hsa-miR-3924 | CTLA4 | 11 |
| hsa-miR-3941 | FASLG | 4 |
| hsa-miR-4438 | CCL5 | 5 |
| hsa-miR-485-3p | FASLG | 4 |
| hsa-miR-5011-5p | CTLA4 | 9 |
| hsa-miR-5089-5p | CCL5 | 5 |
| hsa-miR-5095 | CCL5 | 5 |
| hsa-miR-5195-5p | FASLG | 4 |
| hsa-miR-539-3p | FASLG | 4 |
| hsa-miR-603 | FASLG | 4 |
| hsa-miR-619-5p | CCL5 | 5 |
| hsa-miR-6504-3p | CCL5 | 5 |
| hsa-miR-6506-5p | CCL5 | 5 |
| hsa-miR-7151-3p | CCL5 | 5 |
| hsa-miR-7849-3p | FASLG | 4 |
| hsa-miR-8485 | FASLG | 4 |
| hsa-miR-98-5p | CCL5 | 5 |

**Table 4:**
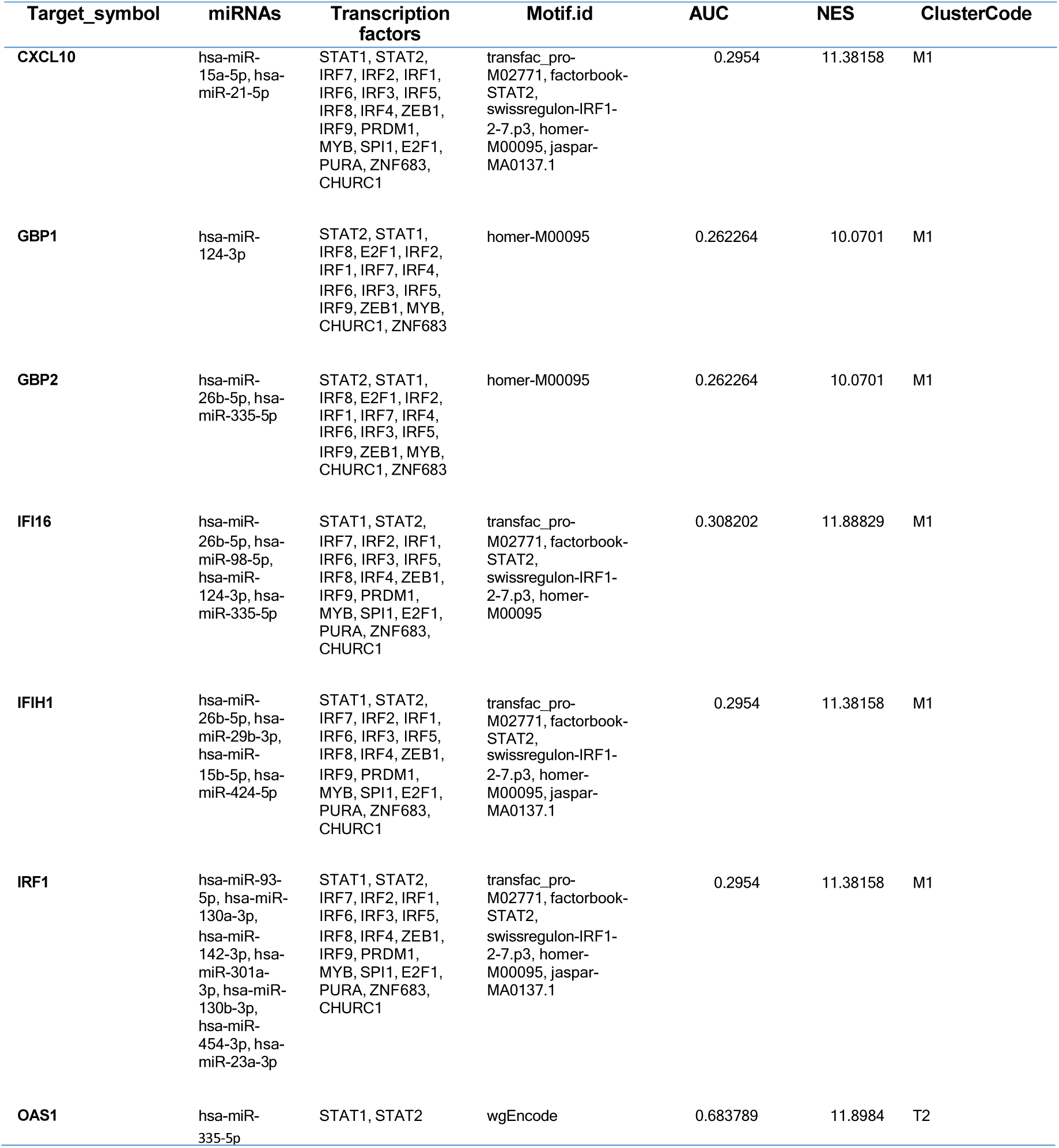
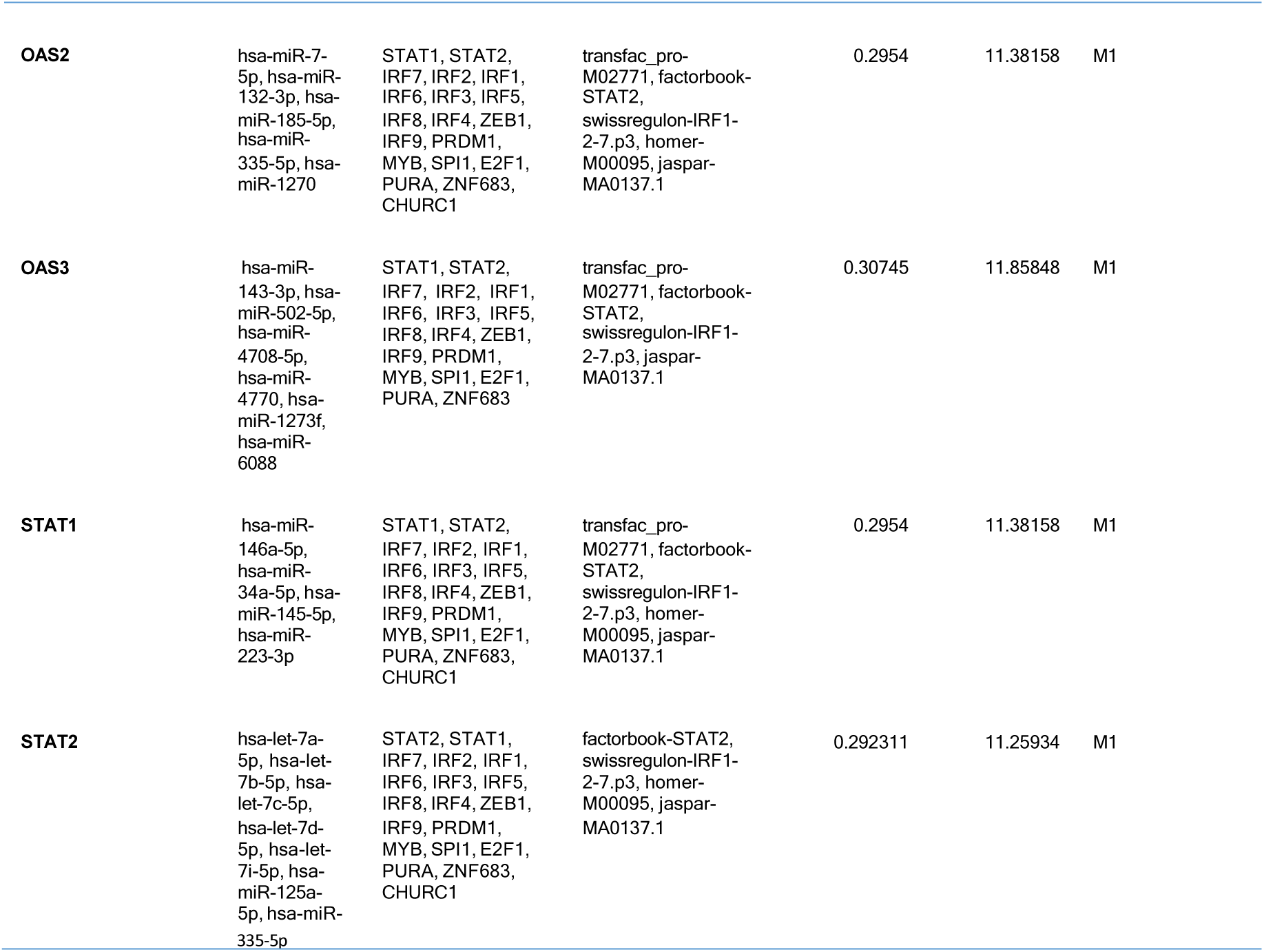
The miRNAs, transcription factors and target gene network enrichment score for hub genes of the blue module.

**Table 5:**
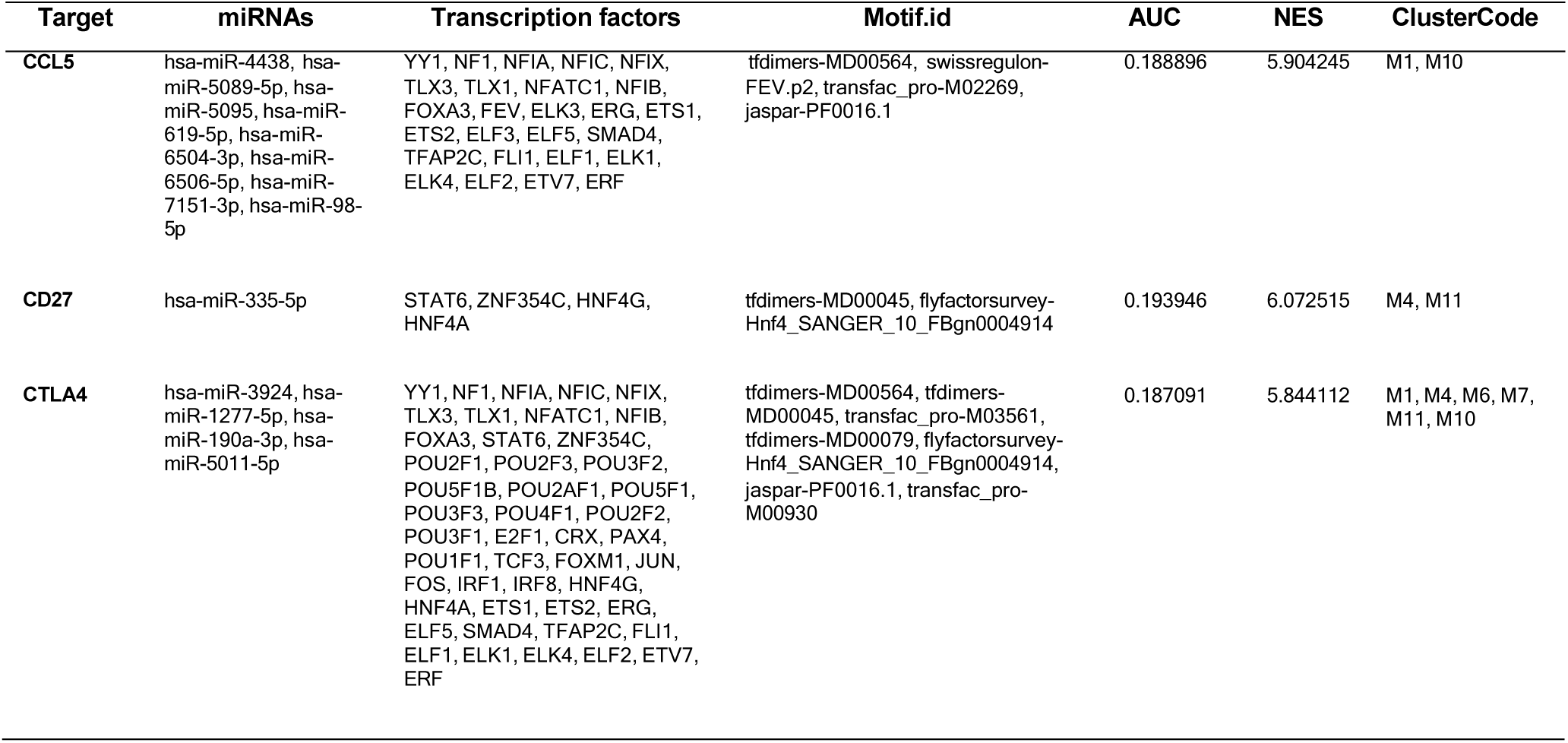
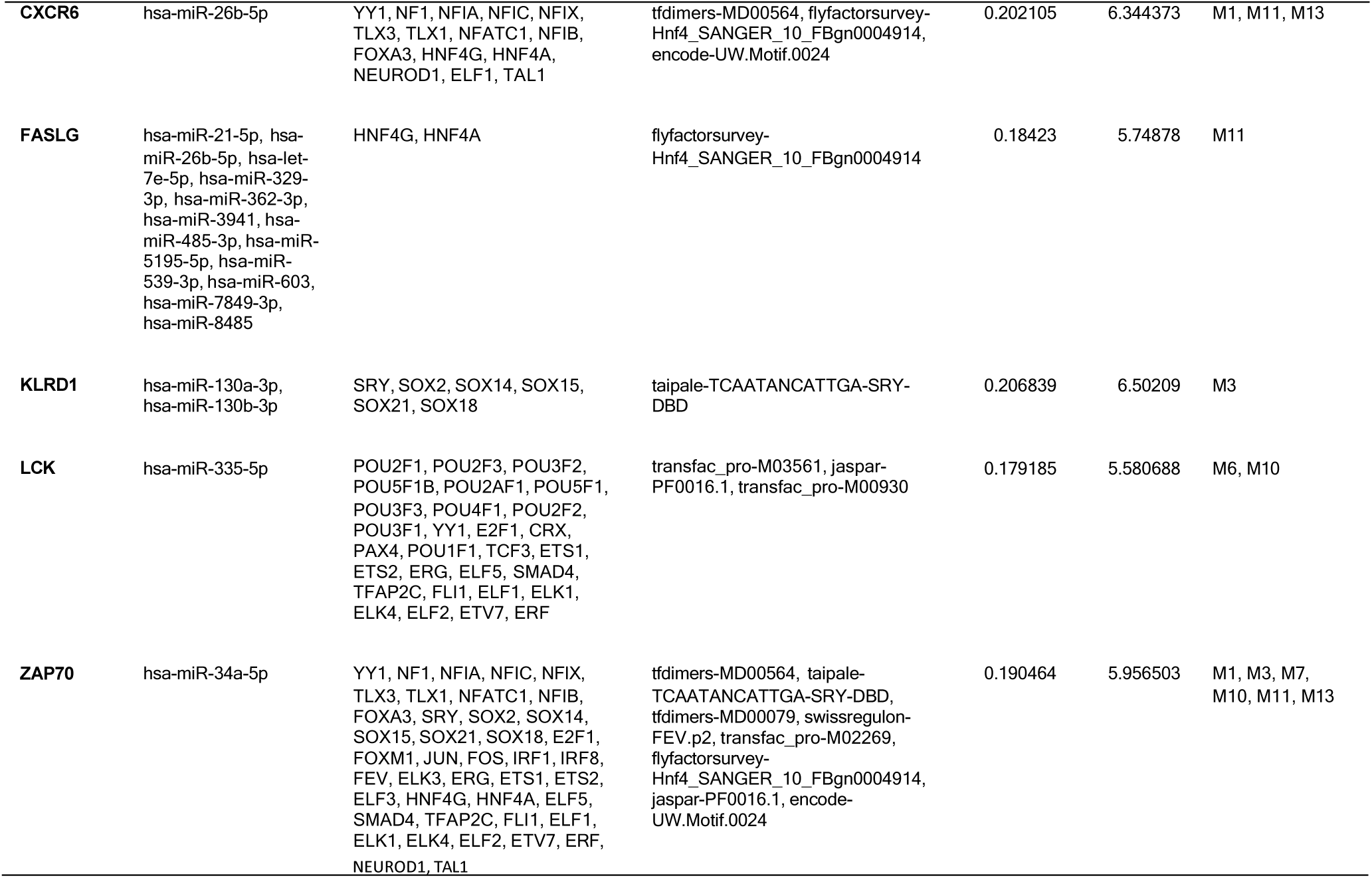
The miRNAs, transcription factors and target gene network enrichment score for hub genes of the green module.

## Discussion

Despite being curable and preventable, TB affects about a quarter of the global population with more than 10 million people falling ill every year [1,23]. Urgent action has been initiated to end the global TB epidemic by 2030 and new vaccines prioritized to rapidly reduce the global number of cases and deaths each year to the levels already achieved in these low-burden countries where successful strategies have been implemented [1]. Effectiveness of vaccination programs in lowering disease rates has been emphasized [2], however, the only licensed vaccine for prevention of TB disease is the widely used Bacille Calmette-Guérin (BCG) vaccine which prevents severe forms of TB in children while there is currently no licensed vaccine that is effective in preventing TB disease in adults, either before or after exposure to TB infection [1]. Even as the M72/AS01E vaccine is now in a Phase III trial, along with other vaccine candidates, their optimization and effective immunization strategies against TB depends heavily depends on the precise identification of molecular signatures active during protection. Transcriptional analyses of PBMCs from M72/AS01E tuberculosis vaccine recipients aims to shed light on vaccine intricacies, offering new horizon in vaccine strategies providing clearer understanding of the elusive specific molecular pathways active in vaccine mechanisms. Dissecting these pathways is vital for pinpointing target regulators and their roles in sustaining vaccination processes and navigating the complexities associated with immune response kinetics, adaptive immunity, and potential correlates of protection against tuberculosis.

In this study, we have utilized network biology approaches which are increasingly applied to identify biomarkers by exploring complex molecular interactions [24]. We harnessed systems and network biology to understand the intricate transcriptome-level landscapes that could unravel potential vaccine strategies in PBMCs. RNA-seq analysis delineated gene expression variances across different timepoints. Through WGCNA, we identified genes in modules notably associated with immune response, specifically pinpointing the blue and green modules as significantly correlated with immnune response. Network analysis further narrowed down 17 hub genes in MYB: CXCL10, DDX58, DHX58, GBP1, GBP2, GBP4, GBP5, IFI16, IFIH1, IRF1, IRF7, IRF9, OAS1, OAS2, OAS3, STAT1, and STAT2 and 14 in MYG: CCL5, CD244, CD27, CTLA4, CXCR6, FASLG, GZMB, IL2RB, KLRD1, LCK, NCR3, PRF1, TBX21, ZAP70.

The identified hub genes are key proteins involved in the immune response to TB, particularly in the context of vaccine efficacy and pathogen recognition. CXCL10, also known as Interferon gamma-induced protein 10, acts as a Th1-chemokine, facilitating the recruitment of leukocytes essential for combating TB. It has emerged as a promising biomarker for differentiating between active and latent TB infections, with elevated levels correlating with disease severity and treatment responses [25]. Early CXCL10 responses noted at D31 in this study correlated with previous observation of early CXCL10 induction during immunization with BCG, ΔsecA2 Mtb mutant, and DNA vaccines [26]. These findings highlight the importance of CXCL10 in TB pathogenesis and its potential as a target for vaccine development and diagnostic applications. Several RNA helicases including DDX58 or RIG-I and DHX58 have been implicated in immune regulation and are critical for detecting viral nucleic acids leading to the production of IFN-I [27]. Interestingly, the role of DExD/H-box helicases in the inflammatory response to bacterial infection remains unclear, and no RNA helicase has been reported to participate in such a process until recently when the role of DDX5 was unraveled to regulate the m6A modification of TLR2/4 mRNA and suppression of inflammatory response pathway during bacterial infection [27]. This potentially establishes that the DExD/H-box helicases including DDX58 may be activated upon Mycobacterium tuberculosis infection contributing to pro-inflammatory cytokine production, while DHX58 may enhance the innate immune response by recognizing mycobacterial RNA, facilitating IFN production. Enhancing the function of DDX58 and DHX58 could improve the efficacy of vaccines and adjuvants by promoting stronger immune responses. Targeting pathways involving both proteins may lead to the development of host-targeted therapeutics that can modulate the immune response in patients with active TB or those vaccinated against it. The Guanylate-binding proteins (GBP1, GBP2, GBP4, GBP5) further enhance the immune defense against TB through various mechanisms, including regulating apoptosis and inflammatory pathways. GBP1, in particular, has demonstrated significant potential as a diagnostic biomarker for TB, showing high sensitivity and specificity in distinguishing active TB from controls [28]. In tuberculosis, GBPs act in a cell-autonomous manner driving vesicular trafficking programs that optimize the delivery of bacteria to degradative compartments, effectively containing and eliminating the pathogen [29]. Importantly, interferon regulatory factors IRF1, IRF7, and IRF9 are crucial for orchestrating the immune response against TB through the regulation of interferon-stimulated genes [30].These factors not only mediate the expression of immune-related genes but also play roles in enhancing vaccine efficacy against TB. Notably, activity of the IRF family members have been reported to involve intricate mechanisms that underpin both their protective and deleterious effects [30]. It therefore surmises a deeper biology may be at play in their significant protective role identified in this study. Future research anticipated in unraveling this interaction would yield valuable insights into the mechanism by which the M72/AS01E candidate vaccine elicit protection to provide pivotal knowledge for advancing TB diagnostics and therapeutics. The 2’-5’-oligoadenylate synthetases (OAS1, OAS2, and OAS3) play crucial roles in the immune response against various pathogens, including M.tb. These interferon-induced genes traditionally associated with antiviral functions, have been found to be upregulated in active TB compared to latent TB infection [31]. Leisching et al. [32] showed that OAS1, OAS2, and OAS3 restrict intracellular M.tb replication and enhance pro-inflammatory cytokine secretion, suggesting a protective role in TB and vaccine efficacy. STAT1 and STAT2 contribute to the immune defense against Mtb. STAT1 plays a pivotal role in mediating the immune response by activating the transcription of various genes essential for fighting infections, including those induced by interferons [33]. STAT2 on the other hand is primarily recognized for its role in the interferon signaling pathway and collaborates with STAT1 to regulate genes critical for a robust immune response to TB, thus providing additional regulatory capacity [33]. Enhanced activity of the STAT proteins may improve protective immunity against TB and help address the challenges posed by latent infections, making them potential targets for novel vaccine development. The immunological roles of CCL5, CD244, CD27, CTLA4, CXCR6, FASLG, GZMB, IL2RB, KLRD1, LCK, NCR3, PRF1, TBX21, and ZAP70 have garnered significant attention in the development of vaccines for TB. These proteins and receptors are integral to the immune response, influencing various aspects such as T cell activation, cytokine production, and the formation of granulomas, which are essential for controlling TB infection. Notably, several of these markers, including CCL5 and CXCR6, have been linked to immune cell recruitment and retention at infection sites, while others like CTLA4 and CD244 play crucial roles in modulating T cell activity and maintaining immune tolerance suggesting potential biomarkers for TB diagnosis and treatment monitoring [34,35]. Exploration of these immune factors may yield novel immunotherapeutic approaches, enhancing the efficacy of existing TB vaccines and fostering the development of new candidates that can elicit robust and durable immune responses against this persistent pathogen. The ability to manipulate immune pathways mediated by these markers could lead to breakthroughs in vaccine efficacy and improved outcomes for TB patients worldwide.

miRNAs play a significant role in regulating the immune response against M. tuberculosis and could serve as novel therapeutic targets in vaccine strategies [36]. They play crucial for modulating both the adaptive and innate immune systems, affecting the differentiation and function of immune cells such as T cells and macrophages [37]. Recent research regarding miRNAs and TB has revealed that the expression profile for particular miRNAs clearly changes upon Mycobacterium tuberculosis infection, vary in the different stages of the disease, and during vaccination [36]. Ruiz-Tagle et. al. [38] reported consistent downregulation of hsa-miR-1270 in serum and sputum samples when comparing samples from patients with active TB and healthy controls while upregulation of hsa-miR-142-3p and hsa-miR-21-5p was reported in PBMC and serum samples. Several of the miRNAs reported in this study have been shown to be biomarkers obtained in vivo or ex vivo from blood cells of TB subjects and controls [36]. These include hsa-let-7e-5p, hsa-let-7a-5p, hsa-let-7b-5p, hsa-miR-125a-5p, hsa-miR-29b-3p, hsa-miR-424-5p, hsa-miR-132-3p, hsa-miR-146a-5p, hsa-miR-223-3p, and hsa-miR-21-5p. hsa-miR-146a-5p was upregulated in M. bovis BCG-infected macrophages from in vitro and in vivo studies while hsa-miR-223-3p directly targets chemokine C-X-C motif ligand 2 (CXCL2), C-C motif ligand 3 (CCL3), IL-6, controls NF-κB activity and negatively regulates cytokine release in TB [36]. Expression of hsa-miR-21-5p is also upregulated in lung macrophages obtained from mice after vaccination with M. bovis BCG and in macrophages and dendritic cells infected ex vivo with M. bovis BCG [36] where it suppresses host Th1 response by targeting IL-12 and promotes dendritic cells’ apoptosis by targeting Bcl-2. hsa-miR-142-3p is downregulated in CD4 T cells and peripheral blood from TB patients, suppresses inflammation [36] and in M. bovis-infected macrophages, negatively regulates the expression of inflammatory cytokines, like NF-κB, TNF-α, and IL-6, in part by targeting the IRAK1 gene. This exploration of miRNAs in the context of TB vaccine development holds considerable promise not only serving as potential therapeutic targets but also as vital components in the development of effective TB vaccines. While significant challenges remain, standardized protocols for miRNA-based diagnostics are necessary to enhance the reliability and applicability of these biomarkers in clinical settings.

Transcription factors play a crucial role in the regulation of gene expression during the immune response to TB infection [39]. This study have highlighted TFs involved in vaccination responses to TB and include PURA, PRDM1, SPI1, E2F1,ZEB1, MYB, IRF3, IRF4, IRF5, IRF2, IRF6,IRF7, IRF8, IRF9, ZNF683, ELF1, and YY1. Interferon Regulatory Factors (IRFs), particularly IRF-1, have been found necessary for normal resistance to mycobacterial infection while IRF8 have been involved in early lung immune response to tuberculosis [39]. Links between miRNA expression, transcription factors and host genes are suggested as regulatory associations in which TF, miRNA, and mRNA can form multiple feed-forward and feed-backward loops in host-pathogen interactions [39]. In the current study, the identified interplay between miRNAs, TFs, and their target genes constitutes a complex regulatory network that governs immune responses illustrating how this interaction regulate cytokine production and immune signaling pathways essential for effective immune defense against M.tb. We found that TFs including PURA, PRDM1, and SPI1 directly regulate 17 miRNAs (Fig. 5G) while OAS2 and OAS3 genes are directly regulated by 12 TFs (Fig. 5H). Additionally, ELF1 and YY1 TFs regulated 8 miRNAs (Fig. 5I) while 6 SOX family of TFs regulate hsa-miR-130b-3p and hsa-miR-130a-3p (Fig. 5I and 5J). The construction of the TF-miRNA-mRNA network provided a good foundation to further distinguish the different mechanisms of immune defense abilities of the M72/AS01E vaccine during vaccination [40]. Future experimental proof of the correlation would be useful in optimizing vaccine strategies and unraveling mechanism of protection.

In conclusion, this study has utilized systems and network biology approaches alongside machine learning to identify key molecular signatures and pathways involved in immune response to the M72/AS01E candidate vaccine against tuberculosis. We highlighted the differential expression patterns of GBP1, GBP2, STAT1, IFIH1, CXCL10, IRF1, GBP5, OAS3, IRF7, OAS1, STAT2, GBP4 in distinguishing vaccinated responders from non-responders, highlighting their significance for advancing vaccine efficacy evaluation. Key pathways altered upon vaccination included NOD-like receptor, RIG-I-like receptor, Toll-like receptor, Chemokine, TNF, and the JAK-STAT signaling pathways, Th1, Th2, and Th17 differentiation, T cell receptor signaling, and cytokine interactions. Further in this study, we pinpointed the potential miRNAs-TFs-target regulatory network driving vaccination response to distinguish the different mechanisms of immune defense abilities of the M72/AS01E vaccine. These findings pave the way to understand the protection correlates of the M72/AS01E vaccine, however, further experimental validation is required to fully elucidate the roles of these biomarkers in vaccine response and to confirm their clinical utility.

## Supporting information

https://drive.google.com/file/d/1eW6qRfTMBVGazXTT9-xPsO9vCHcWIqCW/view?usp=drive_link

## Data Availability

All data produced in the present work are contained in the manuscript

