## Supplementary material for "SYSTEMS AND NETWORK BIOLOGY ANALYSIS COMBINED WITH MACHINE LEARNING IDENTIFIES KEY IMMUNE RESPONSE PROFILES AND POTENTIAL CORRELATES OF PROTECTION FOR THE M72/AS01E TUBERCULOSIS VACCINE": https://drive.google.com/file/d/1eW6qRfTMBVGazXTT9-xPsO9vCHcWIqCW/view?usp=drive_link: Table S1 Differential Gene Expression Analysis (D0 VS D31).docx

| Symbol | padj | pvalue | lfcSE | stat | log2FoldChange | baseMean | Gene Name |
| --- | --- | --- | --- | --- | --- | --- | --- |
| LINGO2 | 7.54E-06 | 1.27E-06 | 0.482 | 4.844416 | 2.335188 | 22.78 | leucine rich repeat and Ig domain containing 2 |
| NUAK1 | 2.27E-04 | 5.89E-05 | 0.5392 | 4.017321 | 2.166214 | 21.48 | NUAK family kinase 1 |
| KRT2 | 1.99E-09 | 1.34E-10 | 0.3362 | 6.422054 | 2.158828 | 12.87 | keratin 2 |
| GFPT2 | 1.45E-05 | 2.67E-06 | 0.452 | 4.694915 | 2.122325 | 14.35 | glutamine-fructose-6-phosphate transaminase 2 |
| LOC105377781 | 9.09E-03 | 3.91E-03 | 0.7203 | 2.884962 | 2.078006 | 33.57 | uncharacterized LOC105377781 |
| LOC105378157 | 1.36E-07 | 1.47E-08 | 0.3661 | 5.665526 | 2.073899 | 44.32 | uncharacterized LOC105378157 |
| KLRC2 | 3.97E-09 | 2.89E-10 | 0.3253 | 6.304711 | 2.050965 | 146.43 | killer cell lectin like receptor C2 |
| KLRF1 | 1.12E-12 | 4.03E-14 | 0.2638 | 7.559992 | 1.994431 | 713.02 | killer cell lectin like receptor F1 |
| DGKK | 1.01E-05 | 1.77E-06 | 0.4149 | 4.77847 | 1.982803 | 51.72 | diacylglycerol kinase kappa |
| NRCAM | 1.20E-06 | 1.63E-07 | 0.372 | 5.237136 | 1.948359 | 75.69 | neuronal cell adhesion molecule |
| KLRC4 | 2.46E-12 | 9.47E-14 | 0.2512 | 7.448132 | 1.870679 | 123.7 | killer cell lectin like receptor C4 |
| KLRC3 | 5.76E-09 | 4.35E-10 | 0.2984 | 6.240854 | 1.86248 | 173.02 | killer cell lectin like receptor C3 |
| LOC107986676 | 2.84E-06 | 4.24E-07 | 0.3676 | 5.05768 | 1.859122 | 11.43 | uncharacterized LOC107986676 |
| XCR1 | 7.62E-08 | 7.71E-09 | 0.3211 | 5.774763 | 1.85431 | 44.68 | X-C motif chemokine receptor 1 |
| UNC45B | 2.21E-14 | 5.90E-16 | 0.2268 | 8.091293 | 1.834809 | 28.9 | unc-45 myosin chaperone B |
| LOC105369656 | 7.00E-07 | 8.95E-08 | 0.341 | 5.346825 | 1.823268 | 25.43 | uncharacterized LOC105369656 |
| EFNA5 | 7.13E-07 | 9.14E-08 | 0.3408 | 5.342972 | 1.820976 | 22.85 | ephrin A5 |
| BNC2 | 2.52E-07 | 2.89E-08 | 0.3281 | 5.54772 | 1.820411 | 41.59 | basonuclin 2 |
| VIT | 5.41E-05 | 1.17E-05 | 0.412 | 4.382871 | 1.805902 | 14.4 | vitrin |
| LIM2 | 3.33E-07 | 3.93E-08 | 0.3285 | 5.49377 | 1.804911 | 16.91 | lens intrinsic membrane protein 2 |
| PCDH1 | 1.06E-09 | 6.81E-11 | 0.2752 | 6.524701 | 1.795683 | 71.63 | protocadherin 1 |
| CYP4F29P | 1.03E-03 | 3.26E-04 | 0.4905 | 3.593741 | 1.762687 | 60.91 | cytochrome P450 family 4 subfamily F member 29, pseudogene |
| RPH3A | 4.59E-05 | 9.68E-06 | 0.3966 | 4.424301 | 1.754729 | 497.62 | rabphilin 3A |
| LOC107984537 | 1.29E-06 | 1.77E-07 | 0.3345 | 5.221582 | 1.746408 | 10.04 | uncharacterized LOC107984537 |
| LOC107985542 | 9.29E-10 | 5.89E-11 | 0.2668 | 6.546615 | 1.746395 | 75.52 |  |
| FGFBP2 | 1.73E-10 | 9.46E-12 | 0.2521 | 6.814482 | 1.717777 | 2247.3 | fibroblast growth factor binding protein 2 |
| LDB2 | 1.37E-05 | 2.50E-06 | 0.363 | 4.708469 | 1.709151 | 19.58 | LIM domain binding 2 |
| LOC105377782 | 1.34E-03 | 4.44E-04 | 0.4858 | 3.512688 | 1.706535 | 158.51 | uncharacterized LOC105377782 |
| LOC107985055 | 4.25E-05 | 8.90E-06 | 0.3839 | 4.442212 | 1.705426 | 24.54 | uncharacterized LOC107985055 |
| LOC101927369 | 4.63E-07 | 5.64E-08 | 0.3136 | 5.429751 | 1.70285 | 63.23 | uncharacterized LOC101927369 |
| IGLV3-13 | 3.49E-03 | 1.32E-03 | 0.529 | 3.211293 | 1.698753 | 12.33 | immunoglobulin lambda variable 3-13 (pseudogene) |
| COLGALT2 | 8.85E-09 | 6.98E-10 | 0.275 | 6.166512 | 1.695975 | 96.51 | collagen beta(1-O)galactosyltransferase 2 |
| PRSS23 | 4.60E-10 | 2.75E-11 | 0.2536 | 6.65946 | 1.688616 | 441.27 | serine protease 23 |
| KLRC1 | 3.72E-08 | 3.46E-09 | 0.2829 | 5.908171 | 1.67166 | 172.44 | killer cell lectin like receptor C1 |
| LOC105373331 | 3.61E-10 | 2.11E-11 | 0.2494 | 6.698315 | 1.670744 | 68.27 | uncharacterized LOC105373331 |
| PGA4 | 3.81E-04 | 1.06E-04 | 0.4307 | 3.87605 | 1.669312 | 16.17 | pepsinogen A4 |
| RNF165 | 6.50E-12 | 2.76E-13 | 0.2279 | 7.305792 | 1.664661 | 106.12 | ring finger protein 165 |
| NMUR1 | 6.03E-17 | 1.14E-18 | 0.1881 | 8.820607 | 1.659504 | 525.77 | neuromedin U receptor 1 |
| PKIB | 2.22E-06 | 3.24E-07 | 0.3241 | 5.108898 | 1.655817 | 12.44 | cAMP-dependent protein kinase inhibitor beta |
| ENPP5 | 3.16E-11 | 1.52E-12 | 0.2333 | 7.073088 | 1.650028 | 116.45 | ectonucleotide pyrophosphatase/phosphodiesterase family member 5 |
| LGR6 | 5.65E-11 | 2.83E-12 | 0.2337 | 6.985995 | 1.632422 | 218.65 | leucine rich repeat containing G protein-coupled receptor 6 |
| LOC105371019 | 1.86E-04 | 4.69E-05 | 0.4001 | 4.07035 | 1.62849 | 13.13 | uncharacterized LOC105371019 |
| HOPX | 8.96E-18 | 1.44E-19 | 0.1797 | 9.049271 | 1.626245 | 747.38 | HOP homeobox |
| SH2D1B | 1.88E-15 | 4.31E-17 | 0.1932 | 8.404123 | 1.62332 | 867.43 | SH2 domain containing 1B |
| SFRP5 | 2.71E-04 | 7.21E-05 | 0.4086 | 3.969102 | 1.621762 | 30.44 | secreted frizzled related protein 5 |
| LOC107984225 | 3.16E-07 | 3.72E-08 | 0.2946 | 5.503536 | 1.621237 | 37.94 | uncharacterized LOC107984225 |
| LOC105375754 | 7.58E-06 | 1.28E-06 | 0.3347 | 4.843051 | 1.621101 | 83.11 | uncharacterized LOC105375754 |
| LOC105372019 | 2.36E-08 | 2.07E-09 | 0.27 | 5.992352 | 1.6182 | 21.78 | uncharacterized LOC105372019 |
| PGA3 | 6.16E-04 | 1.82E-04 | 0.4322 | 3.742253 | 1.61728 | 15.46 | pepsinogen A3 |
| TNR | 8.15E-04 | 2.50E-04 | 0.4387 | 3.662161 | 1.606551 | 10.05 | tenascin R |
| PGA5 | 5.45E-04 | 1.58E-04 | 0.4238 | 3.777452 | 1.600814 | 13.33 | pepsinogen A5 |
| DTHD1 | 2.17E-10 | 1.21E-11 | 0.2359 | 6.779091 | 1.598887 | 283.3 | death domain containing 1 |
| LINC00612 | 5.84E-07 | 7.31E-08 | 0.2967 | 5.383359 | 1.597297 | 13.9 | long intergenic non-protein coding RNA 612 |
| LOC105377019 | 3.07E-05 | 6.18E-06 | 0.3514 | 4.520161 | 1.58832 | 12.14 |  |
| SLC1A7 | 3.23E-05 | 6.55E-06 | 0.3497 | 4.507943 | 1.576497 | 32.3 | solute carrier family 1 member 7 |
| CYP2E1 | 2.06E-08 | 1.78E-09 | 0.261 | 6.016719 | 1.570359 | 27.8 | cytochrome P450 family 2 subfamily E member 1 |
| CENPK | 1.64E-05 | 3.05E-06 | 0.3362 | 4.66748 | 1.569127 | 151.57 | centromere protein K |
| ARHGEF28 | 5.12E-05 | 1.10E-05 | 0.3566 | 4.397086 | 1.567784 | 16.26 | Rho guanine nucleotide exchange factor 28 |
| LINC01801 | 3.18E-05 | 6.42E-06 | 0.3442 | 4.512079 | 1.552965 | 32.39 | long intergenic non-protein coding RNA 1801 |
| LOC105375036 | 3.33E-05 | 6.78E-06 | 0.3449 | 4.500491 | 1.552007 | 99.57 | uncharacterized LOC105375036 |
| LOC107984889 | 1.29E-08 | 1.05E-09 | 0.254 | 6.10106 | 1.549704 | 37.28 | uncharacterized LOC107984889 |
| KRT73 | 6.21E-04 | 1.84E-04 | 0.4144 | 3.739881 | 1.549681 | 119.71 | keratin 73 |
| MYOM2 | 1.83E-02 | 8.70E-03 | 0.5897 | 2.623471 | 1.546965 | 1349.2 | myomesin 2 |
| LRRC43 | 4.17E-05 | 8.71E-06 | 0.3477 | 4.446938 | 1.546082 | 10.65 | leucine rich repeat containing 43 |
| CDNF | 1.11E-08 | 8.98E-10 | 0.2523 | 6.126498 | 1.545716 | 19.22 | cerebral dopamine neurotrophic factor |
| LINC01281 | 3.39E-05 | 6.92E-06 | 0.3432 | 4.496076 | 1.543204 | 12.24 | long intergenic non-protein coding RNA 1281 |
| LOC101929719 | 1.30E-04 | 3.13E-05 | 0.3694 | 4.163598 | 1.537945 | 10.35 | uncharacterized LOC101929719 |
| PRKN | 2.07E-04 | 5.29E-05 | 0.3793 | 4.042279 | 1.533059 | 9.37 | parkin RBR E3 ubiquitin protein ligase |
| RTP5 | 1.43E-05 | 2.61E-06 | 0.3239 | 4.699461 | 1.521937 | 17.44 | receptor transporter protein 5 (putative) |
| CD1E | 6.23E-09 | 4.74E-10 | 0.2439 | 6.227619 | 1.519116 | 41.02 | CD1e molecule |
| SIGLEC17P | 6.54E-15 | 1.62E-16 | 0.1837 | 8.247604 | 1.515239 | 260.34 | sialic acid binding Ig like lectin 17, pseudogene |
| LEXM | 2.51E-07 | 2.89E-08 | 0.272 | 5.547982 | 1.509228 | 25.32 | lymphocyte expansion molecule |
| IGLV3-12 | 3.20E-03 | 1.20E-03 | 0.4637 | 3.239695 | 1.502157 | 43.96 | immunoglobulin lambda variable 3-12 |
| IFI6 | 2.67E-03 | 9.69E-04 | 0.4548 | -3.29939 | -1.50069 | 8641.64 | interferon alpha inducible protein 6 |
| LOC105375035 | 7.53E-05 | 1.70E-05 | 0.3491 | -4.30132 | -1.50158 | 24.02 | uncharacterized LOC105375035 |
| BST2 | 4.96E-12 | 2.06E-13 | 0.2046 | -7.34514 | -1.50311 | 7330.96 | bone marrow stromal cell antigen 2 |
| ZNFX1 | 1.00E-21 | 9.73E-24 | 0.1497 | -10.0443 | -1.50393 | 14532.47 | zinc finger NFX1-type containing 1 |
| IFI44 | 1.10E-03 | 3.52E-04 | 0.4214 | -3.57388 | -1.50616 | 5567.66 | interferon induced protein 44 |
| NMI | 4.33E-17 | 7.99E-19 | 0.1702 | -8.86012 | -1.50776 | 6331.68 | N-myc and STAT interactor |
| PLAUR | 5.40E-15 | 1.32E-16 | 0.1825 | -8.27133 | -1.50919 | 7125.03 | plasminogen activator, urokinase receptor |
| RIPK2 | 3.78E-25 | 2.43E-27 | 0.1393 | -10.8321 | -1.50928 | 1834.11 | receptor interacting serine/threonine kinase 2 |
| CASP4 | 2.49E-20 | 2.82E-22 | 0.1556 | -9.70665 | -1.50989 | 19238.51 | caspase 4 |
| LOC107984880 | 6.97E-10 | 4.32E-11 | 0.2292 | -6.59273 | -1.51136 | 65.09 |  |
| LOC101927283 | 4.35E-06 | 6.82E-07 | 0.3044 | -4.96629 | -1.51193 | 11.95 | uncharacterized LOC101927283 |
| LINC01841 | 1.99E-09 | 1.35E-10 | 0.2355 | -6.42171 | -1.51207 | 20.39 | long intergenic non-protein coding RNA 1841 |
| KIAA0040 | 1.16E-20 | 1.27E-22 | 0.1545 | -9.7875 | -1.51241 | 16576.9 | KIAA0040 |
| PSME1 | 2.41E-30 | 8.13E-33 | 0.1268 | -11.9313 | -1.51317 | 30061.59 | proteasome activator subunit 1 |
| IRAK2 | 1.14E-12 | 4.11E-14 | 0.2003 | -7.55752 | -1.5137 | 720.62 | interleukin 1 receptor associated kinase 2 |
| IFIT2 | 9.96E-07 | 1.33E-07 | 0.288 | -5.27468 | -1.51888 | 28472.08 | interferon induced protein with tetratricopeptide repeats 2 |
| CNDP2 | 7.89E-25 | 5.37E-27 | 0.1412 | -10.7591 | -1.51922 | 8908.04 | carnosine dipeptidase 2 |
| PLEK | 1.52E-21 | 1.50E-23 | 0.152 | -10.0019 | -1.52067 | 36885.2 | pleckstrin |
| MIR9902-1 | 3.48E-05 | 7.12E-06 | 0.3388 | -4.49006 | -1.52105 | 13.96 | microRNA 9902-1 |
| PARP10 | 3.20E-13 | 1.04E-14 | 0.1967 | -7.7337 | -1.52113 | 4388.56 | poly(ADP-ribose) polymerase family member 10 |
| CYBB | 9.69E-25 | 6.65E-27 | 0.1418 | -10.7394 | -1.52296 | 33880.82 | cytochrome b-245 beta chain |
| LRRK2 | 8.53E-13 | 3.00E-14 | 0.2006 | -7.59816 | -1.52436 | 28746.2 | leucine rich repeat kinase 2 |
| ATF5 | 6.22E-21 | 6.71E-23 | 0.1548 | -9.85218 | -1.52559 | 1038.87 | activating transcription factor 5 |
| MLKL | 1.76E-17 | 3.02E-19 | 0.1703 | -8.96814 | -1.52769 | 6181.59 | mixed lineage kinase domain like pseudokinase |
| PCBP1-AS1 | 2.81E-13 | 9.03E-15 | 0.1971 | -7.75223 | -1.52784 | 1130.94 | PCBP1 antisense RNA 1 |
| CRB2 | 6.10E-16 | 1.30E-17 | 0.1788 | -8.54391 | -1.52793 | 292.42 | crumbs cell polarity complex component 2 |
| PADI6 | 6.66E-03 | 2.74E-03 | 0.5117 | -2.99548 | -1.53265 | 26.95 | peptidyl arginine deiminase 6 |
| PSMB8-AS1 | 3.39E-19 | 4.49E-21 | 0.1628 | -9.42058 | -1.5338 | 11520.48 | PSMB8 antisense RNA 1 (head to head) |
| ADM-DT | 2.36E-06 | 3.47E-07 | 0.3011 | -5.09601 | -1.53419 | 38.93 | ADM divergent transcript |
| KCNE5 | 1.90E-07 | 2.11E-08 | 0.2754 | -5.60235 | -1.543 | 31.39 | potassium voltage-gated channel subfamily E regulatory subunit 5 |
| LOC105377449 | 1.79E-13 | 5.49E-15 | 0.1975 | -7.81515 | -1.54377 | 119.67 |  |
| TMEM268 | 1.18E-25 | 6.81E-28 | 0.1413 | -10.9478 | -1.54668 | 2152.57 | transmembrane protein 268 |
| LOC101927243 | 1.00E-05 | 1.75E-06 | 0.3241 | -4.78048 | -1.54946 | 19.24 | uncharacterized LOC101927243 |
| LOC105375924 | 2.24E-08 | 1.95E-09 | 0.2582 | -6.00211 | -1.54998 | 54.33 | uncharacterized LOC105375924 |
| GAS6 | 3.47E-14 | 9.53E-16 | 0.1931 | -8.03272 | -1.55111 | 358.19 | growth arrest specific 6 |
| EGR2 | 4.67E-10 | 2.79E-11 | 0.2332 | -6.65706 | -1.55226 | 68.27 | early growth response 2 |
| LOC107987121 | 1.01E-04 | 2.35E-05 | 0.3676 | -4.22847 | -1.55438 | 13.68 | uncharacterized LOC107987121 |
| OAS2 | 8.02E-08 | 8.16E-09 | 0.2701 | -5.76507 | -1.55686 | 14782.3 | 2'-5'-oligoadenylate synthetase 2 |
| APOL3 | 3.84E-25 | 2.51E-27 | 0.144 | -10.829 | -1.55923 | 8662.86 | apolipoprotein L3 |
| HLX-AS1 | 1.44E-07 | 1.56E-08 | 0.2757 | -5.65517 | -1.55936 | 38.1 | HLX antisense RNA 1 |
| GRINA | 4.45E-22 | 4.07E-24 | 0.1544 | -10.13 | -1.56364 | 12981.61 | glutamate ionotropic receptor NMDA type subunit associated protein 1 |
| RGS1 | 8.24E-07 | 1.07E-07 | 0.2949 | -5.31363 | -1.56702 | 110.53 | regulator of G protein signaling 1 |
| FAM241A | 2.15E-12 | 8.09E-14 | 0.2098 | -7.4688 | -1.56715 | 672.77 | family with sequence similarity 241 member A |
| DHRS9 | 4.79E-11 | 2.36E-12 | 0.2238 | -7.01115 | -1.5691 | 3652.96 | dehydrogenase/reductase 9 |
| FLVCR2 | 4.99E-22 | 4.59E-24 | 0.1553 | -10.1181 | -1.5711 | 913.72 | FLVCR heme transporter 2 |
| TMEM252 | 8.15E-09 | 6.37E-10 | 0.2545 | -6.18106 | -1.57335 | 159.96 | transmembrane protein 252 |
| RCVRN | 1.95E-05 | 3.71E-06 | 0.3403 | -4.62713 | -1.57471 | 17.57 | recoverin |
| BISPR | 8.94E-19 | 1.25E-20 | 0.1691 | -9.31279 | -1.57484 | 1239.83 | BST2 interferon stimulated positive regulator |
| FFAR2 | 2.56E-16 | 5.20E-18 | 0.1822 | -8.64884 | -1.57541 | 22537.19 | free fatty acid receptor 2 |
| WDFY1 | 3.82E-25 | 2.48E-27 | 0.1455 | -10.8302 | -1.57618 | 3847.01 | WD repeat and FYVE domain containing 1 |
| LOC107985279 | 1.42E-03 | 4.72E-04 | 0.4508 | -3.49632 | -1.57626 | 33.1 | uncharacterized LOC107985279 |
| ZNF438 | 6.34E-19 | 8.69E-21 | 0.1686 | -9.35089 | -1.57656 | 2287.72 | zinc finger protein 438 |
| MIR4709 | 5.41E-20 | 6.33E-22 | 0.1644 | -9.62405 | -1.58267 | 1797.04 | microRNA 4709 |
| BCL3 | 1.11E-19 | 1.39E-21 | 0.1661 | -9.54292 | -1.58515 | 5250.01 | BCL3 transcription coactivator |
| CYREN | 7.85E-22 | 7.48E-24 | 0.1575 | -10.0702 | -1.58616 | 11849.17 | cell cycle regulator of NHEJ |
| PSMB10 | 4.73E-25 | 3.17E-27 | 0.1471 | -10.8076 | -1.59017 | 8784.44 | proteasome 20S subunit beta 10 |
| MIR4751 | 2.64E-12 | 1.03E-13 | 0.2145 | -7.43728 | -1.59513 | 48.23 | microRNA 4751 |
| JAK2 | 5.93E-16 | 1.26E-17 | 0.1867 | -8.54731 | -1.59602 | 4343.51 | Janus kinase 2 |
| LOC107984200 | 1.45E-28 | 6.10E-31 | 0.1382 | -11.5664 | -1.599 | 142.94 | uncharacterized LOC107984200 |
| LOC107985047 | 1.88E-05 | 3.55E-06 | 0.3458 | -4.63603 | -1.60335 | 31.97 |  |
| ISG15 | 6.53E-03 | 2.68E-03 | 0.5343 | -3.00221 | -1.60394 | 5086.82 | ISG15 ubiquitin like modifier |
| LINC01531 | 1.39E-09 | 9.16E-11 | 0.2478 | -6.48013 | -1.60566 | 38.95 | long intergenic non-protein coding RNA 1531 |
| PSORS1C3 | 1.05E-02 | 4.60E-03 | 0.567 | -2.83362 | -1.60675 | 37.27 | psoriasis susceptibility 1 candidate 3 |
| TRC-GCA4-1 | 5.72E-07 | 7.14E-08 | 0.2983 | -5.38757 | -1.60723 | 18.67 | tRNA-Cys (anticodon GCA) 4-1 |
| IL15RA | 9.29E-27 | 4.86E-29 | 0.1439 | -11.1845 | -1.60967 | 701.74 | interleukin 15 receptor subunit alpha |
| SNX10 | 2.93E-15 | 6.89E-17 | 0.1931 | -8.34889 | -1.61188 | 5157.82 | sorting nexin 10 |
| LOC101927741 | 1.13E-06 | 1.53E-07 | 0.3071 | -5.24936 | -1.61233 | 117.52 | uncharacterized LOC101927741 |
| BMAL2 | 9.61E-06 | 1.67E-06 | 0.3369 | -4.79003 | -1.61358 | 70.1 | basic helix-loop-helix ARNT like 2 |
| RALB | 9.28E-19 | 1.30E-20 | 0.1735 | -9.30836 | -1.61496 | 16194.95 | RAS like proto-oncogene B |
| TOP1P2 | 1.97E-10 | 1.09E-11 | 0.2379 | -6.79415 | -1.6165 | 12.14 | DNA topoisomerase I pseudogene 2 |
| FFAR3 | 1.10E-05 | 1.94E-06 | 0.3399 | -4.75992 | -1.61813 | 102.83 | free fatty acid receptor 3 |
| SLC38A4-AS1 | 3.37E-07 | 3.99E-08 | 0.2953 | -5.49114 | -1.62173 | 74.56 | SLC38A4 antisense RNA 1 |
| LOC100652833 | 3.58E-05 | 7.36E-06 | 0.362 | -4.48303 | -1.62279 | 12.08 | putative POM121-like protein 1-like |
| H2AC20 | 1.17E-10 | 6.25E-12 | 0.2364 | -6.87379 | -1.625 | 107.35 | H2A clustered histone 20 |
| SNX10-AS1 | 1.68E-13 | 5.11E-15 | 0.2078 | -7.82428 | -1.62622 | 329.01 | SNX10 antisense RNA 1 |
| CASP7 | 7.04E-12 | 3.01E-13 | 0.223 | -7.29389 | -1.62672 | 1711.38 | caspase 7 |
| DDX60L | 1.57E-16 | 3.11E-18 | 0.1869 | -8.7074 | -1.6277 | 14104.77 | DExD/H-box 60 like |
| LOC101929750 | 5.62E-10 | 3.43E-11 | 0.2459 | -6.62689 | -1.62943 | 36.6 | uncharacterized LOC101929750 |
| LOC112268418 | 5.85E-06 | 9.47E-07 | 0.3328 | -4.90234 | -1.63173 | 25.48 | uncharacterized LOC112268418 |
| LGALS3BP | 8.06E-10 | 5.06E-11 | 0.2484 | -6.56906 | -1.63176 | 2401.23 | galectin 3 binding protein |
| DYNLT1 | 1.16E-16 | 2.24E-18 | 0.1868 | -8.74469 | -1.63339 | 5318.06 | dynein light chain Tctex-type 1 |
| NPC2 | 2.39E-25 | 1.46E-27 | 0.1503 | -10.8786 | -1.63476 | 10426.51 | NPC intracellular cholesterol transporter 2 |
| GAS8 | 9.84E-19 | 1.39E-20 | 0.1762 | -9.30128 | -1.63902 | 192.57 | growth arrest specific 8 |
| HERC5 | 1.15E-04 | 2.73E-05 | 0.3917 | -4.1953 | -1.64346 | 6868.33 | HECT and RLD domain containing E3 ubiquitin protein ligase 5 |
| CXCR2P1 | 9.83E-16 | 2.18E-17 | 0.1937 | -8.48367 | -1.64356 | 1685.82 | C-X-C motif chemokine receptor 2 pseudogene 1 |
| TMEM252-DT | 7.40E-08 | 7.45E-09 | 0.2853 | -5.78051 | -1.64901 | 55.37 | TMEM252 divergent transcript |
| RPAP3-DT | 6.47E-20 | 7.86E-22 | 0.1719 | -9.60175 | -1.65027 | 401.05 | RPAP3 divergent transcript |
| SIPA1L1 | 2.97E-21 | 3.05E-23 | 0.1662 | -9.93099 | -1.65056 | 9476.84 | signal induced proliferation associated 1 like 1 |
| DAPP1 | 5.06E-23 | 4.08E-25 | 0.1595 | -10.3525 | -1.65081 | 7453.12 | dual adaptor of phosphotyrosine and 3-phosphoinositides 1 |
| TFEC | 2.82E-10 | 1.61E-11 | 0.2456 | -6.73764 | -1.65456 | 1306.07 | transcription factor EC |
| LINC00513 | 2.86E-05 | 5.71E-06 | 0.3654 | -4.5367 | -1.65767 | 13.32 | long intergenic non-protein coding RNA 513 |
| TTC26 | 1.52E-06 | 2.12E-07 | 0.3204 | -5.18838 | -1.66214 | 289.41 | tetratricopeptide repeat domain 26 |
| DENND1A | 2.54E-28 | 1.11E-30 | 0.1444 | -11.5151 | -1.66263 | 3165.09 | DENN domain containing 1A |
| SHOC1 | 1.57E-15 | 3.59E-17 | 0.1974 | -8.42565 | -1.66295 | 81.15 | shortage in chiasmata 1 |
| MSR1 | 4.48E-04 | 1.27E-04 | 0.4341 | -3.83222 | -1.66365 | 305.42 | macrophage scavenger receptor 1 |
| MT2A | 4.01E-07 | 4.82E-08 | 0.3056 | -5.45772 | -1.66793 | 1749.7 | metallothionein 2A |
| KCNJ2-AS1 | 1.23E-12 | 4.43E-14 | 0.2211 | -7.54778 | -1.66868 | 153.71 | KCNJ2 antisense RNA 1 |
| LOC105372801 | 9.61E-11 | 5.07E-12 | 0.2419 | -6.90355 | -1.66984 | 545.03 | uncharacterized LOC105372801 |
| SQOR | 1.83E-24 | 1.28E-26 | 0.1567 | -10.6785 | -1.67353 | 11692.51 | sulfide quinone oxidoreductase |
| G0S2 | 6.88E-08 | 6.86E-09 | 0.2893 | -5.79436 | -1.67627 | 83.85 | G0/G1 switch 2 |
| GADD45G | 1.66E-13 | 5.06E-15 | 0.2144 | -7.82552 | -1.6781 | 43.47 | growth arrest and DNA damage inducible gamma |
| IFI30 | 2.25E-18 | 3.37E-20 | 0.1825 | -9.20635 | -1.67974 | 60227.68 | IFI30 lysosomal thiol reductase |
| LINC01232 | 1.73E-19 | 2.21E-21 | 0.1769 | -9.49467 | -1.67998 | 236.78 | long intergenic non-protein coding RNA 1232 |
| H1-6 | 9.00E-07 | 1.19E-07 | 0.3174 | -5.29559 | -1.68091 | 12.04 | H1.6 linker histone, cluster member |
| ZMYND15 | 1.29E-10 | 6.97E-12 | 0.2451 | -6.85831 | -1.68124 | 252.49 | zinc finger MYND-type containing 15 |
| LOC105374985 | 3.59E-12 | 1.44E-13 | 0.2275 | -7.39293 | -1.68174 | 372.05 |  |
| HELZ2 | 4.48E-13 | 1.48E-14 | 0.2187 | -7.68895 | -1.68182 | 4809.52 | helicase with zinc finger 2 |
| SBNO2 | 1.25E-21 | 1.22E-23 | 0.1684 | -10.022 | -1.68731 | 10512.17 | strawberry notch homolog 2 |
| LINC00677 | 5.93E-07 | 7.42E-08 | 0.3147 | -5.38058 | -1.69313 | 16.44 | long intergenic non-protein coding RNA 677 |
| NCF1B | 6.08E-20 | 7.28E-22 | 0.1764 | -9.60971 | -1.69538 | 19492.46 | neutrophil cytosolic factor 1B pseudogene |
| CETP | 1.17E-22 | 1.01E-24 | 0.1657 | -10.2654 | -1.7013 | 199.42 | cholesteryl ester transfer protein |
| H2BP1 | 1.02E-09 | 6.53E-11 | 0.2609 | -6.5311 | -1.70394 | 15.17 | H2B histone pseudogene 1 |
| LINC02701 | 1.70E-08 | 1.43E-09 | 0.2816 | -6.05235 | -1.70405 | 28.6 | long intergenic non-protein coding RNA 2701 |
| MFSD6L | 1.67E-11 | 7.60E-13 | 0.2377 | -7.16815 | -1.7041 | 111.66 | major facilitator superfamily domain containing 6 like |
| FHDC1 | 6.14E-13 | 2.10E-14 | 0.2232 | -7.64448 | -1.70605 | 323.67 | FH2 domain containing 1 |
| LOC101926887 | 5.25E-10 | 3.17E-11 | 0.2576 | -6.63831 | -1.71001 | 57.15 | uncharacterized LOC101926887 |
| TNFSF13B | 8.99E-15 | 2.27E-16 | 0.2087 | -8.20711 | -1.71292 | 7559.68 | TNF superfamily member 13b |
| TRANK1 | 1.29E-22 | 1.11E-24 | 0.1671 | -10.2558 | -1.71416 | 21451.45 | tetratricopeptide repeat and ankyrin repeat containing 1 |
| ADAMTSL4-AS2 | 2.27E-18 | 3.42E-20 | 0.1864 | -9.20489 | -1.71541 | 378.59 | ADAMTSL4 antisense RNA 2 |
| NSG2 | 3.29E-05 | 6.69E-06 | 0.3809 | -4.50328 | -1.71548 | 13.81 | neuronal vesicle trafficking associated 2 |
| NUB1 | 2.00E-33 | 4.47E-36 | 0.1368 | -12.5407 | -1.71595 | 8844.35 | negative regulator of ubiquitin like proteins 1 |
| OASL | 1.62E-05 | 3.02E-06 | 0.3681 | -4.66966 | -1.71895 | 5983.03 | 2'-5'-oligoadenylate synthetase like |
| H2AC18 | 4.38E-11 | 2.14E-12 | 0.245 | -7.02537 | -1.7209 | 777.25 | H2A clustered histone 18 |
| LOC105377067 | 1.06E-04 | 2.50E-05 | 0.4088 | -4.21516 | -1.72335 | 409.93 | uncharacterized LOC105377067 |
| H2AC19 | 3.82E-11 | 1.85E-12 | 0.2448 | -7.04506 | -1.72472 | 836.6 | H2A clustered histone 19 |
| NCF1C | 1.46E-17 | 2.47E-19 | 0.192 | -8.9901 | -1.72633 | 20896.34 | neutrophil cytosolic factor 1C pseudogene |
| IFITM1 | 7.82E-16 | 1.70E-17 | 0.2035 | -8.51242 | -1.73192 | 69950.25 | interferon induced transmembrane protein 1 |
| RBCK1 | 5.93E-23 | 4.81E-25 | 0.1676 | -10.3367 | -1.73217 | 11437.3 | RANBP2-type and C3HC4-type zinc finger containing 1 |
| LACTB | 1.10E-27 | 5.04E-30 | 0.1522 | -11.3838 | -1.73255 | 2819.71 | lactamase beta |
| HFE-AS1 | 1.12E-10 | 5.96E-12 | 0.252 | -6.88063 | -1.73382 | 133.99 | HFE antisense RNA 1 |
| ADAMTSL4-AS1 | 2.16E-16 | 4.36E-18 | 0.2001 | -8.66891 | -1.73501 | 226.11 | ADAMTSL4 antisense RNA 1 |
| KCNJ2 | 3.32E-16 | 6.85E-18 | 0.2015 | -8.61741 | -1.73679 | 7426.62 | potassium inwardly rectifying channel subfamily J member 2 |
| NCF1 | 5.26E-15 | 1.28E-16 | 0.21 | -8.27523 | -1.73755 | 34403.63 | neutrophil cytosolic factor 1 |
| URAHP | 4.21E-18 | 6.51E-20 | 0.1903 | -9.13548 | -1.73886 | 55.14 | urate (hydroxyiso-) hydrolase, pseudogene |
| SPATS2L | 2.43E-08 | 2.14E-09 | 0.2906 | -5.9867 | -1.73993 | 581.25 | spermatogenesis associated serine rich 2 like |
| OAS1 | 6.22E-07 | 7.87E-08 | 0.3283 | -5.37014 | -1.76282 | 13115.76 | 2'-5'-oligoadenylate synthetase 1 |
| DDX60 | 1.32E-13 | 3.87E-15 | 0.2243 | -7.85896 | -1.76289 | 4304.49 | DExD/H-box helicase 60 |
| TNFSF10 | 4.85E-17 | 9.02E-19 | 0.1994 | -8.8466 | -1.76405 | 25985.61 | TNF superfamily member 10 |
| RNF213-AS1 | 7.71E-09 | 5.99E-10 | 0.2852 | -6.19082 | -1.76532 | 506.93 | RNF213 antisense RNA 1 |
| LOC101927272 | 1.95E-23 | 1.50E-25 | 0.1692 | -10.4478 | -1.76817 | 265.32 | uncharacterized LOC101927272 |
| NOD2 | 1.57E-15 | 3.57E-17 | 0.2104 | -8.42621 | -1.77286 | 5745.29 | nucleotide binding oligomerization domain containing 2 |
| TRIM21 | 3.48E-25 | 2.18E-27 | 0.1644 | -10.842 | -1.78259 | 10824.63 | tripartite motif containing 21 |
| LOC105374412 | 4.61E-08 | 4.41E-09 | 0.3042 | -5.86822 | -1.78486 | 34.8 | replaced by ID 105374413 |
| RNF213 | 7.33E-23 | 6.02E-25 | 0.1732 | -10.3151 | -1.78656 | 50755.85 | ring finger protein 213 |
| CLEC9A | 1.76E-16 | 3.52E-18 | 0.2056 | -8.69332 | -1.78769 | 311.34 | C-type lectin domain containing 9A |
| TMEM150B | 1.01E-16 | 1.94E-18 | 0.2042 | -8.76095 | -1.78868 | 544.26 | transmembrane protein 150B |
| LOC101927522 | 2.56E-16 | 5.18E-18 | 0.2085 | -8.64942 | -1.80327 | 300.23 | uncharacterized LOC101927522 |
| SLC26A8 | 7.36E-10 | 4.58E-11 | 0.2743 | -6.58396 | -1.80618 | 646.33 | solute carrier family 26 member 8 |
| LIMK2 | 1.96E-23 | 1.51E-25 | 0.1734 | -10.4471 | -1.81147 | 22400.74 | LIM domain kinase 2 |
| FLVCR2-AS1 | 1.67E-17 | 2.84E-19 | 0.2021 | -8.9747 | -1.8135 | 63.17 | FLVCR2 antisense RNA 1 |
| IFI16 | 1.15E-24 | 7.93E-27 | 0.1693 | -10.7231 | -1.8151 | 35767.14 | interferon gamma inducible protein 16 |
| TBC1D30 | 2.45E-17 | 4.38E-19 | 0.2034 | -8.92697 | -1.81533 | 336.24 | TBC1 domain family member 30 |
| LOC107984945 | 3.35E-15 | 7.92E-17 | 0.218 | -8.33237 | -1.81665 | 179.51 | replaced by ID 64744 |
| ACTA2 | 1.50E-15 | 3.39E-17 | 0.2158 | -8.43218 | -1.81942 | 594.94 | actin alpha 2, smooth muscle |
| SOCS3 | 1.01E-14 | 2.56E-16 | 0.2225 | -8.19261 | -1.82282 | 3534.95 | suppressor of cytokine signaling 3 |
| MSRB2 | 1.46E-13 | 4.36E-15 | 0.2326 | -7.84411 | -1.82464 | 1838.89 | methionine sulfoxide reductase B2 |
| CXCL16 | 1.12E-13 | 3.28E-15 | 0.2318 | -7.87988 | -1.82659 | 7818.71 | C-X-C motif chemokine ligand 16 |
| LOC105371873 | 4.37E-08 | 4.16E-09 | 0.3109 | -5.87785 | -1.82742 | 17.14 |  |
| POLB | 3.69E-37 | 4.62E-40 | 0.1383 | -13.2482 | -1.83179 | 2754.99 | DNA polymerase beta |
| AFF1 | 2.06E-25 | 1.24E-27 | 0.1683 | -10.8937 | -1.83342 | 8446.12 | ALF transcription elongation factor 1 |
| SUCNR1 | 1.83E-04 | 4.62E-05 | 0.4508 | -4.07428 | -1.83678 | 119.34 | succinate receptor 1 |
| TNFAIP2 | 1.99E-31 | 5.74E-34 | 0.1513 | -12.1499 | -1.8388 | 48689.28 | TNF alpha induced protein 2 |
| RPS16P5 | 3.48E-17 | 6.32E-19 | 0.2075 | -8.88617 | -1.84419 | 81.01 | ribosomal protein S16 pseudogene 5 |
| SMCO4 | 3.50E-25 | 2.21E-27 | 0.1708 | -10.8405 | -1.85128 | 1560.45 | single-pass membrane protein with coiled-coil domains 4 |
| ACTA2-AS1 | 7.10E-13 | 2.48E-14 | 0.2433 | -7.62307 | -1.85504 | 242.12 | ACTA2 antisense RNA 1 |
| KCNJ10 | 2.61E-06 | 3.87E-07 | 0.3669 | -5.07502 | -1.8618 | 20.11 | potassium inwardly rectifying channel subfamily J member 10 |
| LOC105370635 | 3.50E-09 | 2.51E-10 | 0.2945 | -6.32657 | -1.86346 | 21.49 | uncharacterized LOC105370635 |
| MED12L | 1.08E-18 | 1.53E-20 | 0.201 | -9.29094 | -1.86745 | 102.43 | mediator complex subunit 12L |
| LOC102724237 | 6.84E-06 | 1.13E-06 | 0.3841 | -4.86681 | -1.86945 | 28.05 |  |
| DHRS12 | 4.81E-18 | 7.49E-20 | 0.2054 | -9.12026 | -1.8736 | 2722.1 | dehydrogenase/reductase 12 |
| TRPV4 | 5.85E-11 | 2.95E-12 | 0.2689 | -6.98016 | -1.87723 | 60.47 | transient receptor potential cation channel subfamily V member 4 |
| VSIG10 | 2.24E-08 | 1.96E-09 | 0.313 | -6.00149 | -1.87874 | 327.85 | V-set and immunoglobulin domain containing 10 |
| DTX3L | 5.17E-31 | 1.60E-33 | 0.1564 | -12.0657 | -1.88736 | 13839.57 | deltex E3 ubiquitin ligase 3L |
| MUC1 | 6.47E-20 | 7.83E-22 | 0.1974 | -9.60215 | -1.89553 | 77.15 | mucin 1, cell surface associated |
| PSMB9 | 6.49E-24 | 4.81E-26 | 0.1798 | -10.5552 | -1.89835 | 19010.88 | proteasome 20S subunit beta 9 |
| TMEM140 | 4.46E-21 | 4.73E-23 | 0.1922 | -9.88723 | -1.90036 | 9247.95 | transmembrane protein 140 |
| GADD45B | 1.79E-24 | 1.25E-26 | 0.1782 | -10.6811 | -1.90351 | 4757.54 | growth arrest and DNA damage inducible beta |
| RIGI | 6.38E-16 | 1.38E-17 | 0.2235 | -8.53721 | -1.90822 | 10134.35 | RNA sensor RIG-I |
| MAB21L3 | 8.90E-10 | 5.62E-11 | 0.2914 | -6.5536 | -1.90941 | 24.96 | mab-21 like 3 |
| SOD2 | 2.58E-22 | 2.29E-24 | 0.1876 | -10.186 | -1.91051 | 151790.8 | superoxide dismutase 2 |
| SYNPO2 | 5.70E-12 | 2.39E-13 | 0.2609 | -7.32499 | -1.91127 | 56.88 | synaptopodin 2 |
| LOC105371529 | 2.71E-11 | 1.28E-12 | 0.2706 | -7.09606 | -1.92053 | 205.63 | uncharacterized LOC105371529 |
| IGF2BP3 | 1.38E-17 | 2.32E-19 | 0.2138 | -8.99697 | -1.9239 | 88.24 | insulin like growth factor 2 mRNA binding protein 3 |
| LOC105371461 | 1.47E-11 | 6.60E-13 | 0.2683 | -7.18751 | -1.92851 | 268.76 | replaced by ID 9447 |
| ODF3B | 2.85E-11 | 1.35E-12 | 0.2724 | -7.08861 | -1.93059 | 1030.74 | outer dense fiber of sperm tails 3B |
| MIR194-2HG | 1.91E-08 | 1.63E-09 | 0.3205 | -6.03061 | -1.93251 | 11.26 | MIR194-2 host gene |
| ASPHD2 | 9.15E-29 | 3.58E-31 | 0.167 | -11.6119 | -1.93931 | 693.41 | aspartate beta-hydroxylase domain containing 2 |
| TRIM6 | 2.55E-08 | 2.26E-09 | 0.3247 | -5.9778 | -1.94127 | 100.35 | tripartite motif containing 6 |
| CCDC194 | 1.72E-06 | 2.43E-07 | 0.3761 | -5.16293 | -1.94203 | 16.57 | coiled-coil domain containing 194 |
| LOC102724608 | 6.53E-15 | 1.61E-16 | 0.2355 | -8.24801 | -1.94214 | 244.69 | uncharacterized LOC102724608 |
| KLHDC7B-DT | 4.35E-15 | 1.05E-16 | 0.234 | -8.29943 | -1.94238 | 294.83 | KLHDC7B divergent transcript |
| MDK | 7.17E-11 | 3.68E-12 | 0.2808 | -6.94909 | -1.95149 | 66.84 | midkine |
| PLSCR2 | 1.39E-08 | 1.15E-09 | 0.3207 | -6.08674 | -1.95206 | 22.65 | phospholipid scramblase 2 |
| C4BPA | 8.21E-03 | 3.48E-03 | 0.6696 | -2.92151 | -1.95615 | 1544.61 | complement component 4 binding protein alpha |
| IGSF10 | 2.91E-16 | 5.94E-18 | 0.2267 | -8.63367 | -1.95759 | 34.16 | immunoglobulin superfamily member 10 |
| HTR3B | 2.45E-06 | 3.61E-07 | 0.3855 | -5.08841 | -1.96181 | 18.57 | 5-hydroxytryptamine receptor 3B |
| LINC00189 | 4.37E-08 | 4.15E-09 | 0.3343 | -5.87802 | -1.96495 | 265.48 | long intergenic non-protein coding RNA 189 |
| LOC105374071 | 3.52E-25 | 2.24E-27 | 0.1814 | -10.8393 | -1.9667 | 2035.03 | uncharacterized LOC105374071 |
| IFIH1 | 2.23E-21 | 2.23E-23 | 0.1987 | -9.96215 | -1.97959 | 5275.29 | interferon induced with helicase C domain 1 |
| ASPRV1 | 4.90E-13 | 1.64E-14 | 0.2587 | -7.67585 | -1.98592 | 1572.43 | aspartic peptidase retroviral like 1 |
| H4C5 | 2.92E-07 | 3.41E-08 | 0.3605 | -5.5188 | -1.98967 | 18.92 | H4 clustered histone 5 |
| HCAR2 | 3.23E-20 | 3.69E-22 | 0.2059 | -9.67928 | -1.9931 | 10880.46 | hydroxycarboxylic acid receptor 2 |
| SCO2 | 6.64E-15 | 1.65E-16 | 0.2418 | -8.24543 | -1.99334 | 1292.39 | synthesis of cytochrome C oxidase 2 |
| TCN2 | 2.45E-13 | 7.75E-15 | 0.2579 | -7.77164 | -2.00419 | 881.57 | transcobalamin 2 |
| XAF1 | 4.50E-13 | 1.49E-14 | 0.2612 | -7.68817 | -2.00797 | 10140.02 | XIAP associated factor 1 |
| TAP2 | 9.50E-33 | 2.33E-35 | 0.1619 | -12.4092 | -2.00936 | 17690.67 | transporter 2, ATP binding cassette subfamily B member |
| IFI44L | 2.99E-04 | 8.05E-05 | 0.5098 | -3.94277 | -2.00992 | 4782.51 | interferon induced protein 44 like |
| CLEC6A | 9.35E-09 | 7.43E-10 | 0.3278 | -6.15672 | -2.01841 | 80.61 | C-type lectin domain containing 6A |
| TYMP | 1.11E-14 | 2.83E-16 | 0.2477 | -8.18045 | -2.02608 | 16754.37 | thymidine phosphorylase |
| LOC107985224 | 5.45E-09 | 4.06E-10 | 0.3243 | -6.25167 | -2.02719 | 36.56 | uncharacterized LOC107985224 |
| PLAAT4 | 7.22E-20 | 8.80E-22 | 0.2118 | -9.59009 | -2.03127 | 8885.53 | phospholipase A and acyltransferase 4 |
| CORIN | 1.79E-08 | 1.52E-09 | 0.3398 | -6.04208 | -2.05335 | 73.83 | corin, serine peptidase |
| ERLIN1 | 5.32E-15 | 1.30E-16 | 0.2496 | -8.27366 | -2.0653 | 3247.1 | ER lipid raft associated 1 |
| GBP3 | 3.94E-19 | 5.27E-21 | 0.2201 | -9.40361 | -2.06949 | 3708.48 | guanylate binding protein 3 |
| OAS3 | 1.13E-06 | 1.53E-07 | 0.3945 | -5.24871 | -2.0704 | 16874.98 | 2'-5'-oligoadenylate synthetase 3 |
| AK4 | 2.41E-24 | 1.70E-26 | 0.1951 | -10.6522 | -2.07831 | 92.39 | adenylate kinase 4 |
| LOC107987044 | 5.90E-14 | 1.67E-15 | 0.2613 | -7.96363 | -2.08064 | 17.8 |  |
| KCNJ15 | 3.59E-22 | 3.25E-24 | 0.205 | -10.152 | -2.08094 | 23917.69 | potassium inwardly rectifying channel subfamily J member 15 |
| CARD17P | 7.94E-22 | 7.61E-24 | 0.2069 | -10.0685 | -2.08323 | 885.84 | caspase recruitment domain family member 17, pseudogene |
| RTP4 | 1.43E-13 | 4.25E-15 | 0.2658 | -7.84719 | -2.08576 | 1339.56 | receptor transporter protein 4 |
| NGFR | 2.43E-11 | 1.14E-12 | 0.2934 | -7.11234 | -2.08672 | 45.26 | nerve growth factor receptor |
| BMX | 5.60E-11 | 2.80E-12 | 0.2987 | -6.98723 | -2.08729 | 613.81 | BMX non-receptor tyrosine kinase |
| RSAD2 | 9.61E-04 | 3.02E-04 | 0.5784 | -3.61317 | -2.08973 | 10153.14 | radical S-adenosyl methionine domain containing 2 |
| STX11 | 7.34E-22 | 6.91E-24 | 0.2075 | -10.078 | -2.0909 | 10104.06 | syntaxin 11 |
| HCAR3 | 1.26E-18 | 1.83E-20 | 0.2262 | -9.27189 | -2.09766 | 13388.79 | hydroxycarboxylic acid receptor 3 |
| GRAMD1B | 4.19E-22 | 3.81E-24 | 0.207 | -10.1365 | -2.09859 | 1954.62 | GRAM domain containing 1B |
| GCH1 | 1.01E-20 | 1.11E-22 | 0.2142 | -9.80142 | -2.0995 | 4418.39 | GTP cyclohydrolase 1 |
| MAFF | 1.68E-20 | 1.88E-22 | 0.2156 | -9.74813 | -2.10206 | 637.71 | MAF bZIP transcription factor F |
| STAT2 | 2.36E-26 | 1.27E-28 | 0.1895 | -11.0989 | -2.10376 | 20765 | signal transducer and activator of transcription 2 |
| H4C8 | 6.23E-16 | 1.33E-17 | 0.2464 | -8.54119 | -2.1046 | 131.38 | H4 clustered histone 8 |
| PRMT5-DT | 1.96E-11 | 9.06E-13 | 0.2949 | -7.14411 | -2.10697 | 30.39 | PRMT5 divergent transcript |
| IRF7 | 2.78E-12 | 1.09E-13 | 0.2855 | -7.42996 | -2.12106 | 5637.85 | interferon regulatory factor 7 |
| TRAFD1 | 9.17E-29 | 3.64E-31 | 0.183 | -11.6105 | -2.12485 | 12910.73 | TRAF-type zinc finger domain containing 1 |
| RAB20 | 4.97E-17 | 9.29E-19 | 0.2407 | -8.84339 | -2.12827 | 1288.98 | RAB20, member RAS oncogene family |
| KREMEN1 | 1.85E-11 | 8.50E-13 | 0.2979 | -7.15288 | -2.13111 | 3913.47 | kringle containing transmembrane protein 1 |
| CD59 | 3.49E-21 | 3.63E-23 | 0.2155 | -9.91372 | -2.13607 | 9148.72 | CD59 molecule (CD59 blood group) |
| CMPK2 | 3.00E-06 | 4.50E-07 | 0.4233 | -5.04629 | -2.13619 | 3180.19 | cytidine/uridine monophosphate kinase 2 |
| EGR3 | 4.12E-13 | 1.36E-14 | 0.2776 | -7.70027 | -2.13743 | 91.23 | early growth response 3 |
| MIR7703 | 6.83E-27 | 3.50E-29 | 0.1909 | -11.2136 | -2.14025 | 659.29 | microRNA 7703 |
| MIR4257 | 4.82E-14 | 1.35E-15 | 0.2683 | -7.99023 | -2.14401 | 57.86 | microRNA 4257 |
| TRIM22 | 6.14E-24 | 4.48E-26 | 0.204 | -10.5618 | -2.15485 | 34864.46 | tripartite motif containing 22 |
| C1QB | 1.08E-06 | 1.45E-07 | 0.4119 | -5.25899 | -2.16611 | 449.31 | complement C1q B chain |
| XRN1 | 1.21E-27 | 5.67E-30 | 0.1905 | -11.3734 | -2.16654 | 6815.68 | 5'-3' exoribonuclease 1 |
| RHBDF2 | 3.37E-35 | 5.87E-38 | 0.1688 | -12.8795 | -2.17456 | 7278.57 | rhomboid 5 homolog 2 |
| CACNA1E | 6.07E-21 | 6.51E-23 | 0.2207 | -9.85515 | -2.17533 | 604.08 | calcium voltage-gated channel subunit alpha1 E |
| LOC112268267 | 1.55E-27 | 7.51E-30 | 0.1919 | -11.3489 | -2.17812 | 8645.77 | uncharacterized LOC112268267 |
| C15orf48 | 1.75E-08 | 1.48E-09 | 0.3628 | -6.04681 | -2.19354 | 33.06 | chromosome 15 open reading frame 48 |
| GRIN3A | 2.95E-21 | 3.01E-23 | 0.221 | -9.93224 | -2.19517 | 235.39 | glutamate ionotropic receptor NMDA type subunit 3A |
| CCR5AS | 9.81E-14 | 2.83E-15 | 0.2786 | -7.89811 | -2.20059 | 196.07 | CCR5 antisense RNA |
| FAS | 2.82E-29 | 1.07E-31 | 0.188 | -11.7146 | -2.20248 | 7231.9 | Fas cell surface death receptor |
| IRF1 | 6.31E-34 | 1.30E-36 | 0.1747 | -12.6379 | -2.20837 | 64252.97 | interferon regulatory factor 1 |
| ICAM1 | 8.30E-32 | 2.22E-34 | 0.1814 | -12.2276 | -2.21822 | 5997.55 | intercellular adhesion molecule 1 |
| RMI2 | 1.28E-19 | 1.61E-21 | 0.2328 | -9.52763 | -2.21841 | 262.91 | RecQ mediated genome instability 2 |
| LOC105370355 | 6.68E-12 | 2.84E-13 | 0.3054 | -7.3017 | -2.23021 | 79.99 | uncharacterized LOC105370355 |
| PSME2 | 1.67E-35 | 2.46E-38 | 0.1724 | -12.9466 | -2.2325 | 16630.48 | proteasome activator subunit 2 |
| C11orf91 | 7.52E-16 | 1.63E-17 | 0.2628 | -8.51728 | -2.23811 | 35.31 | chromosome 11 open reading frame 91 |
| STK3 | 3.44E-26 | 1.87E-28 | 0.2025 | -11.0641 | -2.24002 | 926.56 | serine/threonine kinase 3 |
| APOL2 | 1.25E-34 | 2.32E-37 | 0.1756 | -12.7731 | -2.24311 | 14925.94 | apolipoprotein L2 |
| VPS9D1-AS1 | 8.28E-28 | 3.74E-30 | 0.1972 | -11.4097 | -2.2503 | 517.3 | VPS9D1 antisense RNA 1 |
| PARP9 | 1.46E-28 | 6.21E-31 | 0.1949 | -11.5648 | -2.25453 | 20003.44 | poly(ADP-ribose) polymerase family member 9 |
| IFIT3 | 3.99E-08 | 3.75E-09 | 0.3829 | -5.8949 | -2.25744 | 32442.12 | interferon induced protein with tetratricopeptide repeats 3 |
| IL27 | 4.44E-17 | 8.22E-19 | 0.255 | -8.85699 | -2.25894 | 103.23 | interleukin 27 |
| TAP1 | 7.50E-33 | 1.80E-35 | 0.1822 | -12.43 | -2.26485 | 40646.38 | transporter 1, ATP binding cassette subfamily B member |
| C1QC | 2.24E-05 | 4.32E-06 | 0.4932 | -4.59551 | -2.2666 | 126.81 | complement C1q C chain |
| HMGA2-AS1 | 7.93E-11 | 4.11E-12 | 0.3286 | -6.93333 | -2.27839 | 32.8 | HMGA2 antisense RNA 1 |
| NUCB1 | 3.24E-31 | 9.69E-34 | 0.1888 | -12.1071 | -2.28572 | 18353.77 | nucleobindin 1 |
| NECTIN2 | 9.35E-05 | 2.16E-05 | 0.5399 | -4.24743 | -2.29329 | 1118.38 | nectin cell adhesion molecule 2 |
| SORT1 | 1.22E-28 | 4.91E-31 | 0.198 | -11.585 | -2.29359 | 6915.12 | sortilin 1 |
| DOCK4 | 1.17E-19 | 1.46E-21 | 0.2408 | -9.53766 | -2.2965 | 1313.11 | dedicator of cytokinesis 4 |
| GBP2 | 1.57E-37 | 1.79E-40 | 0.1726 | -13.3192 | -2.2993 | 48844.39 | guanylate binding protein 2 |
| RUFY4 | 4.26E-19 | 5.73E-21 | 0.2449 | -9.39491 | -2.30055 | 196.98 | RUN and FYVE domain containing 4 |
| CXCL11 | 1.02E-06 | 1.36E-07 | 0.439 | -5.2708 | -2.31411 | 15.18 | C-X-C motif chemokine ligand 11 |
| LOC105378841 | 4.05E-31 | 1.24E-33 | 0.1919 | -12.0871 | -2.31919 | 299.62 | uncharacterized LOC105378841 |
| LINC01094 | 3.39E-15 | 8.04E-17 | 0.279 | -8.33061 | -2.32402 | 368.18 | long intergenic non-protein coding RNA 1094 |
| FBXO39 | 2.82E-05 | 5.62E-06 | 0.5124 | -4.54007 | -2.32621 | 20.01 | F-box protein 39 |
| NUCB1-AS1 | 2.43E-34 | 4.63E-37 | 0.183 | -12.7192 | -2.32779 | 1983.27 | NUCB1 antisense RNA 1 |
| IFITM3 | 1.19E-07 | 1.27E-08 | 0.4103 | -5.69014 | -2.33449 | 64041 | interferon induced transmembrane protein 3 |
| SDC3 | 3.25E-17 | 5.86E-19 | 0.2628 | -8.89458 | -2.33778 | 350.16 | syndecan 3 |
| PML | 1.30E-25 | 7.59E-28 | 0.2137 | -10.9379 | -2.33794 | 8273.46 | PML nuclear body scaffold |
| EXOC3L1 | 2.93E-06 | 4.38E-07 | 0.4646 | -5.05146 | -2.34715 | 71.06 | exocyst complex component 3 like 1 |
| CCRL2 | 2.60E-21 | 2.63E-23 | 0.2365 | -9.94572 | -2.35236 | 676.83 | C-C motif chemokine receptor like 2 |
| LOC105374304 | 4.28E-25 | 2.82E-27 | 0.2189 | -10.8184 | -2.3678 | 110.65 | uncharacterized LOC105374304 |
| CDHR5 | 4.56E-12 | 1.86E-13 | 0.3221 | -7.35839 | -2.37012 | 24.77 | cadherin related family member 5 |
| APOL6 | 8.01E-37 | 1.05E-39 | 0.1801 | -13.1867 | -2.37452 | 15613.96 | apolipoprotein L6 |
| VPS9D1 | 2.89E-27 | 1.45E-29 | 0.2109 | -11.2915 | -2.38164 | 2761.48 | VPS9 domain containing 1 |
| PRRG4 | 1.20E-30 | 4.00E-33 | 0.2005 | -11.9902 | -2.4039 | 3646.58 | proline rich and Gla domain 4 |
| GK3 | 3.42E-25 | 2.12E-27 | 0.2253 | -10.8443 | -2.44271 | 105.43 | glycerol kinase 3 |
| SAMD9L | 2.07E-27 | 1.01E-29 | 0.2164 | -11.3227 | -2.45012 | 18045.02 | sterile alpha motif domain containing 9 like |
| SAMD4A | 8.59E-22 | 8.27E-24 | 0.2437 | -10.0603 | -2.45175 | 602.79 | sterile alpha motif domain containing 4A |
| ANXA3 | 3.66E-14 | 1.01E-15 | 0.3058 | -8.02578 | -2.45412 | 6905.42 | annexin A3 |
| PLSCR1 | 1.28E-20 | 1.42E-22 | 0.2523 | -9.77673 | -2.46626 | 8257.69 | phospholipid scramblase 1 |
| SLC6A12-AS1 | 6.62E-13 | 2.30E-14 | 0.3249 | -7.63281 | -2.48007 | 18.07 | SLC6A12 antisense RNA 1 |
| KLF5 | 2.19E-17 | 3.86E-19 | 0.2778 | -8.94079 | -2.48336 | 534.55 | KLF transcription factor 5 |
| TIMM10 | 1.12E-15 | 2.51E-17 | 0.2959 | -8.46738 | -2.50585 | 1797.84 | translocase of inner mitochondrial membrane 10 |
| TGM2 | 1.57E-18 | 2.31E-20 | 0.2724 | -9.24707 | -2.51903 | 783.79 | transglutaminase 2 |
| STAT1 | 4.83E-49 | 1.05E-52 | 0.1654 | -15.2793 | -2.52726 | 48636.26 | signal transducer and activator of transcription 1 |
| LOC105378085 | 5.80E-11 | 2.91E-12 | 0.362 | -6.9819 | -2.52732 | 157.89 | uncharacterized LOC105378085 |
| FBXO6 | 1.20E-30 | 3.96E-33 | 0.211 | -11.9909 | -2.52962 | 2457.42 | F-box protein 6 |
| LOC107986193 | 8.00E-17 | 1.52E-18 | 0.288 | -8.78823 | -2.53139 | 16.89 | uncharacterized LOC107986193 |
| LOC105369593 | 1.03E-05 | 1.81E-06 | 0.5318 | -4.77351 | -2.53864 | 24.01 | uncharacterized LOC105369593 |
| AIM2 | 3.71E-30 | 1.29E-32 | 0.2141 | -11.8927 | -2.54599 | 2297.04 | absent in melanoma 2 |
| SLC6A12 | 1.50E-19 | 1.90E-21 | 0.2695 | -9.51051 | -2.56297 | 669.03 | solute carrier family 6 member 12 |
| EPSTI1 | 4.97E-16 | 1.05E-17 | 0.2999 | -8.56869 | -2.5697 | 8582.47 | epithelial stromal interaction 1 |
| LPCAT2 | 3.46E-26 | 1.90E-28 | 0.2327 | -11.0627 | -2.57393 | 11088.02 | lysophosphatidylcholine acyltransferase 2 |
| GK | 6.91E-30 | 2.48E-32 | 0.2176 | -11.838 | -2.57638 | 10543.18 | glycerol kinase |
| LOC112268296 | 2.76E-25 | 1.70E-27 | 0.239 | -10.8646 | -2.59663 | 865.05 |  |
| CCL2 | 6.23E-09 | 4.74E-10 | 0.4176 | -6.22749 | -2.60081 | 63.34 | C-C motif chemokine ligand 2 |
| CD300LD | 5.92E-08 | 5.82E-09 | 0.4471 | -5.82192 | -2.60286 | 160.67 | CD300 molecule like family member d |
| AANAT | 2.24E-22 | 1.97E-24 | 0.2572 | -10.2004 | -2.62314 | 132.99 | aralkylamine N-acetyltransferase |
| TIFA | 2.47E-30 | 8.48E-33 | 0.2214 | -11.9278 | -2.64031 | 2811.19 | TRAF interacting protein with forkhead associated domain |
| IFI35 | 4.30E-23 | 3.42E-25 | 0.2547 | -10.3694 | -2.64126 | 8048.19 | interferon induced protein 35 |
| CFAP97D1 | 1.15E-14 | 2.94E-16 | 0.3237 | -8.1759 | -2.64667 | 14.85 | CFAP97 domain containing 1 |
| PXT1 | 1.94E-09 | 1.30E-10 | 0.4124 | -6.42684 | -2.65021 | 18.3 | peroxisomal testis enriched protein 1 |
| CAPNS2 | 1.88E-14 | 4.96E-16 | 0.328 | -8.11244 | -2.66101 | 32.89 | calpain small subunit 2 |
| PARP14 | 8.67E-32 | 2.36E-34 | 0.218 | -12.2224 | -2.66413 | 40380.12 | poly(ADP-ribose) polymerase family member 14 |
| UBE2L6 | 8.21E-34 | 1.74E-36 | 0.2114 | -12.6151 | -2.66738 | 28934.06 | ubiquitin conjugating enzyme E2 L6 |
| METTL7B | 4.22E-09 | 3.09E-10 | 0.4251 | -6.29414 | -2.67552 | 36.51 | methyltransferase like 7B |
| FGF13 | 2.30E-08 | 2.01E-09 | 0.4462 | -5.99691 | -2.67569 | 115.82 | fibroblast growth factor 13 |
| LOC101929319 | 2.08E-18 | 3.09E-20 | 0.2921 | -9.21595 | -2.69171 | 373.48 | uncharacterized LOC101929319 |
| LHFPL2 | 1.17E-30 | 3.74E-33 | 0.2261 | -11.9956 | -2.7126 | 2595.58 | LHFPL tetraspan subfamily member 2 |
| SECTM1 | 1.35E-28 | 5.59E-31 | 0.2365 | -11.5739 | -2.73731 | 30491.42 | secreted and transmembrane 1 |
| APOL1 | 2.80E-42 | 2.44E-45 | 0.1939 | -14.1313 | -2.74006 | 7710.71 | apolipoprotein L1 |
| LYPD5 | 1.41E-17 | 2.37E-19 | 0.3056 | -8.99466 | -2.7487 | 33.79 | LY6/PLAUR domain containing 5 |
| XXYLT1-AS2 | 1.20E-23 | 9.10E-26 | 0.2637 | -10.4951 | -2.76795 | 68.56 | XXYLT1 antisense RNA 2 |
| FRMD3 | 1.67E-35 | 2.55E-38 | 0.2141 | -12.9436 | -2.7713 | 714.91 | FERM domain containing 3 |
| SNHG28 | 1.69E-28 | 7.25E-31 | 0.2404 | -11.5515 | -2.77736 | 310.63 | small nucleolar RNA host gene 28 |
| ATP1B2 | 1.39E-13 | 4.11E-15 | 0.3543 | -7.85157 | -2.78207 | 64.35 | ATPase Na+/K+ transporting subunit beta 2 |
| GPR84 | 5.09E-32 | 1.30E-34 | 0.2282 | -12.2706 | -2.80044 | 317.71 | G protein-coupled receptor 84 |
| PSTPIP2 | 1.24E-35 | 1.69E-38 | 0.2166 | -12.9754 | -2.81013 | 8510.27 | proline-serine-threonine phosphatase interacting protein 2 |
| CASP5 | 8.62E-27 | 4.46E-29 | 0.2547 | -11.1921 | -2.851 | 2217.25 | caspase 5 |
| FAM225A | 5.58E-20 | 6.62E-22 | 0.2969 | -9.61939 | -2.85588 | 270.18 | family with sequence similarity 225 member A |
| TNFAIP6 | 5.48E-16 | 1.16E-17 | 0.3345 | -8.55679 | -2.86242 | 5000.08 | TNF alpha induced protein 6 |
| LOC105374898 | 2.78E-15 | 6.52E-17 | 0.3478 | -8.35549 | -2.90628 | 91.57 | uncharacterized LOC105374898 |
| FAM225B | 5.53E-22 | 5.15E-24 | 0.2877 | -10.1069 | -2.90767 | 191.12 | family with sequence similarity 225 member B |
| FAP | 2.41E-09 | 1.66E-10 | 0.4571 | -6.3897 | -2.92076 | 16.01 | fibroblast activation protein alpha |
| BCL2L14 | 1.60E-12 | 5.89E-14 | 0.3901 | -7.51056 | -2.92959 | 30.09 | BCL2 like 14 |
| ZDHHC19 | 6.45E-24 | 4.74E-26 | 0.2866 | -10.5565 | -3.02538 | 147.94 | zinc finger DHHC-type palmitoyltransferase 19 |
| SCARF1 | 2.51E-43 | 1.64E-46 | 0.2143 | -14.32 | -3.06945 | 3918.57 | scavenger receptor class F member 1 |
| MYOF | 2.16E-35 | 3.52E-38 | 0.2377 | -12.9189 | -3.07043 | 4292.22 | myoferlin |
| EFCAB2 | 3.68E-34 | 7.31E-37 | 0.243 | -12.6834 | -3.08227 | 1071.64 | EF-hand calcium binding domain 2 |
| GSDMC | 7.05E-10 | 4.38E-11 | 0.4681 | -6.59077 | -3.08536 | 16.83 | gasdermin C |
| CEACAM1 | 2.16E-35 | 3.47E-38 | 0.2413 | -12.9201 | -3.11766 | 5842.13 | CEA cell adhesion molecule 1 |
| CFB | 5.55E-20 | 6.55E-22 | 0.328 | -9.62049 | -3.15556 | 39.71 | complement factor B |
| HCAR1 | 1.38E-13 | 4.07E-15 | 0.4031 | -7.85282 | -3.16525 | 29.64 | hydroxycarboxylic acid receptor 1 |
| C2 | 1.26E-28 | 5.15E-31 | 0.2746 | -11.5809 | -3.18036 | 822.81 | complement C2 |
| SLAMF8 | 5.35E-33 | 1.25E-35 | 0.2581 | -12.4588 | -3.21516 | 1366.71 | SLAM family member 8 |
| LINC01093 | 6.70E-10 | 4.13E-11 | 0.4897 | -6.59917 | -3.23184 | 75.54 | long intergenic non-protein coding RNA 1093 |
| BNIP5 | 2.67E-11 | 1.26E-12 | 0.4559 | -7.0983 | -3.23644 | 28.88 | BCL2 interacting protein 5 |
| LAP3 | 3.23E-31 | 9.49E-34 | 0.2708 | -12.1088 | -3.27929 | 17110.25 | leucine aminopeptidase 3 |
| LOC105371082 | 5.29E-31 | 1.67E-33 | 0.2721 | -12.0623 | -3.28157 | 243.59 | uncharacterized LOC105371082 |
| CXCL9 | 3.41E-14 | 9.36E-16 | 0.4095 | -8.03495 | -3.29048 | 152.8 | C-X-C motif chemokine ligand 9 |
| LOC105374296 | 7.53E-30 | 2.75E-32 | 0.279 | -11.8295 | -3.30014 | 62.66 | uncharacterized LOC105374296 |
| PANDAR | 9.04E-32 | 2.51E-34 | 0.2722 | -12.2174 | -3.32564 | 237.64 | promoter of CDKN1A antisense DNA damage activated RNA |
| NRN1 | 1.21E-11 | 5.40E-13 | 0.465 | -7.21482 | -3.35488 | 122.06 | neuritin 1 |
| GBP7 | 3.56E-21 | 3.72E-23 | 0.3418 | -9.91131 | -3.38752 | 81.43 | guanylate binding protein 7 |
| SOCS1 | 8.36E-26 | 4.78E-28 | 0.3095 | -10.9798 | -3.3979 | 675.58 | suppressor of cytokine signaling 1 |
| GBP4 | 1.46E-48 | 5.55E-52 | 0.2243 | -15.1704 | -3.40252 | 19952.54 | guanylate binding protein 4 |
| VAMP5 | 3.68E-34 | 7.41E-37 | 0.2691 | -12.6823 | -3.41288 | 4412.75 | vesicle associated membrane protein 5 |
| CALHM6 | 2.59E-35 | 4.37E-38 | 0.2688 | -12.9023 | -3.46791 | 3485.92 | calcium homeostasis modulator family member 6 |
| LRRK2-DT | 1.29E-27 | 6.18E-30 | 0.3068 | -11.3659 | -3.48667 | 63.54 | LRRK2 divergent transcript |
| WARS1 | 1.04E-48 | 3.40E-52 | 0.2348 | -15.2026 | -3.56922 | 82747.15 | tryptophanyl-tRNA synthetase 1 |
| IL31RA | 2.61E-21 | 2.65E-23 | 0.3625 | -9.94496 | -3.60475 | 382.53 | interleukin 31 receptor A |
| SEPTIN4-AS1 | 4.13E-30 | 1.46E-32 | 0.3106 | -11.8825 | -3.69018 | 65.47 | SEPTIN4 antisense RNA 1 |
| LINC02555 | 3.79E-33 | 8.67E-36 | 0.2977 | -12.4881 | -3.7173 | 944.78 | long intergenic non-protein coding RNA 2555 |
| H2BC18 | 1.04E-42 | 8.49E-46 | 0.2622 | -14.2054 | -3.72512 | 1112.66 | H2B clustered histone 18 |
| IDO2 | 3.08E-28 | 1.36E-30 | 0.3291 | -11.4974 | -3.78435 | 64.41 | indoleamine 2,3-dioxygenase 2 |
| LOC100996318 | 1.61E-48 | 7.02E-52 | 0.2498 | -15.155 | -3.78555 | 712.16 | uncharacterized LOC100996318 |
| SMTNL1 | 2.10E-22 | 1.84E-24 | 0.3711 | -10.2073 | -3.78827 | 623.45 | smoothelin like 1 |
| GBP1 | 9.74E-46 | 5.83E-49 | 0.2629 | -14.7068 | -3.86645 | 40895.95 | guanylate binding protein 1 |
| FCGR1BP | 2.29E-49 | 3.73E-53 | 0.2531 | -15.3466 | -3.88389 | 9390.2 | Fc gamma receptor Ib, pseudogene |
| H2BP2 | 2.77E-43 | 1.96E-46 | 0.2744 | -14.3076 | -3.92599 | 664.09 | H2B histone pseudogene 2 |
| ATF3 | 2.36E-39 | 2.31E-42 | 0.2933 | -13.6402 | -4.00024 | 832.65 | activating transcription factor 3 |
| LAMP3 | 2.16E-27 | 1.07E-29 | 0.3589 | -11.318 | -4.06171 | 1332.97 | lysosomal associated membrane protein 3 |
| GBP5 | 3.95E-50 | 4.30E-54 | 0.2629 | -15.4862 | -4.07066 | 91021.4 | guanylate binding protein 5 |
| LOC112268237 | 5.51E-47 | 2.70E-50 | 0.2739 | -14.9134 | -4.08427 | 176.28 |  |
| SEPTIN4 | 1.67E-35 | 2.39E-38 | 0.3164 | -12.9487 | -4.0973 | 819.75 | septin 4 |
| CD274 | 2.58E-32 | 6.47E-35 | 0.3381 | -12.3271 | -4.16746 | 5870.77 | CD274 molecule |
| P2RY14 | 7.71E-43 | 5.87E-46 | 0.2937 | -14.2311 | -4.17989 | 5421.43 | purinergic receptor P2Y14 |
| FCGR1CP | 1.04E-48 | 3.18E-52 | 0.2794 | -15.207 | -4.24907 | 6479.85 | Fc gamma receptor Ic, pseudogene |
| LINC02528 | 1.18E-23 | 8.83E-26 | 0.4056 | -10.4979 | -4.25792 | 59.07 | long intergenic non-protein coding RNA 2528 |
| FCGR1A | 1.87E-33 | 4.07E-36 | 0.3427 | -12.5482 | -4.29967 | 16974.07 | Fc gamma receptor Ia |
| LINC02471 | 3.64E-23 | 2.87E-25 | 0.4241 | -10.3859 | -4.40416 | 35.13 | long intergenic non-protein coding RNA 2471 |
| PDCD1LG2 | 1.17E-27 | 5.42E-30 | 0.3895 | -11.3774 | -4.43168 | 444.02 | programmed cell death 1 ligand 2 |
| LOC105373582 | 1.45E-56 | 7.89E-61 | 0.2696 | -16.4537 | -4.43616 | 382.66 | uncharacterized LOC105373582 |
| CXCL10 | 2.03E-26 | 1.08E-28 | 0.4027 | -11.1131 | -4.47491 | 914.92 | C-X-C motif chemokine ligand 10 |
| SERPING1 | 6.75E-26 | 3.82E-28 | 0.415 | -11 | -4.5649 | 16087.93 | serpin family G member 1 |
| BATF2 | 2.07E-25 | 1.25E-27 | 0.4288 | -10.8925 | -4.67034 | 5093.62 | basic leucine zipper ATF-like transcription factor 2 |
| GBP6 | 2.16E-37 | 2.58E-40 | 0.3525 | -13.2918 | -4.68599 | 761.29 | guanylate binding protein family member 6 |
| ETV7 | 1.20E-29 | 4.49E-32 | 0.4123 | -11.7882 | -4.86051 | 3015.31 | ETS variant transcription factor 7 |
| ACOD1 | 6.77E-32 | 1.77E-34 | 0.425 | -12.2458 | -5.20398 | 48.5 | aconitate decarboxylase 1 |
| GBP1P1 | 4.44E-46 | 2.42E-49 | 0.3549 | -14.7663 | -5.24114 | 969.71 | guanylate binding protein 1 pseudogene 1 |
| IDO1 | 1.01E-38 | 1.10E-41 | 0.4082 | -13.526 | -5.52092 | 10135.21 | indoleamine 2,3-dioxygenase 1 |
| LIPM | 3.62E-35 | 6.50E-38 | 0.4308 | -12.8717 | -5.54538 | 737.57 | lipase family member M |
| APOL4 | 1.60E-41 | 1.48E-44 | 0.4045 | -14.0038 | -5.66437 | 1103.98 | apolipoprotein L4 |
| ANKRD22 | 4.16E-39 | 4.31E-42 | 0.4184 | -13.5947 | -5.68816 | 4826 | ankyrin repeat domain 22 |
