## Supplementary material for "SYSTEMS AND NETWORK BIOLOGY ANALYSIS COMBINED WITH MACHINE LEARNING IDENTIFIES KEY IMMUNE RESPONSE PROFILES AND POTENTIAL CORRELATES OF PROTECTION FOR THE M72/AS01E TUBERCULOSIS VACCINE": https://drive.google.com/file/d/1eW6qRfTMBVGazXTT9-xPsO9vCHcWIqCW/view?usp=drive_link: Table S2 Differential Gene Expression Analysis (D0 VS D37).docx

| Symbol | padj | pvalue | lfcSE | stat | log2FoldChange | baseMean | Gene Name |
| --- | --- | --- | --- | --- | --- | --- | --- |
| IGHV1-69D | 1.81E-03 | 2.27E-05 | 0.3549 | -4.23673 | -1.50373 | 610.61 | immunoglobulin heavy variable 1-69D |
| MYBL2 | 2.48E-04 | 2.41E-06 | 0.3193 | -4.71573 | -1.50594 | 596.94 | MYB proto-oncogene like 2 |
| IGKV3-20 | 1.26E-06 | 6.87E-09 | 0.2622 | -5.79403 | -1.51932 | 5365.47 | immunoglobulin kappa variable 3-20 |
| IGLV1-36 | 3.57E-04 | 3.65E-06 | 0.3338 | -4.63017 | -1.54556 | 320.73 | immunoglobulin lambda variable 1-36 |
| IGHV3OR16-17 | 1.28E-03 | 1.51E-05 | 0.3581 | -4.32725 | -1.54973 | 22.45 | immunoglobulin heavy variable 3/OR16-17 (non-functional) |
| IGLV3-9 | 1.64E-02 | 2.69E-04 | 0.4255 | -3.6433 | -1.55006 | 142.19 | immunoglobulin lambda variable 3-9 |
| IGKV1-8 | 4.16E-06 | 2.49E-08 | 0.279 | -5.57395 | -1.55497 | 343.3 | immunoglobulin kappa variable 1-8 |
| LOC112268316 | 3.89E-05 | 3.08E-07 | 0.3048 | -5.11857 | -1.56015 | 12.19 | immunoglobulin heavy variable 3-23-like |
| IGHV4-28 | 1.21E-04 | 1.08E-06 | 0.3214 | -4.87607 | -1.56717 | 239.1 | immunoglobulin heavy variable 4-28 |
| IGKV3D-7 | 7.10E-07 | 3.44E-09 | 0.2654 | -5.90898 | -1.56815 | 341.96 | immunoglobulin kappa variable 3D-7 |
| IGLV2-34 | 3.32E-06 | 1.95E-08 | 0.2794 | -5.6164 | -1.56946 | 225.08 | immunoglobulin lambda variable 2-34 (pseudogene) |
| IGKV1D-8 | 1.56E-03 | 1.87E-05 | 0.3677 | -4.27954 | -1.57351 | 351.3 | immunoglobulin kappa variable 1D-8 |
| IGHG2 | 1.50E-02 | 2.41E-04 | 0.4288 | -3.672 | -1.57438 | 13115.8 | immunoglobulin heavy constant gamma 2 (G2m marker) |
| IGLV1-51 | 6.16E-06 | 3.82E-08 | 0.29 | -5.49917 | -1.59495 | 1167.93 | immunoglobulin lambda variable 1-51 |
| IGKV3D-11 | 7.50E-07 | 3.76E-09 | 0.2734 | -5.89421 | -1.61151 | 991.97 | immunoglobulin kappa variable 3D-11 |
| IGHV4-55 | 9.23E-05 | 7.96E-07 | 0.3297 | -4.93643 | -1.62744 | 226.55 | immunoglobulin heavy variable 4-55 (pseudogene) |
| IGLJ7 | 8.07E-03 | 1.21E-04 | 0.4252 | -3.8434 | -1.63409 | 18.45 | immunoglobulin lambda joining 7 |
| IGKV1D-16 | 9.25E-05 | 8.08E-07 | 0.3318 | -4.93353 | -1.63685 | 252.53 | immunoglobulin kappa variable 1D-16 |
| LOC102723407 | 2.69E-02 | 4.65E-04 | 0.4735 | -3.50033 | -1.65735 | 865.21 | immunoglobulin heavy variable 4-38-2-like |
| IGHV3-7 | 8.57E-04 | 9.66E-06 | 0.3759 | -4.42469 | -1.66317 | 1551.31 | immunoglobulin heavy variable 3-7 |
| TNFRSF17 | 4.24E-04 | 4.46E-06 | 0.3683 | -4.58867 | -1.69019 | 601.04 | TNF receptor superfamily member 17 |
| IGKV1-6 | 2.60E-04 | 2.54E-06 | 0.369 | -4.70457 | -1.73593 | 689.19 | immunoglobulin kappa variable 1-6 |
| IGHV1-12 | 1.26E-05 | 8.35E-08 | 0.3242 | -5.35942 | -1.73779 | 24.57 | immunoglobulin heavy variable 1-12 (pseudogene) |
| IGKV3-7 | 4.65E-04 | 4.99E-06 | 0.3814 | -4.5651 | -1.74093 | 303.99 | immunoglobulin kappa variable 3-7 (non-functional) |
| IGHV1-69 | 4.66E-04 | 5.02E-06 | 0.3834 | -4.564 | -1.74968 | 488.02 | immunoglobulin heavy variable 1-69 |
| LINC02576 | 3.31E-03 | 4.39E-05 | 0.4295 | -4.08569 | -1.75484 | 25.5 | long intergenic non-protein coding RNA 2576 |
| IGLV3-16 | 1.45E-06 | 7.97E-09 | 0.305 | -5.7692 | -1.75934 | 120.38 | immunoglobulin lambda variable 3-16 |
| IGHV3-41 | 1.84E-05 | 1.26E-07 | 0.3362 | -5.28431 | -1.77634 | 100.04 | immunoglobulin heavy variable 3-41 (pseudogene) |
| BLOC1S5-TXNDC5 | 1.19E-07 | 5.11E-10 | 0.2934 | -6.21557 | -1.82337 | 7037.85 | BLOC1S5-TXNDC5 readthrough (NMD candidate) |
| IGLC6 | 3.13E-05 | 2.41E-07 | 0.3546 | -5.16449 | -1.83154 | 1757.47 | immunoglobulin lambda constant 6 |
| MIXL1 | 1.02E-03 | 1.18E-05 | 0.4198 | -4.38114 | -1.83923 | 38.72 | Mix paired-like homeobox |
| TXNDC5 | 1.11E-07 | 4.71E-10 | 0.2966 | -6.22843 | -1.84765 | 6977.33 | thioredoxin domain containing 5 |
| IGHJ2 | 9.42E-07 | 4.99E-09 | 0.3161 | -5.84751 | -1.84816 | 203.33 | immunoglobulin heavy joining 2 |
| IGKV1-17 | 1.41E-04 | 1.29E-06 | 0.3825 | -4.84164 | -1.85192 | 1046.46 | immunoglobulin kappa variable 1-17 |
| IGKV1D-43 | 3.66E-04 | 3.81E-06 | 0.401 | -4.62148 | -1.85304 | 60.52 | immunoglobulin kappa variable 1D-43 |
| IGKV1-16 | 5.40E-07 | 2.56E-09 | 0.3113 | -5.95758 | -1.85455 | 922.36 | immunoglobulin kappa variable 1-16 |
| IGKV5-2 | 3.31E-07 | 1.55E-09 | 0.3078 | -6.03875 | -1.85876 | 101.28 | immunoglobulin kappa variable 5-2 |
| IGLC2 | 5.47E-06 | 3.33E-08 | 0.3383 | -5.52302 | -1.86869 | 18400.12 | immunoglobulin lambda constant 2 |
| IGLC7 | 2.26E-05 | 1.63E-07 | 0.357 | -5.2375 | -1.86957 | 688.99 | immunoglobulin lambda constant 7 |
| IGKJ4 | 3.53E-08 | 1.39E-10 | 0.2917 | -6.41694 | -1.87213 | 2075.73 | immunoglobulin kappa joining 4 |
| IGHV4-39 | 8.30E-05 | 7.07E-07 | 0.3803 | -4.9594 | -1.8863 | 1345.23 | immunoglobulin heavy variable 4-39 |
| IGLV1-44 | 3.70E-05 | 2.91E-07 | 0.3685 | -5.12892 | -1.89013 | 1820.93 | immunoglobulin lambda variable 1-44 |
| IGLC3 | 1.69E-07 | 7.58E-10 | 0.3073 | -6.15348 | -1.89109 | 14430.63 | immunoglobulin lambda constant 3 (Kern-Oz+ marker) |
| IGKV3-11 | 5.87E-08 | 2.44E-10 | 0.2995 | -6.33099 | -1.89604 | 4383.91 | immunoglobulin kappa variable 3-11 |
| IGHV3-48 | 7.94E-04 | 8.86E-06 | 0.4293 | -4.44319 | -1.90726 | 836.74 | immunoglobulin heavy variable 3-48 |
| GLDC | 6.24E-03 | 9.01E-05 | 0.4875 | -3.9157 | -1.90907 | 206.33 | glycine decarboxylase |
| IGHV2-5 | 2.26E-05 | 1.64E-07 | 0.3673 | -5.23565 | -1.92328 | 791.77 | immunoglobulin heavy variable 2-5 |
| IGHV1OR15-3 | 2.92E-05 | 2.21E-07 | 0.374 | -5.18106 | -1.93764 | 70.3 | immunoglobulin heavy variable 1/OR15-3 (pseudogene) |
| IGKV3D-15 | 3.64E-11 | 5.49E-14 | 0.2579 | -7.51973 | -1.93969 | 1664.05 | immunoglobulin kappa variable 3D-15 |
| IGLV1-41 | 2.66E-04 | 2.66E-06 | 0.4136 | -4.6958 | -1.94209 | 288.32 | immunoglobulin lambda variable 1-41 (pseudogene) |
| IGKC | 9.80E-05 | 8.61E-07 | 0.395 | -4.92091 | -1.94395 | 88344.95 | immunoglobulin kappa constant |
| LOC642131 | 2.81E-05 | 2.09E-07 | 0.3747 | -5.19145 | -1.9453 | 74.28 | immunoglobulin IGHV1OR15-3-like pseudogene |
| IGLV6-57 | 1.49E-04 | 1.39E-06 | 0.4059 | -4.82611 | -1.95893 | 745.35 | immunoglobulin lambda variable 6-57 |
| IGLC5 | 5.59E-06 | 3.44E-08 | 0.3596 | -5.51754 | -1.98428 | 1454.63 | immunoglobulin lambda constant 5 (pseudogene) |
| IGKJ2 | 4.99E-09 | 1.56E-11 | 0.2954 | -6.74233 | -1.99186 | 1951.74 | immunoglobulin kappa joining 2 |
| IGLL5 | 7.62E-05 | 6.33E-07 | 0.4024 | -4.98092 | -2.00427 | 8940.95 | immunoglobulin lambda like polypeptide 5 |
| IGLC1 | 9.25E-05 | 8.05E-07 | 0.407 | -4.93411 | -2.00811 | 8103.91 | immunoglobulin lambda constant 1 |
| IGHJ5 | 6.04E-10 | 1.47E-12 | 0.2863 | -7.0774 | -2.0263 | 1247.13 | immunoglobulin heavy joining 5 |
| IGLV2-11 | 3.21E-06 | 1.87E-08 | 0.3622 | -5.62351 | -2.03709 | 2123.76 | immunoglobulin lambda variable 2-11 |
| FER1L4 | 6.78E-06 | 4.31E-08 | 0.3735 | -5.4775 | -2.04587 | 17.7 | fer-1 like family member 4 (pseudogene) |
| IGKJ3 | 3.31E-09 | 9.28E-12 | 0.3023 | -6.81729 | -2.06074 | 1155.08 | immunoglobulin kappa joining 3 |
| IGHJ3 | 3.94E-08 | 1.57E-10 | 0.3251 | -6.39836 | -2.07996 | 597.6 | immunoglobulin heavy joining 3 |
| IGHV4-59 | 9.10E-07 | 4.74E-09 | 0.3563 | -5.85595 | -2.0863 | 1537.85 | immunoglobulin heavy variable 4-59 |
| IGKV1-9 | 1.23E-04 | 1.10E-06 | 0.4305 | -4.87232 | -2.09748 | 1603.2 | immunoglobulin kappa variable 1-9 |
| IGLV4-60 | 4.43E-03 | 6.12E-05 | 0.5247 | -4.00824 | -2.10304 | 243.75 | immunoglobulin lambda variable 4-60 |
| IGHV4-61 | 7.52E-10 | 1.95E-12 | 0.2993 | -7.03832 | -2.10667 | 830.43 | immunoglobulin heavy variable 4-61 |
| IGLV7-46 | 8.92E-11 | 1.54E-13 | 0.2864 | -7.38375 | -2.11494 | 674.77 | immunoglobulin lambda variable 7-46 |
| IGKJ1 | 2.07E-10 | 3.69E-13 | 0.2925 | -7.2666 | -2.12512 | 2858.29 | immunoglobulin kappa joining 1 |
| TSHR | 6.05E-05 | 4.96E-07 | 0.4248 | -5.02796 | -2.13568 | 37.6 | thyroid stimulating hormone receptor |
| IGLV1-47 | 9.10E-07 | 4.76E-09 | 0.365 | -5.85539 | -2.13706 | 1624.42 | immunoglobulin lambda variable 1-47 |
| IGKV3-15 | 1.14E-13 | 9.25E-17 | 0.2571 | -8.31409 | -2.13722 | 3727.24 | immunoglobulin kappa variable 3-15 |
| IGHJ6 | 5.90E-10 | 1.32E-12 | 0.3047 | -7.09255 | -2.1608 | 924.63 | immunoglobulin heavy joining 6 |
| IGKV1D-37 | 2.81E-05 | 2.09E-07 | 0.4181 | -5.19153 | -2.17073 | 195.63 | immunoglobulin kappa variable 1D-37 (non-functional) |
| IGKV1-37 | 3.76E-06 | 2.23E-08 | 0.3889 | -5.59314 | -2.17535 | 282.14 | immunoglobulin kappa variable 1-37 (non-functional) |
| LOC107983983 | 2.78E-08 | 1.06E-10 | 0.337 | -6.45775 | -2.17647 | 110.67 | uncharacterized LOC107983983 |
| IGLJ2 | 3.12E-10 | 6.05E-13 | 0.303 | -7.19939 | -2.18117 | 1811.8 | immunoglobulin lambda joining 2 |
| IGLV4-69 | 6.60E-06 | 4.16E-08 | 0.3978 | -5.48384 | -2.18155 | 820.8 | immunoglobulin lambda variable 4-69 |
| IGKJ5 | 5.97E-10 | 1.41E-12 | 0.3083 | -7.08323 | -2.18388 | 1271.63 | immunoglobulin kappa joining 5 |
| IGHJ1 | 1.75E-08 | 6.31E-11 | 0.3344 | -6.53621 | -2.18539 | 388.31 | immunoglobulin heavy joining 1 |
| IGKV1D-27 | 1.70E-06 | 9.43E-09 | 0.3833 | -5.74073 | -2.20052 | 301.53 | immunoglobulin kappa variable 1D-27 (pseudogene) |
| IGLV2-23 | 2.26E-05 | 1.64E-07 | 0.4216 | -5.23608 | -2.20757 | 3622.98 | immunoglobulin lambda variable 2-23 |
| KCNN3 | 2.45E-08 | 9.11E-11 | 0.3422 | -6.48104 | -2.21791 | 47.76 | potassium calcium-activated channel subfamily N member 3 |
| IGHV1-3 | 1.27E-02 | 1.99E-04 | 0.5969 | -3.72046 | -2.22082 | 425.02 | immunoglobulin heavy variable 1-3 |
| IGHV4-30-2 | 9.42E-07 | 5.05E-09 | 0.3834 | -5.8455 | -2.24103 | 388.51 | immunoglobulin heavy variable 4-30-2 |
| IGLV1-50 | 1.69E-05 | 1.15E-07 | 0.4263 | -5.30141 | -2.26021 | 345.7 | immunoglobulin lambda variable 1-50 (non-functional) |
| IGLJ3 | 7.50E-08 | 3.15E-10 | 0.3622 | -6.29103 | -2.2787 | 940.57 | immunoglobulin lambda joining 3 |
| IGKV1-13 | 5.58E-08 | 2.29E-10 | 0.3609 | -6.34082 | -2.28849 | 588.7 | immunoglobulin kappa variable 1-13 |
| LOC102724971 | 4.75E-10 | 9.47E-13 | 0.3234 | -7.13804 | -2.30838 | 154.45 | putative V-set and immunoglobulin domain-containing-like protein IGHV4OR15-8 |
| MZB1 | 2.70E-08 | 1.02E-10 | 0.3573 | -6.4641 | -2.30977 | 2048.56 | marginal zone B and B1 cell specific protein |
| IGKV1D-39 | 2.25E-11 | 3.04E-14 | 0.3089 | -7.59668 | -2.34626 | 2631.83 | immunoglobulin kappa variable 1D-39 |
| IGKV1D-13 | 2.15E-06 | 1.23E-08 | 0.414 | -5.69552 | -2.358 | 499.27 | immunoglobulin kappa variable 1D-13 |
| IGKV1-27 | 5.05E-09 | 1.60E-11 | 0.35 | -6.73809 | -2.35842 | 771.7 | immunoglobulin kappa variable 1-27 |
| IGHV3-71 | 8.21E-10 | 2.17E-12 | 0.3401 | -7.02314 | -2.38872 | 81.14 | immunoglobulin heavy variable 3-71 (pseudogene) |
| IGLJ1 | 6.10E-07 | 2.93E-09 | 0.4026 | -5.93566 | -2.38984 | 949.97 | immunoglobulin lambda joining 1 |
| IGKV1-39 | 4.33E-12 | 4.79E-15 | 0.3052 | -7.83236 | -2.39072 | 3227.27 | immunoglobulin kappa variable 1-39 |
| IGHV6-1 | 2.33E-07 | 1.05E-09 | 0.3923 | -6.10088 | -2.39365 | 369.22 | immunoglobulin heavy variable 6-1 |
| IGHV7-56 | 3.31E-07 | 1.55E-09 | 0.3971 | -6.03946 | -2.39832 | 75.42 | immunoglobulin heavy variable 7-56 (pseudogene) |
| IGHJ4 | 5.74E-15 | 3.41E-18 | 0.2762 | -8.69705 | -2.40205 | 1520.08 | immunoglobulin heavy joining 4 |
| IGHV1-46 | 2.95E-07 | 1.35E-09 | 0.3988 | -6.06134 | -2.41723 | 532.79 | immunoglobulin heavy variable 1-46 |
| IGHV3-64 | 2.08E-06 | 1.17E-08 | 0.4241 | -5.70476 | -2.41934 | 199.09 | immunoglobulin heavy variable 3-64 |
| IGLV2-14 | 1.30E-08 | 4.40E-11 | 0.3679 | -6.58993 | -2.42453 | 5288.88 | immunoglobulin lambda variable 2-14 |
| IGHV3-38 | 1.36E-08 | 4.78E-11 | 0.3692 | -6.57778 | -2.4283 | 55.48 | immunoglobulin heavy variable 3-38 (non-functional) |
| IGHV3-69-1 | 5.37E-11 | 8.97E-14 | 0.326 | -7.45527 | -2.43025 | 404.91 | immunoglobulin heavy variable 3-69-1 (pseudogene) |
| IGLV1-40 | 9.46E-09 | 3.16E-11 | 0.3699 | -6.63885 | -2.45596 | 2787.61 | immunoglobulin lambda variable 1-40 |
| IGLV5-48 | 6.48E-06 | 4.05E-08 | 0.4513 | -5.4886 | -2.477 | 15.54 | immunoglobulin lambda variable 5-48 (non-functional) |
| CAV1 | 4.45E-09 | 1.32E-11 | 0.3661 | -6.76639 | -2.47749 | 115.4 | caveolin 1 |
| IGHV3-21 | 3.29E-13 | 3.01E-16 | 0.3037 | -8.17275 | -2.48204 | 1658.36 | immunoglobulin heavy variable 3-21 |
| IGKV2D-40 | 5.01E-11 | 7.83E-14 | 0.3336 | -7.47322 | -2.4932 | 135.61 | immunoglobulin kappa variable 2D-40 |
| IGKV2-40 | 2.84E-14 | 1.83E-17 | 0.2936 | -8.50384 | -2.49631 | 93.19 | immunoglobulin kappa variable 2-40 |
| IGKV2D-26 | 9.10E-04 | 1.04E-05 | 0.5704 | -4.40965 | -2.51505 | 83.79 | immunoglobulin kappa variable 2D-26 |
| IGKV2-26 | 4.26E-03 | 5.84E-05 | 0.6302 | -4.0192 | -2.53301 | 170.68 | immunoglobulin kappa variable 2-26 (pseudogene) |
| LOC107984634 | 2.85E-09 | 7.85E-12 | 0.3735 | -6.84129 | -2.55509 | 100.08 | uncharacterized LOC107984634 |
| IGHV3-20 | 2.14E-05 | 1.51E-07 | 0.4875 | -5.25083 | -2.56003 | 637.51 | immunoglobulin heavy variable 3-20 |
| IGHV3OR16-8 | 5.77E-10 | 1.24E-12 | 0.3608 | -7.10032 | -2.56151 | 69.42 | immunoglobulin heavy variable 3/OR16-8 (non-functional) |
| IGKV2-28 | 9.37E-09 | 3.03E-11 | 0.3855 | -6.64506 | -2.56152 | 2020.55 | immunoglobulin kappa variable 2-28 |
| IGLV3-25 | 6.35E-14 | 4.79E-17 | 0.3078 | -8.39168 | -2.58323 | 1782.89 | immunoglobulin lambda variable 3-25 |
| IGKV2D-28 | 1.44E-08 | 5.11E-11 | 0.3945 | -6.56761 | -2.5912 | 1691.41 | immunoglobulin kappa variable 2D-28 |
| LOC102725101 | 1.99E-08 | 7.29E-11 | 0.3979 | -6.51456 | -2.5922 | 39.98 | immunoglobulin heavy variable 3-23-like |
| IGKV1-5 | 3.31E-09 | 9.45E-12 | 0.3807 | -6.81463 | -2.59446 | 6386.43 | immunoglobulin kappa variable 1-5 |
| IGLV5-45 | 1.35E-08 | 4.65E-11 | 0.3947 | -6.5817 | -2.59762 | 260.79 | immunoglobulin lambda variable 5-45 |
| IGHV3-11 | 2.60E-10 | 4.86E-13 | 0.3605 | -7.22913 | -2.6063 | 1128.73 | immunoglobulin heavy variable 3-11 |
| IGKV4-1 | 9.80E-10 | 2.64E-12 | 0.3739 | -6.99554 | -2.61537 | 6975.56 | immunoglobulin kappa variable 4-1 |
| IGHV3-23 | 2.07E-19 | 6.70E-23 | 0.2663 | -9.85228 | -2.62374 | 4122.19 | immunoglobulin heavy variable 3-23 |
| IGHV3-74 | 5.32E-10 | 1.09E-12 | 0.3693 | -7.11859 | -2.62924 | 1061.63 | immunoglobulin heavy variable 3-74 |
| IGLV3-27 | 5.37E-05 | 4.34E-07 | 0.5267 | -5.05345 | -2.66159 | 263.23 | immunoglobulin lambda variable 3-27 |
| IGKV1-12 | 3.89E-09 | 1.13E-11 | 0.3921 | -6.78859 | -2.66176 | 1599.18 | immunoglobulin kappa variable 1-12 |
| IGKV2D-29 | 5.18E-04 | 5.66E-06 | 0.5882 | -4.53859 | -2.66981 | 669.18 | immunoglobulin kappa variable 2D-29 |
| UCHL1 | 7.08E-06 | 4.54E-08 | 0.4888 | -5.46834 | -2.67314 | 22.91 | ubiquitin C-terminal hydrolase L1 |
| IGKV2-29 | 4.06E-03 | 5.51E-05 | 0.6765 | -4.03291 | -2.72826 | 826.76 | immunoglobulin kappa variable 2-29 |
| IGHV3-22 | 7.11E-18 | 2.68E-21 | 0.288 | -9.47437 | -2.72849 | 263.87 | immunoglobulin heavy variable 3-22 (pseudogene) |
| BHLHA15 | 3.70E-05 | 2.91E-07 | 0.533 | -5.12943 | -2.73391 | 34.84 | basic helix-loop-helix family member a15 |
| IGHV3-15 | 2.60E-10 | 4.90E-13 | 0.3853 | -7.22807 | -2.78514 | 1443.03 | immunoglobulin heavy variable 3-15 |
| IGHV1-2 | 3.13E-08 | 1.21E-10 | 0.4355 | -6.43762 | -2.80368 | 1584.87 | immunoglobulin heavy variable 1-2 |
| IGHV3-53 | 3.22E-11 | 4.69E-14 | 0.372 | -7.54036 | -2.80479 | 678.6 | immunoglobulin heavy variable 3-53 |
| IGHV1-18 | 5.97E-10 | 1.42E-12 | 0.3961 | -7.08254 | -2.80505 | 1244.82 | immunoglobulin heavy variable 1-18 |
| IGKV1D-12 | 5.90E-10 | 1.34E-12 | 0.3956 | -7.09052 | -2.80535 | 696.34 | immunoglobulin kappa variable 1D-12 |
| IGHV3-49 | 4.99E-09 | 1.56E-11 | 0.4181 | -6.74257 | -2.8192 | 569.83 | immunoglobulin heavy variable 3-49 |
| IGHV7-4-1 | 4.98E-02 | 9.42E-04 | 0.8548 | -3.30744 | -2.82722 | 415.92 | immunoglobulin heavy variable 7-4-1 |
| IGHV3-73 | 4.80E-09 | 1.45E-11 | 0.4269 | -6.75296 | -2.88306 | 437.42 | immunoglobulin heavy variable 3-73 |
| IGHV3-13 | 4.03E-12 | 4.13E-15 | 0.3681 | -7.85091 | -2.89016 | 431.28 | immunoglobulin heavy variable 3-13 |
| IGLV5-37 | 7.71E-05 | 6.49E-07 | 0.5811 | -4.97615 | -2.89141 | 112.24 | immunoglobulin lambda variable 5-37 |
| ELK2AP | 5.34E-08 | 2.16E-10 | 0.4563 | -6.34951 | -2.89744 | 597.73 | ETS transcription factor ELK2A, pseudogene |
| IGHGP | 4.33E-12 | 4.90E-15 | 0.3768 | -7.82941 | -2.95012 | 4278.57 | immunoglobulin heavy constant gamma P (non-functional) |
| IGHV3-35 | 9.33E-12 | 1.21E-14 | 0.3855 | -7.71525 | -2.97386 | 80.48 | immunoglobulin heavy variable 3-35 (non-functional) |
| IGHV3-16 | 1.06E-17 | 5.16E-21 | 0.3167 | -9.40587 | -2.97862 | 217.65 | immunoglobulin heavy variable 3-16 (non-functional) |
| IGHV3-62 | 2.83E-11 | 3.96E-14 | 0.3942 | -7.56222 | -2.98069 | 105.37 | immunoglobulin heavy variable 3-62 (pseudogene) |
| IGKV2D-30 | 1.09E-23 | 1.77E-27 | 0.2812 | -10.8611 | -3.05438 | 1085.38 | immunoglobulin kappa variable 2D-30 |
| IGHG4 | 5.77E-10 | 1.22E-12 | 0.4302 | -7.10254 | -3.05554 | 5617.83 | immunoglobulin heavy constant gamma 4 (G4m marker) |
| IGHV3-65 | 9.64E-18 | 4.16E-21 | 0.3258 | -9.42859 | -3.07166 | 108.59 | immunoglobulin heavy variable 3-65 (pseudogene) |
| IGKV2-30 | 1.93E-25 | 2.08E-29 | 0.2745 | -11.2593 | -3.09116 | 1977.62 | immunoglobulin kappa variable 2-30 |
| IGHV3-66 | 5.36E-13 | 5.20E-16 | 0.3948 | -8.10662 | -3.20061 | 697.27 | immunoglobulin heavy variable 3-66 |
| IGKV2D-24 | 6.04E-10 | 1.50E-12 | 0.4583 | -7.07463 | -3.24249 | 246.9 | immunoglobulin kappa variable 2D-24 (non-functional) |
| IGKV2-24 | 7.06E-12 | 8.76E-15 | 0.4182 | -7.75608 | -3.24368 | 990.74 | immunoglobulin kappa variable 2-24 |
| IGLV4-3 | 9.76E-06 | 6.36E-08 | 0.6058 | -5.40825 | -3.27646 | 94.85 | immunoglobulin lambda variable 4-3 |
| IGHV3-19 | 5.34E-11 | 8.63E-14 | 0.4415 | -7.46035 | -3.29389 | 35.17 | immunoglobulin heavy variable 3-19 (pseudogene) |
| IGHV4-4 | 1.37E-07 | 5.97E-10 | 0.5567 | -6.19111 | -3.44644 | 871.47 | immunoglobulin heavy variable 4-4 |
| IGHG3 | 1.53E-07 | 6.78E-10 | 0.5594 | -6.17122 | -3.45223 | 26297.37 | immunoglobulin heavy constant gamma 3 (G3m marker) |
| IGHV3-64D | 7.47E-07 | 3.71E-09 | 0.5885 | -5.89681 | -3.47005 | 508.77 | immunoglobulin heavy variable 3-64D |
| IGLV7-43 | 4.47E-12 | 5.30E-15 | 0.4551 | -7.81955 | -3.55846 | 1168.41 | immunoglobulin lambda variable 7-43 |
| IGHV3-33 | 2.02E-20 | 5.44E-24 | 0.3634 | -10.1015 | -3.67067 | 3923.05 | immunoglobulin heavy variable 3-33 |
| LOC102724977 | 1.59E-16 | 8.59E-20 | 0.41 | -9.10543 | -3.73305 | 208.2 | uncharacterized LOC102724977 |
| IGHG1 | 7.12E-10 | 1.80E-12 | 0.5342 | -7.04887 | -3.76539 | 66748.11 | immunoglobulin heavy constant gamma 1 (G1m marker) |
| IGHV3-30 | 5.28E-21 | 1.14E-24 | 0.3809 | -10.2537 | -3.90614 | 5622.77 | immunoglobulin heavy variable 3-30 |
| SDC1 | 3.97E-14 | 2.78E-17 | 0.4794 | -8.45541 | -4.05353 | 99.69 | syndecan 1 |
| IGLV3-10 | 2.92E-13 | 2.52E-16 | 0.5337 | -8.19426 | -4.37317 | 1574.97 | immunoglobulin lambda variable 3-10 |
| IGLV9-49 | 1.71E-25 | 9.21E-30 | 0.468 | -11.331 | -5.30294 | 1690.63 | immunoglobulin lambda variable 9-49 |
