## Supplementary material for "SYSTEMS AND NETWORK BIOLOGY ANALYSIS COMBINED WITH MACHINE LEARNING IDENTIFIES KEY IMMUNE RESPONSE PROFILES AND POTENTIAL CORRELATES OF PROTECTION FOR THE M72/AS01E TUBERCULOSIS VACCINE": https://drive.google.com/file/d/1eW6qRfTMBVGazXTT9-xPsO9vCHcWIqCW/view?usp=drive_link: Table S3 Differential Gene Expression Analysis (D31 VS D37).docx

| Symbol | padj | pvalue | lfcSE | stat | log2FoldChange | baseMean | Gene Name |
| --- | --- | --- | --- | --- | --- | --- | --- |
| IGLV9-49 | 1.90E-28 | 1.17E-30 | 0.4632 | 11.51009 | 5.33197 | 1520.53 | immunoglobulin lambda variable 9-49 |
| IGHG1 | 1.78E-16 | 4.62E-18 | 0.4754 | 8.662493 | 4.118134 | 58896.2 | immunoglobulin heavy constant gamma 1 (G1m marker) |
| IGLV3-10 | 1.53E-10 | 1.05E-11 | 0.6011 | 6.800162 | 4.087374 | 1436.06 | immunoglobulin lambda variable 3-10 |
| SDC1 | 6.90E-19 | 1.30E-20 | 0.4369 | 9.308542 | 4.066722 | 89.46 | syndecan 1 |
| IGHG3 | 1.05E-13 | 3.96E-15 | 0.5124 | 7.856106 | 4.025271 | 22826.16 | immunoglobulin heavy constant gamma 3 (G3m marker) |
| IGLV4-3 | 3.02E-08 | 3.21E-09 | 0.6197 | 5.920689 | 3.669136 | 82.44 | immunoglobulin lambda variable 4-3 |
| IGHV3-64D | 1.09E-08 | 1.06E-09 | 0.6003 | 6.100396 | 3.662185 | 451.41 | immunoglobulin heavy variable 3-64D |
| LOC102724977 | 3.89E-14 | 1.36E-15 | 0.4361 | 7.98864 | 3.484042 | 188.9 | uncharacterized LOC102724977 |
| ELK2AP | 3.57E-15 | 1.08E-16 | 0.4115 | 8.295223 | 3.413588 | 512.33 | ETS transcription factor ELK2A, pseudogene |
| IGHV3-30 | 4.25E-12 | 2.16E-13 | 0.4645 | 7.338704 | 3.408651 | 5186.88 | immunoglobulin heavy variable 3-30 |
| IGHGP | 8.61E-25 | 7.52E-27 | 0.3162 | 10.72799 | 3.392011 | 3698.74 | immunoglobulin heavy constant gamma P (non-functional) |
| IGHG4 | 4.77E-17 | 1.16E-18 | 0.3836 | 8.818771 | 3.383175 | 4891.37 | immunoglobulin heavy constant gamma 4 (G4m marker) |
| IGLC4 | 4.61E-04 | 1.35E-04 | 0.8588 | 3.817045 | 3.278023 | 100.36 | immunoglobulin lambda constant 4 (pseudogene) |
| IGHV3-33 | 2.23E-13 | 8.89E-15 | 0.4219 | 7.754245 | 3.271161 | 3602.49 | immunoglobulin heavy variable 3-33 |
| IGHV4-4 | 3.64E-07 | 4.88E-08 | 0.5939 | 5.455713 | 3.240253 | 797.1 | immunoglobulin heavy variable 4-4 |
| IGLV7-43 | 6.23E-10 | 4.77E-11 | 0.4878 | 6.577864 | 3.208859 | 1068.83 | immunoglobulin lambda variable 7-43 |
| IGHV3-66 | 1.38E-13 | 5.33E-15 | 0.4021 | 7.818808 | 3.144026 | 625.07 | immunoglobulin heavy variable 3-66 |
| IGKV2-29 | 1.41E-05 | 2.72E-06 | 0.661 | 4.690769 | 3.100467 | 708.01 | immunoglobulin kappa variable 2-29 |
| IGKV2D-24 | 9.13E-09 | 8.70E-10 | 0.495 | 6.131627 | 3.034923 | 225.34 | immunoglobulin kappa variable 2D-24 (non-functional) |
| IGKV1D-12 | 4.39E-13 | 1.83E-14 | 0.3947 | 7.662314 | 3.024451 | 607.1 | immunoglobulin kappa variable 1D-12 |
| IGKV2-24 | 6.35E-11 | 4.04E-12 | 0.4357 | 6.935663 | 3.02187 | 904.55 | immunoglobulin kappa variable 2-24 |
| IGHV3-73 | 1.73E-11 | 9.88E-13 | 0.4173 | 7.132112 | 2.976374 | 385.58 | immunoglobulin heavy variable 3-73 |
| IGHV3-35 | 1.60E-12 | 7.58E-14 | 0.3942 | 7.477351 | 2.94731 | 71.79 | immunoglobulin heavy variable 3-35 (non-functional) |
| IGKV2D-29 | 3.39E-06 | 5.60E-07 | 0.5839 | 5.00463 | 2.922002 | 577.58 | immunoglobulin kappa variable 2D-29 |
| IGHV7-4-1 | 2.12E-03 | 7.56E-04 | 0.8647 | 3.368252 | 2.912443 | 365.91 | immunoglobulin heavy variable 7-4-1 |
| IGKV2-26 | 1.54E-05 | 2.99E-06 | 0.6202 | 4.671234 | 2.897291 | 145.19 | immunoglobulin kappa variable 2-26 (pseudogene) |
| IGKV2D-26 | 4.64E-06 | 7.94E-07 | 0.5837 | 4.936744 | 2.881504 | 71.27 | immunoglobulin kappa variable 2D-26 |
| IGKV1-12 | 2.68E-12 | 1.32E-13 | 0.3821 | 7.404266 | 2.829211 | 1398.23 | immunoglobulin kappa variable 1-12 |
| IGKV1-5 | 8.52E-16 | 2.38E-17 | 0.3324 | 8.473698 | 2.816386 | 5586.05 | immunoglobulin kappa variable 1-5 |
| IGHV3-65 | 5.32E-14 | 1.91E-15 | 0.3544 | 7.947006 | 2.816149 | 98.98 | immunoglobulin heavy variable 3-65 (pseudogene) |
| IGHV3-15 | 2.01E-11 | 1.16E-12 | 0.3942 | 7.109803 | 2.802893 | 1287.21 | immunoglobulin heavy variable 3-15 |
| IGKV2-30 | 2.71E-19 | 4.87E-21 | 0.2969 | 9.411869 | 2.794043 | 1814.88 | immunoglobulin kappa variable 2-30 |
| IGHV3-19 | 4.53E-07 | 6.20E-08 | 0.5146 | 5.412886 | 2.785599 | 32.71 | immunoglobulin heavy variable 3-19 (pseudogene) |
| NUAK1 | 9.66E-07 | 1.42E-07 | 0.5215 | 5.262918 | 2.744379 | 28.07 | NUAK family kinase 1 |
| IGKV2D-30 | 4.81E-19 | 8.85E-21 | 0.2924 | 9.348939 | 2.733816 | 998.04 | immunoglobulin kappa variable 2D-30 |
| IGHV3-64 | 2.22E-11 | 1.30E-12 | 0.3815 | 7.09451 | 2.706649 | 171.79 | immunoglobulin heavy variable 3-64 |
| IGHV1-2 | 3.24E-09 | 2.84E-10 | 0.4291 | 6.307144 | 2.706311 | 1421.73 | immunoglobulin heavy variable 1-2 |
| IGHV1-18 | 5.11E-10 | 3.84E-11 | 0.4094 | 6.610016 | 2.706039 | 1118.62 | immunoglobulin heavy variable 1-18 |
| CAV1 | 3.34E-13 | 1.36E-14 | 0.3504 | 7.699807 | 2.698297 | 99.91 | caveolin 1 |
| IGLV1-50 | 1.32E-14 | 4.32E-16 | 0.3315 | 8.129306 | 2.695055 | 290.99 | immunoglobulin lambda variable 1-50 (non-functional) |
| IGHV3-49 | 2.80E-08 | 2.95E-09 | 0.4527 | 5.934476 | 2.686726 | 516.33 | immunoglobulin heavy variable 3-49 |
| BHLHA15 | 8.48E-06 | 1.56E-06 | 0.5566 | 4.803373 | 2.673327 | 31.1 | basic helix-loop-helix family member a15 |
| IGHV3-62 | 1.53E-09 | 1.27E-10 | 0.4155 | 6.430549 | 2.671955 | 96.44 | immunoglobulin heavy variable 3-62 (pseudogene) |
| DGKK | 2.52E-06 | 4.03E-07 | 0.5215 | 5.067553 | 2.64262 | 70.8 | diacylglycerol kinase kappa |
| IGLV1-40 | 6.32E-16 | 1.74E-17 | 0.31 | 8.509899 | 2.63777 | 2422.56 | immunoglobulin lambda variable 1-40 |
| IGHV3-16 | 2.61E-13 | 1.05E-14 | 0.3387 | 7.733002 | 2.619438 | 200.72 | immunoglobulin heavy variable 3-16 (non-functional) |
| LOC107984634 | 3.22E-09 | 2.82E-10 | 0.4128 | 6.308449 | 2.604052 | 88.62 | uncharacterized LOC107984634 |
| IGKV2D-28 | 2.11E-09 | 1.79E-10 | 0.4077 | 6.378216 | 2.600258 | 1499.29 | immunoglobulin kappa variable 2D-28 |
| IGLJ1 | 8.19E-10 | 6.40E-11 | 0.3931 | 6.534047 | 2.568337 | 822.91 | immunoglobulin lambda joining 1 |
| IGKV2-28 | 3.47E-09 | 3.06E-10 | 0.4036 | 6.295778 | 2.540971 | 1796.74 | immunoglobulin kappa variable 2-28 |
| UGT2B17 | 2.04E-03 | 7.24E-04 | 0.7465 | 3.380333 | 2.523496 | 36.37 | UDP glucuronosyltransferase family 2 member B17 |
| IGKV4-1 | 3.14E-09 | 2.74E-10 | 0.399 | 6.313095 | 2.519073 | 6246.36 | immunoglobulin kappa variable 4-1 |
| LINGO2 | 8.97E-08 | 1.06E-08 | 0.4387 | 5.720521 | 2.509506 | 23.53 | leucine rich repeat and Ig domain containing 2 |
| IGKV1-9 | 2.07E-09 | 1.75E-10 | 0.3927 | 6.381689 | 2.505889 | 1351.87 | immunoglobulin kappa variable 1-9 |
| IGHV3-53 | 1.41E-08 | 1.39E-09 | 0.4136 | 6.05612 | 2.50454 | 623.6 | immunoglobulin heavy variable 3-53 |
| IGHV3-38 | 5.10E-10 | 3.83E-11 | 0.3776 | 6.610617 | 2.496167 | 48.77 | immunoglobulin heavy variable 3-38 (non-functional) |
| IGLV2-23 | 7.23E-09 | 6.71E-10 | 0.4039 | 6.172697 | 2.492963 | 3089.87 | immunoglobulin lambda variable 2-23 |
| IGHV3-11 | 5.06E-10 | 3.79E-11 | 0.3767 | 6.612047 | 2.49103 | 1017.21 | immunoglobulin heavy variable 3-11 |
| IGHV3-20 | 2.43E-06 | 3.86E-07 | 0.4902 | 5.075645 | 2.488334 | 568.08 | immunoglobulin heavy variable 3-20 |
| IGKV1-39 | 1.33E-14 | 4.36E-16 | 0.3047 | 8.12816 | 2.476903 | 2832 | immunoglobulin kappa variable 1-39 |
| CPXM1 | 3.61E-07 | 4.83E-08 | 0.4485 | 5.457452 | 2.447675 | 27.75 | carboxypeptidase X, M14 family member 1 |
| IGKV1D-39 | 7.83E-14 | 2.89E-15 | 0.31 | 7.895625 | 2.447667 | 2302.89 | immunoglobulin kappa variable 1D-39 |
| IGKV1D-13 | 1.37E-07 | 1.70E-08 | 0.4339 | 5.640339 | 2.447319 | 436.9 | immunoglobulin kappa variable 1D-13 |
| IGHV3-13 | 3.06E-08 | 3.25E-09 | 0.4134 | 5.918416 | 2.446681 | 401.29 | immunoglobulin heavy variable 3-13 |
| IGKV1-13 | 6.67E-11 | 4.29E-12 | 0.3503 | 6.927355 | 2.426627 | 512.58 | immunoglobulin kappa variable 1-13 |
| IGLV2-14 | 6.44E-09 | 5.93E-10 | 0.3905 | 6.192169 | 2.417806 | 4686.99 | immunoglobulin lambda variable 2-14 |
| IGLV5-45 | 6.99E-08 | 8.11E-09 | 0.4186 | 5.766057 | 2.413499 | 239.78 | immunoglobulin lambda variable 5-45 |
| IGLV4-60 | 2.40E-05 | 4.91E-06 | 0.5241 | 4.568429 | 2.394368 | 206.72 | immunoglobulin lambda variable 4-60 |
| GLDC | 1.95E-06 | 3.03E-07 | 0.4656 | 5.121361 | 2.384344 | 169.92 | glycine decarboxylase |
| IGHV3-74 | 4.34E-09 | 3.87E-10 | 0.3801 | 6.259122 | 2.37921 | 970.85 | immunoglobulin heavy variable 3-74 |
| LOC102724971 | 4.28E-12 | 2.18E-13 | 0.3242 | 7.337453 | 2.379016 | 135.95 | putative V-set and immunoglobulin domain-containing-like protein IGHV4OR15-8 |
| IGHV4-30-2 | 2.78E-11 | 1.67E-12 | 0.3366 | 7.059749 | 2.376455 | 338.36 | immunoglobulin heavy variable 4-30-2 |
| IGHV3-22 | 6.85E-13 | 3.01E-14 | 0.3127 | 7.597689 | 2.375429 | 243.98 | immunoglobulin heavy variable 3-22 (pseudogene) |
| IGLV3-25 | 6.44E-11 | 4.10E-12 | 0.3412 | 6.933556 | 2.365933 | 1625.35 | immunoglobulin lambda variable 3-25 |
| IGHV3-21 | 2.72E-12 | 1.34E-13 | 0.3187 | 7.401916 | 2.358631 | 1490.29 | immunoglobulin heavy variable 3-21 |
| IGKV1D-27 | 6.29E-10 | 4.83E-11 | 0.3584 | 6.576098 | 2.356867 | 260.58 | immunoglobulin kappa variable 1D-27 (pseudogene) |
| IGLV3-27 | 1.60E-04 | 4.11E-05 | 0.5739 | 4.101422 | 2.35364 | 243 | immunoglobulin lambda variable 3-27 |
| IGHV3-23 | 1.66E-13 | 6.48E-15 | 0.3011 | 7.794278 | 2.347241 | 3788.1 | immunoglobulin heavy variable 3-23 |
| IGHV1-46 | 1.96E-08 | 1.99E-09 | 0.3912 | 5.998554 | 2.34648 | 474.9 | immunoglobulin heavy variable 1-46 |
| TSHR | 2.33E-06 | 3.68E-07 | 0.4586 | 5.084697 | 2.331792 | 32.33 | thyroid stimulating hormone receptor |
| IGLC1 | 5.50E-09 | 5.00E-10 | 0.3729 | 6.21909 | 2.319327 | 6837.83 | immunoglobulin lambda constant 1 |
| LOC102725101 | 3.46E-07 | 4.62E-08 | 0.4236 | 5.465294 | 2.314949 | 36.57 | immunoglobulin heavy variable 3-23-like |
| LOC105378157 | 9.62E-08 | 1.15E-08 | 0.4053 | 5.707695 | 2.313093 | 47.53 | uncharacterized LOC105378157 |
| MZB1 | 4.72E-09 | 4.23E-10 | 0.3692 | 6.245255 | 2.306047 | 1813.42 | marginal zone B and B1 cell specific protein |
| IGKV1D-37 | 1.10E-08 | 1.07E-09 | 0.3774 | 6.098398 | 2.301698 | 169.62 | immunoglobulin kappa variable 1D-37 (non-functional) |
| IGHV7-56 | 1.99E-07 | 2.54E-08 | 0.4131 | 5.570449 | 2.30111 | 67.59 | immunoglobulin heavy variable 7-56 (pseudogene) |
| MIXL1 | 6.64E-08 | 7.64E-09 | 0.3979 | 5.776124 | 2.298213 | 31.85 | Mix paired-like homeobox |
| IGLL5 | 7.59E-09 | 7.12E-10 | 0.3728 | 6.163472 | 2.29759 | 7559.63 | immunoglobulin lambda like polypeptide 5 |
| VIT | 9.73E-06 | 1.81E-06 | 0.4809 | 4.773346 | 2.295521 | 17.75 | vitrin |
| IGKV1-37 | 6.00E-10 | 4.59E-11 | 0.3484 | 6.583777 | 2.293797 | 245.61 | immunoglobulin kappa variable 1-37 (non-functional) |
| IGLV4-69 | 3.18E-08 | 3.39E-09 | 0.3878 | 5.91148 | 2.29227 | 720.96 | immunoglobulin lambda variable 4-69 |
| LOC107986676 | 1.26E-07 | 1.53E-08 | 0.4051 | 5.657917 | 2.29199 | 13.67 | uncharacterized LOC107986676 |
| IGKJ5 | 1.42E-12 | 6.64E-14 | 0.3058 | 7.494741 | 2.29169 | 1109.8 | immunoglobulin kappa joining 5 |
| IGHV6-1 | 1.38E-06 | 2.09E-07 | 0.4399 | 5.191542 | 2.283833 | 332.29 | immunoglobulin heavy variable 6-1 |
| IGHV3-71 | 8.88E-10 | 7.00E-11 | 0.3497 | 6.520647 | 2.280447 | 72.78 | immunoglobulin heavy variable 3-71 (pseudogene) |
| FER1L4 | 2.20E-07 | 2.82E-08 | 0.4085 | 5.552315 | 2.26789 | 15.11 | fer-1 like family member 4 (pseudogene) |
| IGHV3OR16-8 | 7.53E-09 | 7.05E-10 | 0.3663 | 6.16506 | 2.257979 | 63.8 | immunoglobulin heavy variable 3/OR16-8 (non-functional) |
| LINC02576 | 8.53E-07 | 1.23E-07 | 0.4245 | 5.288284 | 2.244763 | 20.76 | long intergenic non-protein coding RNA 2576 |
| IGKV2D-40 | 5.31E-10 | 4.02E-11 | 0.3382 | 6.603188 | 2.233521 | 124.29 | immunoglobulin kappa variable 2D-40 |
| IGHG2 | 3.65E-10 | 2.67E-11 | 0.335 | 6.663909 | 2.232587 | 10364.62 | immunoglobulin heavy constant gamma 2 (G2m marker) |
| IGHV3-69-1 | 4.66E-10 | 3.48E-11 | 0.337 | 6.62485 | 2.232346 | 367.19 | immunoglobulin heavy variable 3-69-1 (pseudogene) |
| IGLJ3 | 2.16E-09 | 1.83E-10 | 0.3493 | 6.374904 | 2.22661 | 837.54 | immunoglobulin lambda joining 3 |
| IGLJ2 | 7.92E-12 | 4.22E-13 | 0.3067 | 7.248164 | 2.223355 | 1592.52 | immunoglobulin lambda joining 2 |
| EFNA5 | 6.56E-08 | 7.53E-09 | 0.3836 | 5.778621 | 2.216764 | 26.74 | ephrin A5 |
| LOC107983983 | 3.94E-10 | 2.90E-11 | 0.3327 | 6.651653 | 2.213204 | 97.68 | uncharacterized LOC107983983 |
| IGHV4-61 | 1.55E-13 | 6.03E-15 | 0.2832 | 7.803357 | 2.210069 | 722.99 | immunoglobulin heavy variable 4-61 |
| SLC1A7 | 5.75E-08 | 6.53E-09 | 0.3806 | 5.802604 | 2.208425 | 43.05 | solute carrier family 1 member 7 |
| IGKV3-15 | 1.43E-15 | 4.09E-17 | 0.2624 | 8.410319 | 2.206704 | 3273.28 | immunoglobulin kappa variable 3-15 |
| IGKV1-27 | 3.43E-07 | 4.58E-08 | 0.4032 | 5.46699 | 2.204461 | 696.23 | immunoglobulin kappa variable 1-27 |
| COL13A1 | 1.59E-04 | 4.10E-05 | 0.5369 | 4.101812 | 2.202376 | 26.83 | collagen type XIII alpha 1 chain |
| LOC105369656 | 6.67E-09 | 6.15E-10 | 0.3545 | 6.186516 | 2.193279 | 29.39 | uncharacterized LOC105369656 |
| IGHV1OR15-3 | 1.36E-07 | 1.67E-08 | 0.3886 | 5.643107 | 2.193126 | 59.58 | immunoglobulin heavy variable 1/OR15-3 (pseudogene) |
| IGLV1-41 | 4.83E-08 | 5.38E-09 | 0.3757 | 5.835091 | 2.192029 | 245.06 | immunoglobulin lambda variable 1-41 (pseudogene) |
| IGKV1-17 | 1.51E-10 | 1.03E-11 | 0.3218 | 6.802897 | 2.189378 | 877.72 | immunoglobulin kappa variable 1-17 |
| KLRC2 | 1.62E-08 | 1.62E-09 | 0.3609 | 6.03173 | 2.1767 | 147.37 | killer cell lectin like receptor C2 |
| IGKJ3 | 2.98E-11 | 1.80E-12 | 0.3087 | 7.049325 | 2.176163 | 1003.63 | immunoglobulin kappa joining 3 |
| LOC642131 | 1.97E-07 | 2.50E-08 | 0.3891 | 5.573333 | 2.168662 | 63.27 | immunoglobulin IGHV1OR15-3-like pseudogene |
| TNFRSF17 | 9.44E-10 | 7.47E-11 | 0.3327 | 6.510885 | 2.166233 | 489.85 | TNF receptor superfamily member 17 |
| IGHV4-59 | 2.82E-10 | 2.02E-11 | 0.3214 | 6.704401 | 2.155032 | 1344.86 | immunoglobulin heavy variable 4-59 |
| IGLC5 | 3.52E-10 | 2.57E-11 | 0.3217 | 6.669595 | 2.145413 | 1252.91 | immunoglobulin lambda constant 5 (pseudogene) |
| IGKJ1 | 1.12E-11 | 6.12E-13 | 0.2973 | 7.197874 | 2.139748 | 2521.97 | immunoglobulin kappa joining 1 |
| IGHJ1 | 2.28E-08 | 2.35E-09 | 0.3566 | 5.971594 | 2.129501 | 346.15 | immunoglobulin heavy joining 1 |
| KLRC3 | 1.14E-09 | 9.22E-11 | 0.3274 | 6.479267 | 2.121085 | 187.8 | killer cell lectin like receptor C3 |
| LIM2 | 7.74E-09 | 7.28E-10 | 0.3436 | 6.159957 | 2.116339 | 18.89 | lens intrinsic membrane protein 2 |
| IGHJ4 | 5.38E-11 | 3.38E-12 | 0.3037 | 6.960789 | 2.114156 | 1399.31 | immunoglobulin heavy joining 4 |
| IGKJ2 | 2.72E-12 | 1.34E-13 | 0.2846 | 7.401873 | 2.106572 | 1694.04 | immunoglobulin kappa joining 2 |
| UCHL1 | 1.23E-04 | 3.06E-05 | 0.5052 | 4.168694 | 2.105981 | 21.81 | ubiquitin C-terminal hydrolase L1 |
| IGKV2-40 | 2.34E-11 | 1.38E-12 | 0.2968 | 7.086191 | 2.102831 | 87.05 | immunoglobulin kappa variable 2-40 |
| PCDH1 | 1.38E-12 | 6.44E-14 | 0.2782 | 7.498806 | 2.085859 | 79.1 | protocadherin 1 |
| KLRF1 | 1.73E-14 | 5.78E-16 | 0.2561 | 8.093833 | 2.072448 | 699.24 | killer cell lectin like receptor F1 |
| IGLC6 | 2.41E-09 | 2.06E-10 | 0.3257 | 6.356569 | 2.070093 | 1489.53 | immunoglobulin lambda constant 6 |
| IGKV1-6 | 6.76E-11 | 4.35E-12 | 0.2986 | 6.925235 | 2.068006 | 576.76 | immunoglobulin kappa variable 1-6 |
| TXNDC5 | 1.00E-11 | 5.47E-13 | 0.2858 | 7.213135 | 2.061163 | 5938.11 | thioredoxin domain containing 5 |
| NMUR1 | 7.11E-22 | 9.64E-24 | 0.2047 | 10.0453 | 2.056129 | 615.37 | neuromedin U receptor 1 |
| IGKC | 4.96E-11 | 3.11E-12 | 0.2947 | 6.972705 | 2.054559 | 76641.49 | immunoglobulin kappa constant |
| IGLV1-44 | 9.35E-10 | 7.38E-11 | 0.3148 | 6.512758 | 2.050044 | 1567.59 | immunoglobulin lambda variable 1-44 |
| XCR1 | 1.66E-12 | 7.87E-14 | 0.2742 | 7.472527 | 2.04869 | 46.76 | X-C motif chemokine receptor 1 |
| IGHJ6 | 8.01E-10 | 6.25E-11 | 0.3129 | 6.537604 | 2.045941 | 830.55 | immunoglobulin heavy joining 6 |
| JCHAIN | 1.55E-06 | 2.37E-07 | 0.3958 | 5.168002 | 2.045648 | 9189.59 | joining chain of multimeric IgA and IgM |
| PODN | 3.78E-06 | 6.31E-07 | 0.4102 | 4.981527 | 2.043469 | 31.59 | podocan |
| IGKV1-16 | 2.47E-09 | 2.12E-10 | 0.3215 | 6.35224 | 2.042238 | 787.22 | immunoglobulin kappa variable 1-16 |
| BLOC1S5-TXNDC5 | 1.13E-11 | 6.20E-13 | 0.2832 | 7.196101 | 2.038258 | 5983.74 | BLOC1S5-TXNDC5 readthrough (NMD candidate) |
| ZMAT4 | 1.50E-03 | 5.11E-04 | 0.5857 | 3.47518 | 2.035576 | 22.72 | zinc finger matrin-type 4 |
| BNC2 | 5.45E-08 | 6.15E-09 | 0.3487 | 5.812657 | 2.027058 | 43.83 | basonuclin 2 |
| IGHJ3 | 1.06E-08 | 1.02E-09 | 0.3311 | 6.105744 | 2.021684 | 531.02 | immunoglobulin heavy joining 3 |
| IGLV2-11 | 2.99E-08 | 3.17E-09 | 0.3411 | 5.92251 | 2.020241 | 1874.42 | immunoglobulin lambda variable 2-11 |
| IGHV4-39 | 2.09E-08 | 2.13E-09 | 0.336 | 5.987205 | 2.0115 | 1159.36 | immunoglobulin heavy variable 4-39 |
| KCNN3 | 2.35E-07 | 3.03E-08 | 0.3629 | 5.53986 | 2.010636 | 43.44 | potassium calcium-activated channel subfamily N member 3 |
| IGLV6-57 | 1.17E-05 | 2.21E-06 | 0.4241 | 4.733071 | 2.007311 | 652.23 | immunoglobulin lambda variable 6-57 |
| IGHV5-10-1 | 4.37E-03 | 1.73E-03 | 0.6404 | 3.13347 | 2.006644 | 199.71 | immunoglobulin heavy variable 5-10-1 |
| IGLV1-47 | 3.47E-08 | 3.75E-09 | 0.3402 | 5.894967 | 2.005545 | 1459.82 | immunoglobulin lambda variable 1-47 |
| IGKV3D-15 | 8.14E-13 | 3.64E-14 | 0.2647 | 7.573294 | 2.00443 | 1456.69 | immunoglobulin kappa variable 3D-15 |
| FGFBP2 | 1.57E-12 | 7.39E-14 | 0.2677 | 7.480734 | 2.002368 | 2474 | fibroblast growth factor binding protein 2 |
| PRSS23 | 3.67E-11 | 2.25E-12 | 0.2852 | 7.018221 | 2.001736 | 493.32 | serine protease 23 |
| ARHGEF28 | 4.72E-06 | 8.09E-07 | 0.4013 | 4.9331 | 1.979409 | 19.14 | Rho guanine nucleotide exchange factor 28 |
| COLGALT2 | 2.02E-14 | 6.80E-16 | 0.2445 | 8.073987 | 1.974042 | 105.87 | collagen beta(1-O)galactosyltransferase 2 |
| IGHV3-41 | 1.38E-09 | 1.14E-10 | 0.3059 | 6.447568 | 1.972588 | 85.66 | immunoglobulin heavy variable 3-41 (pseudogene) |
| LDB2 | 1.15E-09 | 9.33E-11 | 0.3045 | 6.477436 | 1.972289 | 21.38 | LIM domain binding 2 |
| LOC107985055 | 8.37E-06 | 1.53E-06 | 0.4091 | 4.806632 | 1.966438 | 26.67 | uncharacterized LOC107985055 |
| LOC107985542 | 6.83E-09 | 6.31E-10 | 0.318 | 6.182427 | 1.966171 | 80.23 |  |
| KRT2 | 5.51E-09 | 5.01E-10 | 0.3153 | 6.218742 | 1.960477 | 10.77 | keratin 2 |
| IGLC2 | 3.70E-09 | 3.26E-10 | 0.3119 | 6.285667 | 1.960275 | 15971.48 | immunoglobulin lambda constant 2 |
| IGKV3-11 | 5.61E-09 | 5.11E-10 | 0.315 | 6.21573 | 1.958114 | 3818.74 | immunoglobulin kappa variable 3-11 |
| CYP4F29P | 1.52E-04 | 3.89E-05 | 0.4758 | 4.113703 | 1.957362 | 63.86 | cytochrome P450 family 4 subfamily F member 29, pseudogene |
| IGLVI-70 | 3.49E-03 | 1.33E-03 | 0.6028 | 3.209389 | 1.934632 | 148.87 | immunoglobulin lambda variable (I)-70 (pseudogene) |
| IGHJ5 | 1.27E-09 | 1.04E-10 | 0.2993 | 6.461696 | 1.933902 | 1115.1 | immunoglobulin heavy joining 5 |
| LGR6 | 1.72E-11 | 9.79E-13 | 0.2711 | 7.133383 | 1.933886 | 242.9 | leucine rich repeat containing G protein-coupled receptor 6 |
| IGKJ4 | 7.06E-10 | 5.45E-11 | 0.2935 | 6.558135 | 1.92484 | 1811.53 | immunoglobulin kappa joining 4 |
| NRCAM | 5.55E-06 | 9.70E-07 | 0.393 | 4.897633 | 1.924603 | 70.28 | neuronal cell adhesion molecule |
| KIR2DS4 | 3.71E-04 | 1.06E-04 | 0.4948 | 3.87711 | 1.91841 | 241.31 | killer cell immunoglobulin like receptor, two Ig domains and short cytoplasmic tail 4 |
| RNF165 | 1.12E-15 | 3.18E-17 | 0.2271 | 8.439677 | 1.917007 | 114.81 | ring finger protein 165 |
| LOC101927369 | 5.80E-08 | 6.59E-09 | 0.3304 | 5.801076 | 1.916543 | 66.99 | uncharacterized LOC101927369 |
| IGLC7 | 5.48E-06 | 9.57E-07 | 0.39 | 4.900202 | 1.911231 | 603.67 | immunoglobulin lambda constant 7 |
| IGLC3 | 7.54E-09 | 7.06E-10 | 0.3084 | 6.164751 | 1.901015 | 12694.73 | immunoglobulin lambda constant 3 (Kern-Oz+ marker) |
| IGKV3-7 | 1.79E-07 | 2.25E-08 | 0.3394 | 5.591756 | 1.89799 | 260.35 | immunoglobulin kappa variable 3-7 (non-functional) |
| KRT73 | 1.23E-04 | 3.05E-05 | 0.4546 | 4.169781 | 1.895658 | 136.13 | keratin 73 |
| LOC112268314 | 5.00E-05 | 1.11E-05 | 0.4295 | 4.393684 | 1.887071 | 19.53 | immunoglobulin kappa variable 1-39-like |
| IGKV5-2 | 5.65E-07 | 7.86E-08 | 0.3475 | 5.370288 | 1.866297 | 88.87 | immunoglobulin kappa variable 5-2 |
| IGKV1D-16 | 6.04E-08 | 6.87E-09 | 0.321 | 5.794025 | 1.859635 | 213 | immunoglobulin kappa variable 1D-16 |
| IGHV1-3 | 8.27E-03 | 3.56E-03 | 0.6353 | 2.914314 | 1.851426 | 397.17 | immunoglobulin heavy variable 1-3 |
| IGKV1D-8 | 2.75E-08 | 2.89E-09 | 0.3108 | 5.937677 | 1.845546 | 293.85 | immunoglobulin kappa variable 1D-8 |
| IGKV1D-17 | 6.43E-08 | 7.37E-09 | 0.319 | 5.782281 | 1.844755 | 148.8 | immunoglobulin kappa variable 1D-17 |
| IGHV1-69 | 3.63E-07 | 4.86E-08 | 0.3357 | 5.456465 | 1.831515 | 423.17 | immunoglobulin heavy variable 1-69 |
| IGHV4-55 | 4.87E-08 | 5.43E-09 | 0.313 | 5.833357 | 1.825598 | 191.87 | immunoglobulin heavy variable 4-55 (pseudogene) |
| ZNF215 | 3.87E-04 | 1.11E-04 | 0.4709 | 3.865307 | 1.820071 | 19.23 | zinc finger protein 215 |
| DENND2B | 2.48E-08 | 2.58E-09 | 0.3055 | 5.956424 | 1.819918 | 34.48 | DENN domain containing 2B |
| LOC107985032 | 1.60E-10 | 1.09E-11 | 0.2669 | 6.793585 | 1.813379 | 13.33 |  |
| PDGFD | 3.74E-12 | 1.90E-13 | 0.246 | 7.355978 | 1.80931 | 217.17 | platelet derived growth factor D |
| CDNF | 9.75E-11 | 6.44E-12 | 0.2628 | 6.86954 | 1.805536 | 20.9 | cerebral dopamine neurotrophic factor |
| LOC105375130 | 6.12E-07 | 8.59E-08 | 0.3364 | 5.354354 | 1.801433 | 194.92 | uncharacterized LOC105375130 |
| RPH3A | 3.42E-04 | 9.65E-05 | 0.4614 | 3.899347 | 1.799346 | 480.83 | rabphilin 3A |
| IGLV1-36 | 3.00E-08 | 3.18E-09 | 0.3035 | 5.921957 | 1.797048 | 268.93 | immunoglobulin lambda variable 1-36 |
| LOC107984889 | 1.79E-11 | 1.03E-12 | 0.2517 | 7.126912 | 1.793592 | 40.17 | uncharacterized LOC107984889 |
| IGKV1D-43 | 2.64E-04 | 7.21E-05 | 0.4517 | 3.969288 | 1.793057 | 53.96 | immunoglobulin kappa variable 1D-43 |
| DTHD1 | 1.20E-12 | 5.46E-14 | 0.2378 | 7.520347 | 1.788291 | 296.53 | death domain containing 1 |
| SLC44A5 | 1.80E-02 | 8.68E-03 | 0.6799 | 2.624402 | 1.784286 | 19.14 | solute carrier family 44 member 5 |
| IGHJ2 | 7.30E-08 | 8.51E-09 | 0.3097 | 5.758126 | 1.783568 | 180.99 | immunoglobulin heavy joining 2 |
| UBXN10-AS1 | 1.71E-05 | 3.38E-06 | 0.3815 | 4.646256 | 1.772577 | 11.97 | replaced by ID 391013 |
| LINC01801 | 4.19E-06 | 7.09E-07 | 0.3574 | 4.958756 | 1.772245 | 34.43 | long intergenic non-protein coding RNA 1801 |
| LOC102723407 | 7.99E-04 | 2.51E-04 | 0.4836 | 3.661367 | 1.77055 | 742.43 | immunoglobulin heavy variable 4-38-2-like |
| ANKRD36BP2 | 1.12E-08 | 1.09E-09 | 0.2903 | 6.095789 | 1.769456 | 62.34 | ankyrin repeat domain 36B pseudogene 2 |
| LOC102724104 | 5.43E-13 | 2.32E-14 | 0.2298 | 7.63173 | 1.753455 | 121.37 | uncharacterized LOC102724104 |
| KLRC4 | 1.30E-11 | 7.22E-13 | 0.2437 | 7.175159 | 1.748843 | 109.21 | killer cell lectin like receptor C4 |
| LOC107984667 | 5.66E-08 | 6.42E-09 | 0.3005 | 5.805539 | 1.744773 | 31.15 | replaced by ID 105370413 |
| LOC105372019 | 2.98E-08 | 3.16E-09 | 0.2938 | 5.923016 | 1.740212 | 21.99 | uncharacterized LOC105372019 |
| PRKN | 2.91E-05 | 6.08E-06 | 0.3846 | 4.52357 | 1.739774 | 9.93 | parkin RBR E3 ubiquitin protein ligase |
| LEXM | 2.34E-07 | 3.02E-08 | 0.313 | 5.540367 | 1.734063 | 26.99 | lymphocyte expansion molecule |
| IGLV3-12 | 1.03E-03 | 3.34E-04 | 0.4833 | 3.58758 | 1.733977 | 47.02 | immunoglobulin lambda variable 3-12 |
| KIR2DL1 | 9.12E-06 | 1.69E-06 | 0.362 | 4.787069 | 1.732888 | 168.44 | killer cell immunoglobulin like receptor, two Ig domains and long cytoplasmic tail 1 |
| LOC105375754 | 3.67E-06 | 6.12E-07 | 0.3453 | 4.987499 | 1.722058 | 83.01 | uncharacterized LOC105375754 |
| SLC16A14 | 2.27E-08 | 2.34E-09 | 0.2879 | 5.972225 | 1.719152 | 38.22 | solute carrier family 16 member 14 |
| KLRC1 | 2.81E-08 | 2.96E-09 | 0.2887 | 5.933928 | 1.713351 | 166.86 | killer cell lectin like receptor C1 |
| GNLY | 6.57E-08 | 7.56E-09 | 0.2963 | 5.778067 | 1.712113 | 14341.01 | granulysin |
| DEPDC1 | 1.20E-04 | 2.96E-05 | 0.4099 | 4.176572 | 1.711889 | 18.17 | DEP domain containing 1 |
| IGLV1-51 | 1.40E-09 | 1.16E-10 | 0.265 | 6.445043 | 1.707964 | 1004.26 | immunoglobulin lambda variable 1-51 |
| SIGLEC17P | 1.35E-12 | 6.24E-14 | 0.2275 | 7.502958 | 1.706959 | 272.93 | sialic acid binding Ig like lectin 17, pseudogene |
| TRDJ1 | 3.63E-12 | 1.83E-13 | 0.2314 | 7.360438 | 1.702998 | 155.22 | T cell receptor delta joining 1 |
| LINC01281 | 4.94E-06 | 8.52E-07 | 0.3454 | 4.92316 | 1.700378 | 12.6 | long intergenic non-protein coding RNA 1281 |
| KLRD1 | 2.69E-15 | 8.07E-17 | 0.204 | 8.33027 | 1.699416 | 2902.82 | killer cell lectin like receptor D1 |
| HOPX | 2.49E-20 | 4.13E-22 | 0.1757 | 9.667955 | 1.698909 | 735.35 | HOP homeobox |
| LOC105372105 | 9.10E-06 | 1.69E-06 | 0.3545 | 4.787652 | 1.697364 | 30.27 | uncharacterized LOC105372105 |
| GPRC5D | 1.73E-05 | 3.42E-06 | 0.3654 | 4.643688 | 1.696997 | 74.94 | G protein-coupled receptor class C group 5 member D |
| GLB1L2 | 1.15E-05 | 2.18E-06 | 0.3583 | 4.736236 | 1.696958 | 80.37 | galactosidase beta 1 like 2 |
| HEATR9 | 4.08E-08 | 4.47E-09 | 0.289 | 5.865782 | 1.695037 | 54.27 | HEAT repeat containing 9 |
| ADGRG1 | 1.73E-09 | 1.45E-10 | 0.2644 | 6.410918 | 1.694757 | 2225.4 | adhesion G protein-coupled receptor G1 |
| NINL | 1.56E-05 | 3.04E-06 | 0.363 | 4.668186 | 1.694499 | 43.65 | ninein like |
| GRIK4 | 3.40E-02 | 1.78E-02 | 0.7133 | 2.369712 | 1.690263 | 49.67 | glutamate ionotropic receptor kainate type subunit 4 |
| SH2D1B | 6.64E-17 | 1.64E-18 | 0.1925 | 8.779678 | 1.689667 | 850.75 | SH2 domain containing 1B |
| FZD4 | 1.66E-08 | 1.66E-09 | 0.28 | 6.027976 | 1.687928 | 17.42 | frizzled class receptor 4 |
| UNC45B | 6.94E-08 | 8.03E-09 | 0.2919 | 5.767795 | 1.683784 | 25.1 | unc-45 myosin chaperone B |
| IGHV3OR16-17 | 2.46E-05 | 5.05E-06 | 0.3688 | 4.562734 | 1.682647 | 19.16 | immunoglobulin heavy variable 3/OR16-17 (non-functional) |
| ENPP5 | 3.78E-10 | 2.77E-11 | 0.2526 | 6.658104 | 1.681579 | 112.06 | ectonucleotide pyrophosphatase/phosphodiesterase family member 5 |
| TRPV3 | 1.41E-07 | 1.74E-08 | 0.298 | 5.636117 | 1.679586 | 35.39 | transient receptor potential cation channel subfamily V member 3 |
| LOC107984225 | 2.37E-06 | 3.77E-07 | 0.3301 | 5.080361 | 1.677102 | 36.96 | uncharacterized LOC107984225 |
| IGHV3-7 | 4.57E-06 | 7.80E-07 | 0.3395 | 4.940399 | 1.677087 | 1353.73 | immunoglobulin heavy variable 3-7 |
| IGKV3D-7 | 2.06E-08 | 2.09E-09 | 0.2798 | 5.990516 | 1.675857 | 294.36 | immunoglobulin kappa variable 3D-7 |
| IGLV3-16 | 4.81E-06 | 8.26E-07 | 0.3391 | 4.929055 | 1.67147 | 107.56 | immunoglobulin lambda variable 3-16 |
| IGKV3D-11 | 2.43E-08 | 2.51E-09 | 0.2802 | 5.96048 | 1.670123 | 860.54 | immunoglobulin kappa variable 3D-11 |
| IGLV2-34 | 9.94E-09 | 9.54E-10 | 0.2725 | 6.11691 | 1.666842 | 195.02 | immunoglobulin lambda variable 2-34 (pseudogene) |
| LOC112267867 | 1.52E-10 | 1.03E-11 | 0.2446 | 6.801829 | 1.663979 | 116.63 |  |
| NCAM1 | 7.16E-13 | 3.17E-14 | 0.2186 | 7.591207 | 1.659232 | 364.67 | neural cell adhesion molecule 1 |
| IGHV1-45 | 4.53E-05 | 9.98E-06 | 0.3754 | 4.417637 | 1.658356 | 11.46 | immunoglobulin heavy variable 1-45 |
| KIR2DL3 | 3.00E-07 | 3.95E-08 | 0.3018 | 5.493241 | 1.657977 | 168.32 | killer cell immunoglobulin like receptor, two Ig domains and long cytoplasmic tail 3 |
| NCR1 | 3.39E-14 | 1.18E-15 | 0.2067 | 8.006748 | 1.654691 | 205.7 | natural cytotoxicity triggering receptor 1 |
| LOC730101 | 1.18E-03 | 3.89E-04 | 0.4629 | 3.547115 | 1.641956 | 19.66 | uncharacterized LOC730101 |
| C1orf21 | 1.22E-15 | 3.47E-17 | 0.1948 | 8.429385 | 1.641696 | 402.65 | chromosome 1 open reading frame 21 |
| TRGJP1 | 2.06E-09 | 1.74E-10 | 0.2565 | 6.382451 | 1.637225 | 18.48 | T cell receptor gamma joining P1 |
| PRSS30P | 2.45E-10 | 1.73E-11 | 0.243 | 6.726788 | 1.634776 | 64.57 | serine protease 30, pseudogene |
| LRFN2 | 1.21E-03 | 4.01E-04 | 0.4611 | 3.539163 | 1.631861 | 15.6 | leucine rich repeat and fibronectin type III domain containing 2 |
| IGHV2-5 | 1.24E-04 | 3.08E-05 | 0.3913 | 4.167559 | 1.630913 | 734.89 | immunoglobulin heavy variable 2-5 |
| CHPF | 4.11E-11 | 2.54E-12 | 0.2328 | 7.00086 | 1.629938 | 193.1 | chondroitin polymerizing factor |
| KRT72 | 7.51E-03 | 3.19E-03 | 0.5524 | 2.948915 | 1.629016 | 233.61 | keratin 72 |
| PTGDR | 7.20E-12 | 3.81E-13 | 0.2242 | 7.262235 | 1.628473 | 326.26 | prostaglandin D2 receptor |
| KLHL14 | 7.71E-07 | 1.10E-07 | 0.3054 | 5.308857 | 1.621434 | 269.85 | kelch like family member 14 |
| CDHR1 | 3.52E-08 | 3.80E-09 | 0.2749 | 5.892601 | 1.620102 | 104.48 | cadherin related family member 1 |
| LOC105370660 | 1.46E-16 | 3.74E-18 | 0.1865 | 8.686332 | 1.619995 | 106.35 | uncharacterized LOC105370660 |
| RGS9 | 6.00E-11 | 3.81E-12 | 0.2324 | 6.944219 | 1.614083 | 179.43 | regulator of G protein signaling 9 |
| RTP5 | 1.24E-05 | 2.37E-06 | 0.3416 | 4.719051 | 1.612053 | 17.29 | receptor transporter protein 5 (putative) |
| ERBB2 | 1.75E-13 | 6.88E-15 | 0.2062 | 7.78672 | 1.605801 | 366.13 | erb-b2 receptor tyrosine kinase 2 |
| LOC105369772 | 1.04E-03 | 3.36E-04 | 0.4478 | 3.585493 | 1.605661 | 27.08 | replaced by ID 319101 |
| CERCAM | 3.22E-07 | 4.27E-08 | 0.2929 | 5.479481 | 1.60501 | 78.19 | cerebral endothelial cell adhesion molecule |
| IGHV1-12 | 8.95E-05 | 2.15E-05 | 0.3775 | 4.249087 | 1.604073 | 22.27 | immunoglobulin heavy variable 1-12 (pseudogene) |
| IGHV3-48 | 2.12E-03 | 7.55E-04 | 0.4757 | 3.368867 | 1.602433 | 773.09 | immunoglobulin heavy variable 3-48 |
| KRT73-AS1 | 2.31E-06 | 3.65E-07 | 0.314 | 5.086539 | 1.597037 | 77.72 | KRT73 antisense RNA 1 |
| KIR3DL2 | 1.14E-06 | 1.69E-07 | 0.3048 | 5.230687 | 1.59439 | 196.08 | killer cell immunoglobulin like receptor, three Ig domains and long cytoplasmic tail 2 |
| IGKV1D-42 | 1.44E-04 | 3.65E-05 | 0.3856 | 4.128841 | 1.592157 | 92.85 | immunoglobulin kappa variable 1D-42 (non-functional) |
| SFRP5 | 4.89E-04 | 1.44E-04 | 0.4185 | 3.800689 | 1.590434 | 28.38 | secreted frizzled related protein 5 |
| ABCB9 | 3.47E-08 | 3.75E-09 | 0.2697 | 5.894852 | 1.589966 | 121.96 | ATP binding cassette subfamily B member 9 |
| CPNE7 | 4.30E-04 | 1.25E-04 | 0.4139 | 3.836504 | 1.58788 | 17.01 | copine 7 |
| IGHV1-69D | 5.71E-06 | 1.00E-06 | 0.3237 | 4.891274 | 1.583434 | 526.93 | immunoglobulin heavy variable 1-69D |
| LOC105377782 | 5.36E-03 | 2.18E-03 | 0.5164 | 3.065058 | 1.582876 | 140.59 | uncharacterized LOC105377782 |
| TRDC | 6.17E-13 | 2.70E-14 | 0.2074 | 7.61202 | 1.578525 | 3125.44 | T cell receptor delta constant |
| ENAM | 2.70E-05 | 5.61E-06 | 0.3475 | 4.540636 | 1.57789 | 18.34 | enamelin |
| IGLV3-1 | 1.71E-05 | 3.38E-06 | 0.3388 | 4.646423 | 1.574398 | 824.52 | immunoglobulin lambda variable 3-1 |
| BOK | 7.44E-07 | 1.06E-07 | 0.2962 | 5.3155 | 1.574216 | 89.82 | BCL2 family apoptosis regulator BOK |
| LOC107985236 | 8.16E-14 | 3.03E-15 | 0.1992 | 7.889674 | 1.57193 | 124.07 | uncharacterized LOC107985236 |
| LINC01163 | 3.03E-07 | 3.99E-08 | 0.2859 | 5.49122 | 1.569846 | 36.41 | long intergenic non-protein coding RNA 1163 |
| MMP23A | 1.93E-07 | 2.45E-08 | 0.281 | 5.576593 | 1.566931 | 16.78 | matrix metallopeptidase 23A (pseudogene) |
| IGLJ7 | 2.21E-03 | 7.90E-04 | 0.4662 | 3.356191 | 1.564704 | 16.48 | immunoglobulin lambda joining 7 |
| S1PR5 | 3.22E-11 | 1.95E-12 | 0.222 | 7.037767 | 1.562065 | 1172.67 | sphingosine-1-phosphate receptor 5 |
| IGHV4-34 | 8.03E-08 | 9.41E-09 | 0.2706 | 5.741087 | 1.553344 | 758.05 | immunoglobulin heavy variable 4-34 |
| DLG5 | 1.55E-12 | 7.28E-14 | 0.2075 | 7.482741 | 1.552847 | 232.89 | discs large MAGUK scaffold protein 5 |
| CLDND2 | 7.25E-09 | 6.74E-10 | 0.25 | 6.171958 | 1.542926 | 134.22 | claudin domain containing 2 |
| SPON2 | 8.92E-09 | 8.49E-10 | 0.2514 | 6.13555 | 1.542697 | 1086.39 | spondin 2 |
| PPP1R9A | 1.65E-06 | 2.54E-07 | 0.2986 | 5.154712 | 1.539141 | 26 | protein phosphatase 1 regulatory subunit 9A |
| TRGV9 | 2.00E-08 | 2.04E-09 | 0.2567 | 5.994952 | 1.539058 | 195.19 | T cell receptor gamma variable 9 |
| LINC02937 | 3.41E-06 | 5.64E-07 | 0.3072 | 5.003127 | 1.536972 | 46.42 | long intergenic non-protein coding RNA 2937 |
| LINC00612 | 2.67E-07 | 3.47E-08 | 0.2786 | 5.515829 | 1.536675 | 12.77 | long intergenic non-protein coding RNA 612 |
| LINC02481 | 5.09E-13 | 2.15E-14 | 0.2006 | 7.641549 | 1.532803 | 200.42 | long intergenic non-protein coding RNA 2481 |
| ADAMTS1 | 1.16E-06 | 1.73E-07 | 0.2913 | 5.226742 | 1.522757 | 85.57 | ADAM metallopeptidase with thrombospondin type 1 motif 1 |
| CFAP97D2 | 7.50E-03 | 3.18E-03 | 0.514 | 2.949672 | 1.516048 | 86 | CFAP97 domain containing 2 |
| DERL3 | 6.53E-08 | 7.49E-09 | 0.2623 | 5.779478 | 1.515789 | 265.78 | derlin 3 |
| LOC105379208 | 1.60E-03 | 5.48E-04 | 0.438 | 3.456215 | 1.513686 | 9.55 | uncharacterized LOC105379208 |
| LOC107986848 | 5.39E-05 | 1.21E-05 | 0.3458 | 4.375312 | 1.5129 | 67.12 |  |
| EBF4 | 2.71E-08 | 2.84E-09 | 0.2545 | 5.94039 | 1.511769 | 36.87 | EBF family member 4 |
| ARHGEF25 | 8.94E-08 | 1.06E-08 | 0.2639 | 5.721151 | 1.510002 | 17.02 | Rho guanine nucleotide exchange factor 25 |
| DDR2 | 1.26E-03 | 4.18E-04 | 0.4277 | 3.528229 | 1.509039 | 18.57 | discoidin domain receptor tyrosine kinase 2 |
| CLIC3 | 4.81E-09 | 4.33E-10 | 0.2416 | 6.241758 | 1.508308 | 221.1 | chloride intracellular channel 3 |
| KIF19 | 6.36E-05 | 1.46E-05 | 0.3478 | 4.334737 | 1.507714 | 37.09 | kinesin family member 19 |
| TRDV3 | 2.20E-03 | 7.87E-04 | 0.4484 | 3.357303 | 1.505362 | 31.65 | T cell receptor delta variable 3 |
| IGHV1-58 | 3.57E-04 | 1.01E-04 | 0.3871 | 3.887535 | 1.504793 | 26.94 | immunoglobulin heavy variable 1-58 |
| IGHV1OR15-1 | 2.25E-06 | 3.55E-07 | 0.295 | 5.091432 | 1.501948 | 158.9 | immunoglobulin heavy variable 1/OR15-1 (non-functional) |
| GIPR | 3.41E-10 | 2.48E-11 | 0.2249 | 6.674378 | 1.501226 | 78.61 | gastric inhibitory polypeptide receptor |
| LOC105371019 | 4.14E-04 | 1.20E-04 | 0.3901 | 3.847114 | 1.500748 | 11.7 | uncharacterized LOC105371019 |
| MIR4751 | 6.14E-11 | 3.90E-12 | 0.2161 | -6.94077 | -1.50005 | 51.22 | microRNA 4751 |
| ACSL4 | 6.56E-11 | 4.18E-12 | 0.2166 | -6.93083 | -1.50107 | 5536.96 | acyl-CoA synthetase long chain family member 4 |
| IGFLR1 | 2.75E-16 | 7.28E-18 | 0.1744 | -8.61037 | -1.50132 | 3858.74 | IGF like family receptor 1 |
| LOC107987121 | 3.11E-04 | 8.66E-05 | 0.3826 | -3.9254 | -1.50203 | 14.44 | uncharacterized LOC107987121 |
| ZNF267 | 2.09E-09 | 1.77E-10 | 0.2354 | -6.38007 | -1.50211 | 3020.63 | zinc finger protein 267 |
| FOLR2 | 7.45E-09 | 6.96E-10 | 0.2437 | -6.16701 | -1.50282 | 358.84 | folate receptor beta |
| IL1B | 1.90E-12 | 9.12E-14 | 0.2017 | -7.45309 | -1.50325 | 5010.61 | interleukin 1 beta |
| SLC1A3 | 1.10E-10 | 7.31E-12 | 0.2195 | -6.85146 | -1.50413 | 58.47 | solute carrier family 1 member 3 |
| H2BC4 | 7.89E-13 | 3.52E-14 | 0.1993 | -7.57772 | -1.50987 | 753.39 | H2B clustered histone 4 |
| LINC02818 | 5.39E-07 | 7.49E-08 | 0.2807 | -5.37911 | -1.50988 | 17.21 | long intergenic non-protein coding RNA 2818 |
| MRPL44 | 1.79E-27 | 1.22E-29 | 0.1336 | -11.3064 | -1.5106 | 2511.38 | mitochondrial ribosomal protein L44 |
| LOC105373148 | 1.17E-09 | 9.46E-11 | 0.2335 | -6.47538 | -1.51212 | 27.69 | uncharacterized LOC105373148 |
| CYLD-AS1 | 1.11E-27 | 7.44E-30 | 0.1334 | -11.3497 | -1.51429 | 413.63 | CYLD antisense RNA 1 |
| OR4F16 | 5.09E-06 | 8.79E-07 | 0.3082 | -4.91705 | -1.51528 | 39.32 | olfactory receptor family 4 subfamily F member 16 |
| ISG20 | 1.34E-19 | 2.35E-21 | 0.1597 | -9.48804 | -1.51564 | 10383.14 | interferon stimulated exonuclease gene 20 |
| BCL6 | 9.73E-09 | 9.33E-10 | 0.248 | -6.12047 | -1.51772 | 39480.97 | BCL6 transcription repressor |
| NUDT16L2P | 9.89E-06 | 1.85E-06 | 0.3186 | -4.76965 | -1.51984 | 398.71 | nudix hydrolase 16 like 2, pseudogene |
| LOC400499 | 2.11E-12 | 1.02E-13 | 0.2045 | -7.43819 | -1.52105 | 3022.84 | putative uncharacterized protein LOC400499 |
| DUSP3 | 4.33E-24 | 4.21E-26 | 0.144 | -10.5677 | -1.52155 | 3331.69 | dual specificity phosphatase 3 |
| G0S2 | 2.83E-06 | 4.61E-07 | 0.3019 | -5.04204 | -1.52216 | 90.11 | G0/G1 switch 2 |
| DNAH17 | 1.03E-09 | 8.25E-11 | 0.2343 | -6.49597 | -1.52216 | 1105.74 | dynein axonemal heavy chain 17 |
| CTSL | 3.37E-09 | 2.96E-10 | 0.2418 | -6.3007 | -1.52326 | 806.91 | cathepsin L |
| NRADDP | 9.14E-09 | 8.72E-10 | 0.2485 | -6.13131 | -1.52356 | 43.85 | neurotrophin receptor associated death domain, pseudogene |
| DRAM1 | 1.72E-24 | 1.57E-26 | 0.143 | -10.6597 | -1.52407 | 1404.3 | DNA damage regulated autophagy modulator 1 |
| PDE4B | 6.19E-18 | 1.32E-19 | 0.1683 | -9.05848 | -1.52497 | 6499.58 | phosphodiesterase 4B |
| TLR8 | 5.06E-13 | 2.13E-14 | 0.1996 | -7.64246 | -1.52521 | 11994.17 | toll like receptor 8 |
| DNAJC25-GNG10 | 3.03E-08 | 3.22E-09 | 0.2578 | -5.91988 | -1.5259 | 1338.1 | DNAJC25-GNG10 readthrough |
| LOC107984871 | 6.10E-13 | 2.66E-14 | 0.2004 | -7.61378 | -1.52593 | 46.67 | uncharacterized LOC107984871 |
| GNG10 | 2.84E-08 | 2.99E-09 | 0.2572 | -5.93205 | -1.52597 | 1344.43 | G protein subunit gamma 10 |
| CALCOCO2 | 6.42E-34 | 2.10E-36 | 0.1214 | -12.6003 | -1.52915 | 11809.45 | calcium binding and coiled-coil domain 2 |
| LOC105371934 | 9.14E-23 | 1.07E-24 | 0.1491 | -10.2598 | -1.52934 | 705.09 | uncharacterized LOC105371934 |
| MOV10 | 3.87E-21 | 5.81E-23 | 0.1552 | -9.86652 | -1.53126 | 5532.79 | Mov10 RISC complex RNA helicase |
| MSRB1 | 8.11E-14 | 3.00E-15 | 0.1941 | -7.89081 | -1.53131 | 21332.95 | methionine sulfoxide reductase B1 |
| LOC102724008 | 1.67E-04 | 4.32E-05 | 0.3745 | -4.08958 | -1.53139 | 26.91 | uncharacterized LOC102724008 |
| HLX | 4.46E-11 | 2.78E-12 | 0.2194 | -6.98846 | -1.53318 | 1840.44 | H2.0 like homeobox |
| TRG-TCC2-6 | 2.69E-04 | 7.38E-05 | 0.3868 | -3.96383 | -1.53336 | 20.35 | tRNA-Gly (anticodon TCC) 2-6 |
| KCNE5 | 3.76E-08 | 4.09E-09 | 0.2607 | -5.8807 | -1.53337 | 32.83 | potassium voltage-gated channel subfamily E regulatory subunit 5 |
| HSPA6 | 3.37E-11 | 2.05E-12 | 0.2182 | -7.03082 | -1.53405 | 10057.82 | heat shock protein family A (Hsp70) member 6 |
| PLEK | 5.40E-20 | 9.17E-22 | 0.1602 | -9.58589 | -1.53576 | 38390.65 | pleckstrin |
| TDRD7 | 4.61E-29 | 2.62E-31 | 0.1321 | -11.6388 | -1.537 | 2887.66 | tudor domain containing 7 |
| APOBEC3B-AS1 | 5.11E-17 | 1.25E-18 | 0.1747 | -8.81001 | -1.53906 | 608.46 | APOBEC3B antisense RNA 1 |
| CHRNB2 | 2.44E-27 | 1.69E-29 | 0.1367 | -11.2775 | -1.54125 | 17604.73 | cholinergic receptor nicotinic beta 2 subunit |
| MICB | 1.62E-19 | 2.86E-21 | 0.1629 | -9.46769 | -1.5425 | 2440.04 | MHC class I polypeptide-related sequence B |
| PSMB10 | 4.71E-22 | 6.16E-24 | 0.1531 | -10.0893 | -1.54429 | 9254.06 | proteasome 20S subunit beta 10 |
| LOC112268418 | 1.46E-05 | 2.83E-06 | 0.3299 | -4.68298 | -1.54483 | 27.03 | uncharacterized LOC112268418 |
| RGS1 | 3.62E-06 | 6.02E-07 | 0.3096 | -4.99044 | -1.54503 | 115.94 | regulator of G protein signaling 1 |
| UBQLNL | 1.06E-09 | 8.57E-11 | 0.2381 | -6.49029 | -1.54563 | 78.24 | ubiquilin like |
| MIR194-2HG | 8.83E-05 | 2.11E-05 | 0.3635 | -4.25307 | -1.54592 | 12.55 | MIR194-2 host gene |
| MAFB | 1.66E-13 | 6.46E-15 | 0.1985 | -7.79457 | -1.547 | 2098.87 | MAF bZIP transcription factor B |
| SP110 | 1.15E-30 | 5.42E-33 | 0.1293 | -11.965 | -1.54734 | 15965.94 | SP110 nuclear body protein |
| ADGRE5 | 1.89E-28 | 1.16E-30 | 0.1351 | -11.5113 | -1.55531 | 58642.46 | adhesion G protein-coupled receptor E5 |
| LOC105377499 | 3.75E-11 | 2.31E-12 | 0.2218 | -7.01461 | -1.55567 | 279.16 | uncharacterized LOC105377499 |
| SERPINI2 | 4.72E-06 | 8.09E-07 | 0.3155 | -4.93317 | -1.55637 | 19.18 | serpin family I member 2 |
| PLA2G7 | 5.52E-04 | 1.65E-04 | 0.4131 | -3.7675 | -1.55644 | 504.77 | phospholipase A2 group VII |
| H4C4 | 6.48E-07 | 9.12E-08 | 0.2915 | -5.34354 | -1.55755 | 32.71 | H4 clustered histone 4 |
| LOC105370121 | 7.94E-06 | 1.45E-06 | 0.3234 | -4.81838 | -1.55828 | 17.87 | uncharacterized LOC105370121 |
| GNS | 1.91E-23 | 2.02E-25 | 0.1496 | -10.4197 | -1.55873 | 16842.38 | glucosamine (N-acetyl)-6-sulfatase |
| H3P4 | 4.31E-05 | 9.45E-06 | 0.3525 | -4.42938 | -1.56145 | 21.54 | H3 histone pseudogene 4 |
| TRL-CAG1-6 | 4.92E-04 | 1.45E-04 | 0.4121 | -3.79882 | -1.56541 | 15.64 | tRNA-Leu (anticodon CAG) 1-6 |
| BCL3 | 1.87E-17 | 4.30E-19 | 0.1756 | -8.92897 | -1.56794 | 5503.26 | BCL3 transcription coactivator |
| RIPK2 | 3.58E-22 | 4.55E-24 | 0.155 | -10.1189 | -1.56806 | 1894.62 | receptor interacting serine/threonine kinase 2 |
| SELL | 1.30E-20 | 2.09E-22 | 0.1611 | -9.73718 | -1.5685 | 192898.3 | selectin L |
| MVP | 3.85E-32 | 1.47E-34 | 0.1281 | -12.2606 | -1.57087 | 20482.32 | major vault protein |
| IRF9 | 1.22E-28 | 7.12E-31 | 0.136 | -11.5531 | -1.57148 | 12609.83 | interferon regulatory factor 9 |
| PSME1 | 6.05E-32 | 2.45E-34 | 0.1288 | -12.2195 | -1.57351 | 31043.79 | proteasome activator subunit 1 |
| MIR21 | 1.21E-17 | 2.69E-19 | 0.1755 | -8.98067 | -1.57569 | 40.01 | microRNA 21 |
| KIAA0040 | 2.01E-20 | 3.30E-22 | 0.163 | -9.6909 | -1.5794 | 17102.03 | KIAA0040 |
| LINC00487 | 4.08E-03 | 1.59E-03 | 0.5003 | -3.15694 | -1.57953 | 33.53 | long intergenic non-protein coding RNA 487 |
| CNDP2 | 1.78E-23 | 1.87E-25 | 0.1515 | -10.427 | -1.58006 | 9202.24 | carnosine dipeptidase 2 |
| PSMB8 | 2.63E-27 | 1.84E-29 | 0.1404 | -11.2703 | -1.58262 | 16485.08 | proteasome 20S subunit beta 8 |
| PKN2-AS1 | 5.51E-04 | 1.65E-04 | 0.4204 | -3.76807 | -1.58398 | 16.66 | PKN2 antisense RNA 1 |
| LINC00877 | 2.17E-24 | 2.01E-26 | 0.1493 | -10.6366 | -1.58802 | 670.44 | long intergenic non-protein coding RNA 877 |
| MIR3614 | 2.04E-12 | 9.87E-14 | 0.2137 | -7.4427 | -1.5905 | 276.43 | microRNA 3614 |
| SMPDL3B | 2.54E-32 | 9.28E-35 | 0.1297 | -12.298 | -1.59498 | 118.69 | sphingomyelin phosphodiesterase acid like 3B |
| LOC101929750 | 2.98E-07 | 3.91E-08 | 0.2903 | -5.49473 | -1.59519 | 38.51 | uncharacterized LOC101929750 |
| CREG1 | 2.66E-14 | 9.12E-16 | 0.1985 | -8.03809 | -1.59556 | 5106.43 | cellular repressor of E1A stimulated genes 1 |
| CFAP58 | 3.02E-13 | 1.23E-14 | 0.2071 | -7.71299 | -1.59724 | 182.32 | cilia and flagella associated protein 58 |
| SLC22A4 | 2.35E-12 | 1.15E-13 | 0.2153 | -7.42216 | -1.598 | 1732.56 | solute carrier family 22 member 4 |
| TMEM268 | 9.11E-25 | 8.01E-27 | 0.149 | -10.7222 | -1.59811 | 2228.9 | transmembrane protein 268 |
| LOC107984200 | 1.22E-29 | 6.28E-32 | 0.136 | -11.7599 | -1.59895 | 149.54 | uncharacterized LOC107984200 |
| WDR86 | 3.13E-25 | 2.58E-27 | 0.1478 | -10.8263 | -1.59974 | 942.23 | WD repeat domain 86 |
| H4C5 | 3.35E-05 | 7.15E-06 | 0.3564 | -4.48916 | -1.59992 | 21.16 | H4 clustered histone 5 |
| ATG3 | 9.95E-21 | 1.58E-22 | 0.1639 | -9.76561 | -1.60083 | 6867.02 | autophagy related 3 |
| LOC100419583 | 2.22E-29 | 1.20E-31 | 0.1368 | -11.7052 | -1.60095 | 4631.34 | ring finger protein 4 pseudogene |
| CRB2 | 6.65E-17 | 1.65E-18 | 0.1824 | -8.77929 | -1.6014 | 301.5 | crumbs cell polarity complex component 2 |
| DISC2 | 1.01E-09 | 8.09E-11 | 0.2465 | -6.49888 | -1.60185 | 19.41 | disrupted in schizophrenia 2 |
| PIK3AP1 | 1.07E-17 | 2.37E-19 | 0.1784 | -8.99474 | -1.60502 | 10726.08 | phosphoinositide-3-kinase adaptor protein 1 |
| APOBEC3A | 2.06E-18 | 4.10E-20 | 0.175 | -9.1854 | -1.60743 | 9044.35 | apolipoprotein B mRNA editing enzyme catalytic subunit 3A |
| NMRAL2P | 4.04E-04 | 1.17E-04 | 0.4172 | -3.85302 | -1.60745 | 12.29 | NmrA like redox sensor 2, pseudogene |
| CSF2RB | 6.38E-15 | 2.01E-16 | 0.1957 | -8.2214 | -1.60897 | 45703.22 | colony stimulating factor 2 receptor subunit beta |
| EPB41L3 | 2.51E-21 | 3.69E-23 | 0.1625 | -9.91202 | -1.61086 | 1840.98 | erythrocyte membrane protein band 4.1 like 3 |
| LOC105377587 | 2.01E-05 | 4.04E-06 | 0.3495 | -4.60927 | -1.61089 | 86.99 | uncharacterized LOC105377587 |
| CARD16 | 1.46E-12 | 6.82E-14 | 0.2151 | -7.49127 | -1.61132 | 6033.15 | caspase recruitment domain family member 16 |
| LOC105369736 | 2.00E-06 | 3.12E-07 | 0.315 | -5.11597 | -1.61151 | 16.07 | uncharacterized LOC105369736 |
| SIPA1L2 | 5.20E-08 | 5.83E-09 | 0.2772 | -5.82156 | -1.61347 | 1931.46 | signal induced proliferation associated 1 like 2 |
| CDCP1 | 5.60E-09 | 5.09E-10 | 0.2597 | -6.21633 | -1.61425 | 81.1 | CUB domain containing protein 1 |
| LOC107986113 | 1.75E-06 | 2.70E-07 | 0.314 | -5.14327 | -1.61499 | 30.12 |  |
| KPTN | 2.39E-23 | 2.56E-25 | 0.1554 | -10.3969 | -1.6158 | 402.3 | kaptin, actin binding protein |
| H2AC18 | 4.26E-10 | 3.16E-11 | 0.2436 | -6.6391 | -1.61707 | 828.75 | H2A clustered histone 18 |
| CHMP5 | 6.91E-11 | 4.47E-12 | 0.2337 | -6.92154 | -1.6173 | 2534.41 | charged multivesicular body protein 5 |
| ZCCHC2 | 3.13E-19 | 5.68E-21 | 0.1721 | -9.39574 | -1.61746 | 4870.4 | zinc finger CCHC-type containing 2 |
| H2AC19 | 5.11E-10 | 3.84E-11 | 0.2449 | -6.61003 | -1.6189 | 892.38 | H2A clustered histone 19 |
| FAM177B | 5.76E-21 | 8.85E-23 | 0.1648 | -9.82434 | -1.61948 | 412.48 | family with sequence similarity 177 member B |
| ALDH1A1 | 1.16E-17 | 2.57E-19 | 0.1803 | -8.98576 | -1.62002 | 2378.48 | aldehyde dehydrogenase 1 family member A1 |
| LY96 | 9.69E-11 | 6.40E-12 | 0.2358 | -6.87053 | -1.62024 | 766.24 | lymphocyte antigen 96 |
| ACTA2 | 4.76E-09 | 4.28E-10 | 0.2596 | -6.2435 | -1.62066 | 644.74 | actin alpha 2, smooth muscle |
| H2BP1 | 5.48E-07 | 7.62E-08 | 0.3017 | -5.37588 | -1.62211 | 16.12 | H2B histone pseudogene 1 |
| FGL2 | 4.85E-22 | 6.41E-24 | 0.1609 | -10.0853 | -1.623 | 40640.91 | fibrinogen like 2 |
| TRIM5 | 7.09E-30 | 3.60E-32 | 0.1375 | -11.8067 | -1.6231 | 2995.7 | tripartite motif containing 5 |
| LOC107985047 | 3.32E-06 | 5.48E-07 | 0.3244 | -5.0086 | -1.62484 | 33.31 |  |
| VCPIP1 | 3.10E-16 | 8.32E-18 | 0.1892 | -8.5951 | -1.62627 | 4789.33 | valosin containing protein interacting protein 1 |
| LOC101927272 | 2.28E-16 | 5.97E-18 | 0.1889 | -8.63306 | -1.63049 | 284.5 | uncharacterized LOC101927272 |
| AP5B1 | 3.90E-18 | 8.07E-20 | 0.1791 | -9.11223 | -1.63164 | 10869.73 | adaptor related protein complex 5 subunit beta 1 |
| PARP10 | 2.60E-20 | 4.32E-22 | 0.1689 | -9.66331 | -1.63178 | 4497.04 | poly(ADP-ribose) polymerase family member 10 |
| LOC105377449 | 2.33E-14 | 7.92E-16 | 0.2027 | -8.0554 | -1.63298 | 123.21 |  |
| RERE | 9.18E-28 | 6.12E-30 | 0.1437 | -11.3668 | -1.63309 | 13908.68 | arginine-glutamic acid dipeptide repeats |
| RCVRN | 1.55E-06 | 2.37E-07 | 0.3161 | -5.16746 | -1.63368 | 18.19 | recoverin |
| LOC112267968 | 4.12E-06 | 6.95E-07 | 0.33 | -4.96263 | -1.63776 | 18.87 | uncharacterized LOC112267968 |
| MEFV | 3.76E-22 | 4.85E-24 | 0.162 | -10.1128 | -1.63794 | 13707.31 | MEFV innate immunity regulator, pyrin |
| LOC105378443 | 1.18E-23 | 1.20E-25 | 0.1567 | -10.4688 | -1.64016 | 3652.62 | uncharacterized LOC105378443 |
| PLAUR | 4.85E-15 | 1.49E-16 | 0.1986 | -8.25701 | -1.64025 | 7271.75 | plasminogen activator, urokinase receptor |
| LOC107986304 | 3.95E-18 | 8.25E-20 | 0.1802 | -9.10989 | -1.64138 | 77.95 |  |
| DYSF | 7.62E-12 | 4.05E-13 | 0.2263 | -7.25393 | -1.64192 | 23270.94 | dysferlin |
| GNB4 | 5.06E-20 | 8.57E-22 | 0.1714 | -9.5928 | -1.64445 | 3218.29 | G protein subunit beta 4 |
| CASP4 | 2.10E-20 | 3.45E-22 | 0.1699 | -9.68635 | -1.64552 | 19624.36 | caspase 4 |
| RAB39A | 5.25E-10 | 3.97E-11 | 0.2495 | -6.60534 | -1.64785 | 50.44 | RAB39A, member RAS oncogene family |
| ZNFX1 | 5.56E-35 | 1.55E-37 | 0.1288 | -12.8044 | -1.64914 | 14796.06 | zinc finger NFX1-type containing 1 |
| CASP1 | 2.97E-28 | 1.90E-30 | 0.144 | -11.4685 | -1.65098 | 19950.22 | caspase 1 |
| LOC105374263 | 5.44E-05 | 1.23E-05 | 0.3782 | -4.37252 | -1.65384 | 13.64 | uncharacterized LOC105374263 |
| ZBP1 | 5.56E-23 | 6.35E-25 | 0.1607 | -10.31 | -1.65671 | 4450.35 | Z-DNA binding protein 1 |
| ACTA2-AS1 | 3.13E-08 | 3.33E-09 | 0.2804 | -5.91437 | -1.65825 | 262.37 | ACTA2 antisense RNA 1 |
| PSMD6-AS2 | 3.52E-11 | 2.15E-12 | 0.2362 | -7.02448 | -1.65891 | 576.27 | PSMD6 antisense RNA 2 |
| APOL3 | 2.41E-28 | 1.53E-30 | 0.1447 | -11.4874 | -1.66224 | 8900.01 | apolipoprotein L3 |
| SLC31A2 | 1.43E-24 | 1.29E-26 | 0.1561 | -10.6782 | -1.66669 | 4549.3 | solute carrier family 31 member 2 |
| CYBB | 7.75E-24 | 7.75E-26 | 0.1591 | -10.5103 | -1.67178 | 34509.67 | cytochrome b-245 beta chain |
| GAS8 | 1.23E-16 | 3.14E-18 | 0.1922 | -8.70645 | -1.67364 | 200.44 | growth arrest specific 8 |
| GPR42 | 3.74E-07 | 5.02E-08 | 0.3077 | -5.45061 | -1.67722 | 45.54 | G protein-coupled receptor 42 |
| LRRK2 | 5.29E-14 | 1.89E-15 | 0.2113 | -7.94808 | -1.67908 | 29249.31 | leucine rich repeat kinase 2 |
| MFSD6L | 4.96E-12 | 2.54E-13 | 0.2297 | -7.31677 | -1.68063 | 117.45 | major facilitator superfamily domain containing 6 like |
| SAT1 | 1.07E-22 | 1.28E-24 | 0.1643 | -10.2424 | -1.68282 | 34682.32 | spermidine/spermine N1-acetyltransferase 1 |
| LOC105371873 | 1.35E-07 | 1.66E-08 | 0.2982 | -5.64386 | -1.68302 | 18.4 |  |
| PCBP1-AS1 | 2.99E-11 | 1.81E-12 | 0.2388 | -7.04835 | -1.68328 | 1150.69 | PCBP1 antisense RNA 1 |
| ZNF200 | 5.74E-18 | 1.22E-19 | 0.1857 | -9.06751 | -1.68338 | 2352.22 | zinc finger protein 200 |
| IGSF6 | 1.40E-17 | 3.16E-19 | 0.1881 | -8.96315 | -1.68616 | 17858.28 | immunoglobulin superfamily member 6 |
| BAZ1A | 3.55E-21 | 5.32E-23 | 0.1708 | -9.87543 | -1.68642 | 11877.3 | bromodomain adjacent to zinc finger domain 1A |
| P2RX7 | 4.46E-20 | 7.52E-22 | 0.1756 | -9.60624 | -1.68649 | 1588.02 | purinergic receptor P2X 7 |
| LINC00513 | 5.36E-05 | 1.21E-05 | 0.3857 | -4.37666 | -1.68795 | 13.89 | long intergenic non-protein coding RNA 513 |
| CDC42EP2 | 2.18E-17 | 5.12E-19 | 0.1895 | -8.90972 | -1.68852 | 3429.27 | CDC42 effector protein 2 |
| MIR3945HG | 1.66E-06 | 2.55E-07 | 0.3282 | -5.15428 | -1.69176 | 483.39 | MIR3945 host gene |
| DENND1A | 2.02E-33 | 6.85E-36 | 0.1354 | -12.5069 | -1.69402 | 3298.67 | DENN domain containing 1A |
| MIR9902-1 | 3.30E-05 | 7.03E-06 | 0.3771 | -4.49284 | -1.69442 | 14.15 | microRNA 9902-1 |
| LOC105370635 | 6.82E-06 | 1.22E-06 | 0.3494 | -4.8526 | -1.69535 | 23.18 | uncharacterized LOC105370635 |
| CES1 | 1.91E-04 | 5.02E-05 | 0.4181 | -4.0548 | -1.69542 | 1259.33 | carboxylesterase 1 |
| WDFY1 | 3.96E-24 | 3.81E-26 | 0.1603 | -10.5771 | -1.69603 | 3944.9 | WD repeat and FYVE domain containing 1 |
| BST2 | 5.09E-17 | 1.24E-18 | 0.1926 | -8.81067 | -1.6965 | 7410.23 | bone marrow stromal cell antigen 2 |
| IL15RA | 5.27E-30 | 2.59E-32 | 0.1434 | -11.8345 | -1.69687 | 724 | interleukin 15 receptor subunit alpha |
| LOC105374985 | 1.31E-11 | 7.27E-13 | 0.2368 | -7.17417 | -1.6987 | 388.88 |  |
| CYREN | 4.31E-28 | 2.83E-30 | 0.1487 | -11.434 | -1.69972 | 12163.23 | cell cycle regulator of NHEJ |
| C1QA | 1.33E-06 | 2.00E-07 | 0.3272 | -5.19945 | -1.7014 | 425.89 | complement C1q A chain |
| EGR2 | 3.08E-12 | 1.54E-13 | 0.2308 | -7.38364 | -1.70383 | 69.62 | early growth response 2 |
| LOC105374102 | 5.89E-13 | 2.55E-14 | 0.2236 | -7.61922 | -1.7039 | 255.34 | uncharacterized LOC105374102 |
| MLKL | 6.94E-20 | 1.19E-21 | 0.1783 | -9.55887 | -1.70419 | 6269.18 | mixed lineage kinase domain like pseudokinase |
| LINC01841 | 7.92E-08 | 9.27E-09 | 0.2968 | -5.74363 | -1.70481 | 20.66 | long intergenic non-protein coding RNA 1841 |
| PSMB8-AS1 | 5.02E-27 | 3.62E-29 | 0.1522 | -11.2106 | -1.70661 | 11694.33 | PSMB8 antisense RNA 1 (head to head) |
| LOC107984880 | 2.27E-11 | 1.33E-12 | 0.2415 | -7.09069 | -1.71209 | 65.7 |  |
| LOC105375035 | 7.48E-06 | 1.35E-06 | 0.3544 | -4.83151 | -1.71248 | 24.2 | uncharacterized LOC105375035 |
| ALPK1 | 3.04E-21 | 4.51E-23 | 0.1737 | -9.89205 | -1.71832 | 8124.86 | alpha kinase 1 |
| URAHP | 3.37E-15 | 1.02E-16 | 0.2071 | -8.30263 | -1.71958 | 58.05 | urate (hydroxyiso-) hydrolase, pseudogene |
| CCR1 | 1.24E-23 | 1.28E-25 | 0.1648 | -10.4631 | -1.72465 | 13323.24 | C-C motif chemokine receptor 1 |
| EIF2AK2 | 2.31E-18 | 4.65E-20 | 0.1881 | -9.17187 | -1.72525 | 5855.64 | eukaryotic translation initiation factor 2 alpha kinase 2 |
| PSORS1C3 | 9.27E-03 | 4.06E-03 | 0.6007 | -2.87308 | -1.72581 | 38.3 | psoriasis susceptibility 1 candidate 3 |
| SBNO2 | 7.74E-20 | 1.34E-21 | 0.1808 | -9.54624 | -1.72592 | 10951.57 | strawberry notch homolog 2 |
| KCNJ10 | 5.05E-05 | 1.13E-05 | 0.3932 | -4.39103 | -1.72635 | 21.59 | potassium inwardly rectifying channel subfamily J member 10 |
| CASP7 | 3.00E-12 | 1.49E-13 | 0.2337 | -7.38791 | -1.72639 | 1763.63 | caspase 7 |
| NGFR | 3.61E-08 | 3.91E-09 | 0.2936 | -5.88793 | -1.72863 | 50.3 | nerve growth factor receptor |
| RERE-AS1 | 3.59E-16 | 9.76E-18 | 0.202 | -8.57676 | -1.73271 | 457 | RERE antisense RNA 1 |
| TMEM272 | 2.89E-13 | 1.17E-14 | 0.225 | -7.71917 | -1.73708 | 793.52 | transmembrane protein 272 |
| MED12L | 3.10E-17 | 7.35E-19 | 0.1959 | -8.86948 | -1.73783 | 109.94 | mediator complex subunit 12L |
| FLVCR2-AS1 | 4.41E-15 | 1.35E-16 | 0.2105 | -8.26892 | -1.74037 | 67.15 | FLVCR2 antisense RNA 1 |
| LGALS3BP | 1.14E-10 | 7.57E-12 | 0.2542 | -6.84643 | -1.74042 | 2471.1 | galectin 3 binding protein |
| FFAR3 | 1.24E-07 | 1.51E-08 | 0.3085 | -5.66016 | -1.74603 | 105.44 | free fatty acid receptor 3 |
| JAK2 | 8.39E-18 | 1.82E-19 | 0.1937 | -9.02361 | -1.74805 | 4435.57 | Janus kinase 2 |
| TRPV4 | 3.23E-07 | 4.28E-08 | 0.3191 | -5.47886 | -1.74843 | 64.83 | transient receptor potential cation channel subfamily V member 4 |
| BISPR | 1.28E-26 | 9.54E-29 | 0.1574 | -11.1244 | -1.75052 | 1260.23 | BST2 interferon stimulated positive regulator |
| CES1P2 | 4.26E-05 | 9.32E-06 | 0.3953 | -4.43241 | -1.75215 | 48.72 | carboxylesterase 1 pseudogene 2 |
| NMI | 9.84E-23 | 1.17E-24 | 0.172 | -10.2512 | -1.76309 | 6340.31 | N-myc and STAT interactor |
| FLVCR2 | 2.41E-22 | 3.00E-24 | 0.1735 | -10.1598 | -1.76321 | 926.42 | FLVCR heme transporter 2 |
| RPAP3-DT | 1.28E-16 | 3.26E-18 | 0.2031 | -8.70188 | -1.76744 | 412.27 | RPAP3 divergent transcript |
| RALB | 2.18E-19 | 3.91E-21 | 0.1877 | -9.43502 | -1.77105 | 16533.98 | RAS like proto-oncogene B |
| IGSF10 | 9.56E-12 | 5.18E-13 | 0.2453 | -7.22056 | -1.77148 | 36.99 | immunoglobulin superfamily member 10 |
| FHDC1 | 1.43E-10 | 9.68E-12 | 0.2603 | -6.81122 | -1.77325 | 335.99 | FH2 domain containing 1 |
| NCF1B | 2.24E-23 | 2.39E-25 | 0.1707 | -10.4036 | -1.7756 | 20182.32 | neutrophil cytosolic factor 1B pseudogene |
| SNX10-AS1 | 7.44E-11 | 4.83E-12 | 0.2573 | -6.91035 | -1.7777 | 336.36 | SNX10 antisense RNA 1 |
| LOC105370816 | 1.64E-05 | 3.21E-06 | 0.3818 | -4.65701 | -1.77787 | 13.47 | uncharacterized LOC105370816 |
| PRLR | 9.85E-07 | 1.45E-07 | 0.339 | -5.25891 | -1.78301 | 61.43 | prolactin receptor |
| DAPP1 | 6.97E-21 | 1.09E-22 | 0.1822 | -9.80366 | -1.78596 | 7643.45 | dual adaptor of phosphotyrosine and 3-phosphoinositides 1 |
| ZNF438 | 1.19E-22 | 1.44E-24 | 0.1747 | -10.2311 | -1.78711 | 2313.27 | zinc finger protein 438 |
| SHOC1 | 2.60E-11 | 1.55E-12 | 0.2532 | -7.07002 | -1.78982 | 83.38 | shortage in chiasmata 1 |
| DHRS9 | 3.24E-16 | 8.72E-18 | 0.2087 | -8.58967 | -1.79239 | 3685.43 | dehydrogenase/reductase 9 |
| CES1P1 | 7.26E-05 | 1.69E-05 | 0.4169 | -4.30189 | -1.79336 | 38.03 | carboxylesterase 1 pseudogene 1 |
| MIR9902-2 | 2.90E-05 | 6.07E-06 | 0.397 | -4.52392 | -1.79595 | 22.92 | microRNA 9902-2 |
| KCNJ2-AS1 | 9.08E-12 | 4.90E-13 | 0.2486 | -7.22799 | -1.7972 | 157.93 | KCNJ2 antisense RNA 1 |
| RBCK1 | 5.13E-25 | 4.43E-27 | 0.1668 | -10.7769 | -1.79805 | 11882.56 | RANBP2-type and C3HC4-type zinc finger containing 1 |
| SIGLEC1 | 1.20E-04 | 2.97E-05 | 0.4314 | -4.17559 | -1.80114 | 659.41 | sialic acid binding Ig like lectin 1 |
| SYNDIG1L | 9.84E-10 | 7.85E-11 | 0.2775 | -6.50345 | -1.80483 | 27.45 | synapse differentiation inducing 1 like |
| NCF1C | 5.94E-23 | 6.85E-25 | 0.1754 | -10.3028 | -1.80713 | 21656.4 | neutrophil cytosolic factor 1C pseudogene |
| RNF213-AS1 | 6.59E-11 | 4.22E-12 | 0.2619 | -6.92971 | -1.81469 | 528.33 | RNF213 antisense RNA 1 |
| NRIR | 1.32E-06 | 1.98E-07 | 0.349 | -5.20089 | -1.81511 | 127.27 | negative regulator of interferon response |
| NUB1 | 8.46E-43 | 1.25E-45 | 0.1281 | -14.1783 | -1.81621 | 9137.11 | negative regulator of ubiquitin like proteins 1 |
| NUDT16-DT | 5.90E-14 | 2.14E-15 | 0.2292 | -7.93303 | -1.81822 | 40.18 | NUDT16 divergent transcript |
| SQOR | 1.90E-28 | 1.19E-30 | 0.1581 | -11.5092 | -1.81958 | 11979.93 | sulfide quinone oxidoreductase |
| SOCS3 | 2.79E-11 | 1.68E-12 | 0.2582 | -7.05901 | -1.82282 | 3717.07 | suppressor of cytokine signaling 3 |
| MARCO | 1.98E-11 | 1.14E-12 | 0.2567 | -7.11225 | -1.82542 | 540.37 | macrophage receptor with collagenous structure |
| SIPA1L1 | 2.06E-34 | 6.07E-37 | 0.1438 | -12.698 | -1.82592 | 9660.77 | signal induced proliferation associated 1 like 1 |
| SNX10 | 1.29E-12 | 5.93E-14 | 0.2433 | -7.50956 | -1.827 | 5221.54 | sorting nexin 10 |
| CETP | 5.68E-22 | 7.63E-24 | 0.182 | -10.0683 | -1.83288 | 205.01 | cholesteryl ester transfer protein |
| NPC2 | 1.88E-27 | 1.29E-29 | 0.1623 | -11.3013 | -1.83367 | 10588.02 | NPC intracellular cholesterol transporter 2 |
| LOC105375924 | 1.57E-08 | 1.56E-09 | 0.304 | -6.03763 | -1.83573 | 54.33 | uncharacterized LOC105375924 |
| LINC02773 | 6.58E-11 | 4.21E-12 | 0.265 | -6.92992 | -1.83666 | 24.42 | long intergenic non-protein coding RNA 2773 |
| HORMAD1 | 1.90E-11 | 1.09E-12 | 0.258 | -7.11837 | -1.83674 | 56.56 | HORMA domain containing 1 |
| LOC107987044 | 1.01E-15 | 2.85E-17 | 0.2181 | -8.45271 | -1.84334 | 19.46 |  |
| TRANK1 | 3.32E-32 | 1.23E-34 | 0.1506 | -12.2752 | -1.8491 | 22046.06 | tetratricopeptide repeat and ankyrin repeat containing 1 |
| NCF1 | 2.91E-20 | 4.87E-22 | 0.1918 | -9.65104 | -1.85075 | 35499.6 | neutrophil cytosolic factor 1 |
| IFIT5 | 1.47E-21 | 2.09E-23 | 0.1858 | -9.96856 | -1.85189 | 3366.27 | interferon induced protein with tetratricopeptide repeats 5 |
| ASPHD2 | 9.51E-27 | 6.96E-29 | 0.1661 | -11.1526 | -1.85295 | 739.85 | aspartate beta-hydroxylase domain containing 2 |
| MIR4709 | 5.58E-25 | 4.84E-27 | 0.1723 | -10.7686 | -1.85534 | 1801.55 | microRNA 4709 |
| LOC107985279 | 3.39E-04 | 9.53E-05 | 0.4769 | -3.90221 | -1.86104 | 33.08 | uncharacterized LOC107985279 |
| OAS2 | 6.85E-13 | 3.01E-14 | 0.2456 | -7.59769 | -1.86629 | 14725.85 | 2'-5'-oligoadenylate synthetase 2 |
| VSIG10 | 3.18E-07 | 4.20E-08 | 0.3408 | -5.48211 | -1.86817 | 345.56 | V-set and immunoglobulin domain containing 10 |
| LINC02701 | 2.58E-07 | 3.35E-08 | 0.3384 | -5.52201 | -1.8687 | 29.26 | long intergenic non-protein coding RNA 2701 |
| H1-6 | 1.29E-07 | 1.58E-08 | 0.3308 | -5.65256 | -1.87015 | 12.22 | H1.6 linker histone, cluster member |
| TTC26 | 5.92E-07 | 8.28E-08 | 0.3491 | -5.36091 | -1.87134 | 293.53 | tetratricopeptide repeat domain 26 |
| DYNLT1 | 1.11E-20 | 1.77E-22 | 0.1923 | -9.75416 | -1.87546 | 5366.81 | dynein light chain Tctex-type 1 |
| FFAR2 | 4.78E-22 | 6.30E-24 | 0.186 | -10.0871 | -1.87646 | 22488.28 | free fatty acid receptor 2 |
| DDX60L | 3.35E-21 | 4.99E-23 | 0.19 | -9.88179 | -1.87771 | 14214.17 | DExD/H-box 60 like |
| RMI2 | 1.67E-10 | 1.15E-11 | 0.2768 | -6.78628 | -1.8784 | 291.31 | RecQ mediated genome instability 2 |
| IFI30 | 3.58E-22 | 4.55E-24 | 0.1857 | -10.119 | -1.87871 | 61273.67 | IFI30 lysosomal thiol reductase |
| FAM241A | 5.27E-15 | 1.63E-16 | 0.228 | -8.2463 | -1.87982 | 670.38 | family with sequence similarity 241 member A |
| RPS16P5 | 1.86E-16 | 4.86E-18 | 0.2186 | -8.65665 | -1.89245 | 84.64 | ribosomal protein S16 pseudogene 5 |
| LINC01531 | 3.04E-12 | 1.51E-13 | 0.257 | -7.38592 | -1.89786 | 38.97 | long intergenic non-protein coding RNA 1531 |
| LOC105372412 | 7.69E-09 | 7.23E-10 | 0.3081 | -6.16106 | -1.89828 | 61.64 | phospholipase A2 inhibitor and Ly6/PLAUR domain-containing protein-like |
| CFAP58-DT | 9.73E-13 | 4.39E-14 | 0.2521 | -7.54903 | -1.90318 | 253.12 | CFAP58 divergent transcript |
| GADD45B | 3.72E-25 | 3.15E-27 | 0.1771 | -10.8082 | -1.91409 | 5004.06 | growth arrest and DNA damage inducible beta |
| NOD2 | 5.81E-14 | 2.10E-15 | 0.2415 | -7.93523 | -1.91627 | 5909.69 | nucleotide binding oligomerization domain containing 2 |
| NSG2 | 1.34E-04 | 3.37E-05 | 0.4623 | -4.14716 | -1.91709 | 14.06 | neuronal vesicle trafficking associated 2 |
| MT2A | 5.73E-13 | 2.47E-14 | 0.2516 | -7.62334 | -1.91766 | 1766.9 | metallothionein 2A |
| TRIM21 | 2.49E-32 | 8.97E-35 | 0.156 | -12.3008 | -1.91932 | 11149.44 | tripartite motif containing 21 |
| LACTB | 1.57E-28 | 9.37E-31 | 0.1668 | -11.5295 | -1.9229 | 2878.02 | lactamase beta |
| POLB | 2.43E-31 | 1.02E-33 | 0.1592 | -12.1028 | -1.92719 | 2858.63 | DNA polymerase beta |
| MAB21L3 | 1.47E-10 | 9.98E-12 | 0.2833 | -6.80678 | -1.92819 | 26.27 | mab-21 like 3 |
| RNF213 | 4.08E-32 | 1.58E-34 | 0.1574 | -12.2549 | -1.9293 | 52245.86 | ring finger protein 213 |
| IFITM1 | 1.41E-28 | 8.31E-31 | 0.1672 | -11.5398 | -1.92939 | 71323.64 | interferon induced transmembrane protein 1 |
| LOC105372801 | 8.32E-11 | 5.45E-12 | 0.2802 | -6.89331 | -1.93161 | 549.38 | uncharacterized LOC105372801 |
| TMEM150B | 1.16E-17 | 2.59E-19 | 0.2151 | -8.98507 | -1.93302 | 560.34 | transmembrane protein 150B |
| PLAAT4 | 1.81E-15 | 5.27E-17 | 0.2316 | -8.38041 | -1.94095 | 9499.93 | phospholipase A and acyltransferase 4 |
| AFF1 | 3.36E-23 | 3.73E-25 | 0.1874 | -10.361 | -1.9416 | 8749.44 | ALF transcription elongation factor 1 |
| SYNPO2 | 7.73E-13 | 3.44E-14 | 0.2563 | -7.58045 | -1.94254 | 59.74 | synaptopodin 2 |
| TBC1D30 | 1.89E-18 | 3.71E-20 | 0.2114 | -9.19619 | -1.94403 | 347.07 | TBC1 domain family member 30 |
| HELZ2 | 5.40E-30 | 2.69E-32 | 0.1649 | -11.8312 | -1.95068 | 4846.51 | helicase with zinc finger 2 |
| LOC101927741 | 4.21E-10 | 3.11E-11 | 0.2941 | -6.64112 | -1.95325 | 116.91 | uncharacterized LOC101927741 |
| PSMB9 | 2.09E-30 | 9.93E-33 | 0.1641 | -11.9146 | -1.95567 | 19865.55 | proteasome 20S subunit beta 9 |
| LIMK2 | 5.63E-22 | 7.50E-24 | 0.1946 | -10.07 | -1.95975 | 23057.51 | LIM domain kinase 2 |
| ZMYND15 | 3.21E-16 | 8.64E-18 | 0.2284 | -8.59077 | -1.96205 | 254.01 | zinc finger MYND-type containing 15 |
| PRMT5-DT | 4.15E-09 | 3.69E-10 | 0.3136 | -6.26672 | -1.96524 | 32.74 | PRMT5 divergent transcript |
| LOC105373033 | 1.09E-17 | 2.41E-19 | 0.2192 | -8.99268 | -1.97075 | 145.57 | uncharacterized LOC105373033 |
| TFEC | 7.59E-14 | 2.79E-15 | 0.2497 | -7.89986 | -1.97275 | 1306.35 | transcription factor EC |
| TMEM252 | 8.94E-12 | 4.82E-13 | 0.273 | -7.23027 | -1.97406 | 157.51 | transmembrane protein 252 |
| IFI16 | 1.04E-31 | 4.30E-34 | 0.1627 | -12.1735 | -1.98121 | 36737.54 | interferon gamma inducible protein 16 |
| DHRS12 | 1.22E-18 | 2.33E-20 | 0.2152 | -9.24582 | -1.98954 | 2820.52 | dehydrogenase/reductase 12 |
| LOC105371529 | 1.02E-11 | 5.56E-13 | 0.2764 | -7.2108 | -1.9931 | 214.52 | uncharacterized LOC105371529 |
| HFE-AS1 | 5.43E-15 | 1.69E-16 | 0.2424 | -8.24194 | -1.99772 | 135.39 | HFE antisense RNA 1 |
| LOC102724237 | 1.05E-04 | 2.58E-05 | 0.4749 | -4.20803 | -1.99831 | 28.98 |  |
| LOC101927522 | 1.90E-17 | 4.38E-19 | 0.226 | -8.92692 | -2.01772 | 306.25 | uncharacterized LOC101927522 |
| TNFAIP2 | 5.97E-32 | 2.38E-34 | 0.1655 | -12.2217 | -2.02223 | 49950.76 | TNF alpha induced protein 2 |
| LINC01232 | 2.45E-22 | 3.06E-24 | 0.1993 | -10.1577 | -2.02492 | 236.2 | long intergenic non-protein coding RNA 1232 |
| TMEM252-DT | 3.92E-09 | 3.48E-10 | 0.3231 | -6.27584 | -2.02795 | 54.86 | TMEM252 divergent transcript |
| SLC26A8 | 6.36E-10 | 4.88E-11 | 0.3104 | -6.57449 | -2.04104 | 657.55 | solute carrier family 26 member 8 |
| LOC101927243 | 8.82E-08 | 1.04E-08 | 0.3568 | -5.72373 | -2.04244 | 18.71 | uncharacterized LOC101927243 |
| KREMEN1 | 2.14E-07 | 2.74E-08 | 0.3677 | -5.55726 | -2.04366 | 4186.58 | kringle containing transmembrane protein 1 |
| SMCO4 | 1.20E-24 | 1.07E-26 | 0.1912 | -10.6957 | -2.04515 | 1599.59 | single-pass membrane protein with coiled-coil domains 4 |
| ADAMTSL4-AS2 | 3.61E-22 | 4.62E-24 | 0.2022 | -10.1176 | -2.04596 | 379.12 | ADAMTSL4 antisense RNA 2 |
| KCNJ2 | 4.82E-17 | 1.17E-18 | 0.2333 | -8.81729 | -2.05738 | 7453.57 | potassium inwardly rectifying channel subfamily J member 2 |
| TMEM140 | 5.40E-30 | 2.71E-32 | 0.1741 | -11.8306 | -2.05992 | 9536.81 | transmembrane protein 140 |
| IFIT1 | 4.36E-09 | 3.89E-10 | 0.3297 | -6.25838 | -2.06354 | 8433.7 | interferon induced protein with tetratricopeptide repeats 1 |
| H4C8 | 5.49E-15 | 1.72E-16 | 0.2505 | -8.24041 | -2.06402 | 139.67 | H4 clustered histone 8 |
| SPATS2L | 5.41E-13 | 2.31E-14 | 0.2705 | -7.63224 | -2.06465 | 583.16 | spermatogenesis associated serine rich 2 like |
| RAB20 | 1.14E-11 | 6.28E-13 | 0.2872 | -7.19434 | -2.0663 | 1374.69 | RAB20, member RAS oncogene family |
| CLEC9A | 1.74E-17 | 3.96E-19 | 0.2319 | -8.93801 | -2.07284 | 314.74 | C-type lectin domain containing 9A |
| LINC00189 | 3.96E-06 | 6.64E-07 | 0.4176 | -4.97156 | -2.07624 | 275.91 | long intergenic non-protein coding RNA 189 |
| MDK | 3.02E-12 | 1.50E-13 | 0.2815 | -7.38674 | -2.0797 | 69.36 | midkine |
| CXCL16 | 4.82E-18 | 1.02E-19 | 0.2292 | -9.08695 | -2.08315 | 7942.98 | C-X-C motif chemokine ligand 16 |
| BMX | 6.05E-09 | 5.53E-10 | 0.3365 | -6.20327 | -2.08718 | 648.99 | BMX non-receptor tyrosine kinase |
| LOC105374412 | 1.04E-09 | 8.33E-11 | 0.3221 | -6.49447 | -2.09163 | 35.07 | replaced by ID 105374413 |
| MSRB2 | 3.61E-18 | 7.46E-20 | 0.2294 | -9.12079 | -2.09199 | 1865.55 | methionine sulfoxide reductase B2 |
| SUCNR1 | 6.54E-07 | 9.21E-08 | 0.3924 | -5.3417 | -2.09627 | 121.37 | succinate receptor 1 |
| HTR3B | 2.66E-06 | 4.29E-07 | 0.4146 | -5.05576 | -2.09631 | 19.25 | 5-hydroxytryptamine receptor 3B |
| SOD2 | 1.69E-22 | 2.09E-24 | 0.2061 | -10.1947 | -2.10132 | 155959 | superoxide dismutase 2 |
| PLSCR4 | 1.76E-04 | 4.57E-05 | 0.5171 | -4.07654 | -2.10782 | 15.65 | phospholipid scramblase 4 |
| MAFF | 1.37E-18 | 2.64E-20 | 0.2287 | -9.23276 | -2.11186 | 673.85 | MAF bZIP transcription factor F |
| ADAMTSL4-AS1 | 6.95E-18 | 1.49E-19 | 0.2335 | -9.04569 | -2.11213 | 225.37 | ADAMTSL4 antisense RNA 1 |
| DTX3L | 1.65E-41 | 2.79E-44 | 0.1516 | -13.9586 | -2.11609 | 14142.34 | deltex E3 ubiquitin ligase 3L |
| DDX60 | 1.14E-22 | 1.37E-24 | 0.2069 | -10.2359 | -2.11754 | 4307.21 | DExD/H-box helicase 60 |
| TAP2 | 1.89E-40 | 3.82E-43 | 0.1555 | -13.7708 | -2.1407 | 18374.13 | transporter 2, ATP binding cassette subfamily B member |
| RHBDF2 | 4.42E-34 | 1.40E-36 | 0.1708 | -12.6324 | -2.15784 | 7727.07 | rhomboid 5 homolog 2 |
| CORIN | 7.52E-12 | 3.99E-13 | 0.2975 | -7.25585 | -2.15842 | 76.98 | corin, serine peptidase |
| MX1 | 3.02E-12 | 1.51E-13 | 0.2933 | -7.3867 | -2.16673 | 17667.6 | MX dynamin like GTPase 1 |
| CD59 | 1.28E-18 | 2.44E-20 | 0.2348 | -9.24088 | -2.16991 | 9647.76 | CD59 molecule (CD59 blood group) |
| C11orf91 | 1.02E-13 | 3.86E-15 | 0.2774 | -7.85929 | -2.18003 | 37.7 | chromosome 11 open reading frame 91 |
| LOC105377067 | 2.49E-06 | 3.98E-07 | 0.43 | -5.07001 | -2.18028 | 404.32 | uncharacterized LOC105377067 |
| IFIT2 | 9.21E-21 | 1.45E-22 | 0.2233 | -9.77413 | -2.18294 | 27012.32 | interferon induced protein with tetratricopeptide repeats 2 |
| LOC107984945 | 2.40E-21 | 3.49E-23 | 0.2222 | -9.91771 | -2.20374 | 179.42 | replaced by ID 64744 |
| XRN1 | 1.26E-23 | 1.30E-25 | 0.2107 | -10.4611 | -2.20408 | 7188.86 | 5'-3' exoribonuclease 1 |
| VPS9D1-AS1 | 1.43E-22 | 1.77E-24 | 0.2159 | -10.211 | -2.20434 | 551.81 | VPS9D1 antisense RNA 1 |
| ICAM1 | 1.56E-27 | 1.06E-29 | 0.1948 | -11.319 | -2.20475 | 6369.3 | intercellular adhesion molecule 1 |
| LOC105371461 | 1.79E-15 | 5.21E-17 | 0.2645 | -8.38181 | -2.21668 | 273.08 | replaced by ID 9447 |
| KCNJ15 | 1.92E-15 | 5.62E-17 | 0.2648 | -8.37295 | -2.21689 | 24875.44 | potassium inwardly rectifying channel subfamily J member 15 |
| TRC-GCA4-1 | 4.33E-14 | 1.53E-15 | 0.2792 | -7.97479 | -2.22651 | 17.87 | tRNA-Cys (anticodon GCA) 4-1 |
| TNFSF10 | 3.01E-29 | 1.66E-31 | 0.1908 | -11.6775 | -2.22833 | 25665.57 | TNF superfamily member 10 |
| CACNA1E | 2.37E-18 | 4.78E-20 | 0.2431 | -9.16893 | -2.2288 | 635.77 | calcium voltage-gated channel subunit alpha1 E |
| GRAMD1B | 7.74E-24 | 7.70E-26 | 0.2121 | -10.5109 | -2.22968 | 2035.85 | GRAM domain containing 1B |
| HCAR2 | 4.71E-19 | 8.65E-21 | 0.2385 | -9.35145 | -2.23045 | 11151.09 | hydroxycarboxylic acid receptor 2 |
| PLSCR2 | 2.96E-08 | 3.13E-09 | 0.3769 | -5.9248 | -2.23316 | 23.09 | phospholipid scramblase 2 |
| TNFSF13B | 1.89E-28 | 1.16E-30 | 0.1945 | -11.5113 | -2.23851 | 7391.76 | TNF superfamily member 13b |
| OAS1 | 2.67E-16 | 7.07E-18 | 0.2602 | -8.61384 | -2.24135 | 12934.7 | 2'-5'-oligoadenylate synthetase 1 |
| OASL | 1.73E-17 | 3.93E-19 | 0.2511 | -8.93888 | -2.24473 | 5851.07 | 2'-5'-oligoadenylate synthetase like |
| ERLIN1 | 1.03E-14 | 3.30E-16 | 0.2757 | -8.16169 | -2.25014 | 3357.52 | ER lipid raft associated 1 |
| NUCB1 | 2.56E-31 | 1.09E-33 | 0.1861 | -12.0973 | -2.2515 | 19564 | nucleobindin 1 |
| MIR7703 | 2.06E-26 | 1.56E-28 | 0.2032 | -11.0807 | -2.25178 | 689.14 | microRNA 7703 |
| SCO2 | 3.47E-20 | 5.83E-22 | 0.2348 | -9.63253 | -2.26189 | 1320.47 | synthesis of cytochrome C oxidase 2 |
| MUC1 | 5.68E-26 | 4.31E-28 | 0.2061 | -10.9891 | -2.26436 | 77.53 | mucin 1, cell surface associated |
| LOC105374071 | 1.89E-40 | 3.73E-43 | 0.1651 | -13.7725 | -2.27335 | 2067.07 | uncharacterized LOC105374071 |
| GCH1 | 3.26E-25 | 2.75E-27 | 0.2106 | -10.8207 | -2.27848 | 4578.18 | GTP cyclohydrolase 1 |
| CARD17P | 5.01E-22 | 6.65E-24 | 0.2261 | -10.0818 | -2.27909 | 915.37 | caspase recruitment domain family member 17, pseudogene |
| TGM2 | 1.71E-13 | 6.70E-15 | 0.2926 | -7.78994 | -2.27916 | 857.39 | transglutaminase 2 |
| ASPRV1 | 1.38E-15 | 3.94E-17 | 0.2714 | -8.4147 | -2.28378 | 1600.06 | aspartic peptidase retroviral like 1 |
| LOC112268267 | 9.44E-33 | 3.25E-35 | 0.1852 | -12.3825 | -2.29357 | 9039.39 | uncharacterized LOC112268267 |
| TRAFD1 | 1.69E-35 | 4.62E-38 | 0.1779 | -12.898 | -2.29515 | 13394.12 | TRAF-type zinc finger domain containing 1 |
| NUCB1-AS1 | 2.09E-34 | 6.38E-37 | 0.1816 | -12.694 | -2.30476 | 2112.69 | NUCB1 antisense RNA 1 |
| HCAR3 | 3.96E-18 | 8.29E-20 | 0.2535 | -9.10935 | -2.30879 | 13814.67 | hydroxycarboxylic acid receptor 3 |
| IFIH1 | 4.78E-39 | 1.04E-41 | 0.1709 | -13.5297 | -2.31257 | 5346.71 | interferon induced with helicase C domain 1 |
| GBP3 | 4.81E-23 | 5.47E-25 | 0.2244 | -10.3244 | -2.31643 | 3809.33 | guanylate binding protein 3 |
| PSME2 | 3.44E-39 | 7.14E-42 | 0.1713 | -13.5577 | -2.32232 | 17465.46 | proteasome activator subunit 2 |
| RIGI | 4.03E-31 | 1.79E-33 | 0.1929 | -12.0565 | -2.3252 | 10140.73 | RNA sensor RIG-I |
| EGR3 | 1.24E-13 | 4.70E-15 | 0.2977 | -7.83471 | -2.33277 | 94.45 | early growth response 3 |
| KLHDC7B-DT | 5.21E-21 | 7.96E-23 | 0.2373 | -9.83492 | -2.33362 | 296.42 | KLHDC7B divergent transcript |
| TCN2 | 4.24E-18 | 8.89E-20 | 0.2566 | -9.10174 | -2.33593 | 894.88 | transcobalamin 2 |
| ATP1B2 | 6.52E-08 | 7.48E-09 | 0.4047 | -5.77968 | -2.33883 | 71.95 | ATPase Na+/K+ transporting subunit beta 2 |
| TYMP | 5.40E-24 | 5.28E-26 | 0.2219 | -10.5464 | -2.33998 | 17054.77 | thymidine phosphorylase |
| HMGA2-AS1 | 4.03E-10 | 2.97E-11 | 0.3536 | -6.64826 | -2.35096 | 34.57 | HMGA2 antisense RNA 1 |
| DOCK4 | 1.65E-15 | 4.77E-17 | 0.2805 | -8.3922 | -2.3536 | 1385.55 | dedicator of cytokinesis 4 |
| RTP4 | 9.17E-22 | 1.26E-23 | 0.2351 | -10.0186 | -2.35579 | 1373.15 | receptor transporter protein 4 |
| IRF1 | 1.42E-38 | 3.26E-41 | 0.1753 | -13.4457 | -2.35695 | 67001.51 | interferon regulatory factor 1 |
| ODF3B | 3.41E-23 | 3.80E-25 | 0.2276 | -10.3593 | -2.35829 | 1031.98 | outer dense fiber of sperm tails 3B |
| AK4 | 3.96E-22 | 5.12E-24 | 0.2335 | -10.1074 | -2.35959 | 94.59 | adenylate kinase 4 |
| STX11 | 4.32E-29 | 2.43E-31 | 0.2036 | -11.645 | -2.37106 | 10348.6 | syntaxin 11 |
| APOL2 | 7.49E-41 | 1.39E-43 | 0.1715 | -13.8436 | -2.37409 | 15608.3 | apolipoprotein L2 |
| TRIM6 | 8.75E-12 | 4.70E-13 | 0.3282 | -7.23382 | -2.37447 | 100.46 | tripartite motif containing 6 |
| TAP1 | 1.58E-40 | 3.02E-43 | 0.1729 | -13.7877 | -2.38376 | 42586.37 | transporter 1, ATP binding cassette subfamily B member |
| IRF7 | 1.30E-20 | 2.09E-22 | 0.2457 | -9.7371 | -2.39257 | 5787.6 | interferon regulatory factor 7 |
| STK3 | 3.61E-23 | 4.05E-25 | 0.2317 | -10.3532 | -2.39881 | 966.27 | serine/threonine kinase 3 |
| LOC105374304 | 1.78E-21 | 2.56E-23 | 0.2419 | -9.94851 | -2.40681 | 117.17 | uncharacterized LOC105374304 |
| KLF5 | 2.42E-13 | 9.66E-15 | 0.3114 | -7.74371 | -2.41157 | 574.03 | KLF transcription factor 5 |
| STAT2 | 8.85E-45 | 9.18E-48 | 0.1664 | -14.519 | -2.41589 | 21206.98 | signal transducer and activator of transcription 2 |
| CCRL2 | 9.92E-19 | 1.88E-20 | 0.2607 | -9.26894 | -2.41679 | 714.78 | C-C motif chemokine receptor like 2 |
| LOC102724608 | 2.03E-24 | 1.88E-26 | 0.2273 | -10.6431 | -2.41881 | 243.76 | uncharacterized LOC102724608 |
| FAS | 1.43E-26 | 1.07E-28 | 0.2178 | -11.1141 | -2.42058 | 7485.24 | Fas cell surface death receptor |
| LOC107985224 | 4.03E-13 | 1.66E-14 | 0.3159 | -7.67436 | -2.42446 | 36.83 | uncharacterized LOC107985224 |
| C15orf48 | 2.55E-09 | 2.20E-10 | 0.3825 | -6.34678 | -2.42795 | 34.15 | chromosome 15 open reading frame 48 |
| IL27 | 1.28E-25 | 1.01E-27 | 0.2228 | -10.912 | -2.43111 | 107.59 | interleukin 27 |
| GBP2 | 1.50E-44 | 1.64E-47 | 0.168 | -14.4792 | -2.4327 | 51141.39 | guanylate binding protein 2 |
| TRIM22 | 2.95E-36 | 7.59E-39 | 0.1876 | -13.0365 | -2.44523 | 35762.42 | tripartite motif containing 22 |
| PML | 5.15E-34 | 1.66E-36 | 0.1946 | -12.6189 | -2.45554 | 8686.98 | PML nuclear body scaffold |
| CD300LD | 1.24E-05 | 2.36E-06 | 0.5237 | -4.72015 | -2.47187 | 173.75 | CD300 molecule like family member d |
| IFITM3 | 5.40E-14 | 1.95E-15 | 0.3113 | -7.9446 | -2.47346 | 67111.36 | interferon induced transmembrane protein 3 |
| GRIN3A | 1.99E-19 | 3.54E-21 | 0.2621 | -9.44552 | -2.47592 | 242.08 | glutamate ionotropic receptor NMDA type subunit 3A |
| SORT1 | 3.90E-29 | 2.17E-31 | 0.2125 | -11.6547 | -2.47634 | 7204.2 | sortilin 1 |
| C4BPA | 6.96E-04 | 2.14E-04 | 0.6704 | -3.70152 | -2.4815 | 1531.55 | complement component 4 binding protein alpha |
| XAF1 | 1.17E-26 | 8.60E-29 | 0.2232 | -11.1337 | -2.48493 | 10140.1 | XIAP associated factor 1 |
| ANXA3 | 1.76E-09 | 1.47E-10 | 0.389 | -6.40814 | -2.49278 | 7327.47 | annexin A3 |
| VPS9D1 | 1.82E-29 | 9.64E-32 | 0.2135 | -11.7237 | -2.50298 | 2901.77 | VPS9 domain containing 1 |
| MIR4257 | 1.55E-17 | 3.50E-19 | 0.281 | -8.95183 | -2.51539 | 58.82 | microRNA 4257 |
| CCR5AS | 2.11E-13 | 8.37E-15 | 0.3249 | -7.76184 | -2.5217 | 200.89 | CCR5 antisense RNA |
| LINC01094 | 2.24E-14 | 7.58E-16 | 0.3137 | -8.06074 | -2.52849 | 383.2 | long intergenic non-protein coding RNA 1094 |
| PRRG4 | 4.03E-31 | 1.81E-33 | 0.2101 | -12.0558 | -2.53316 | 3831.8 | proline rich and Gla domain 4 |
| HERC5 | 6.95E-20 | 1.20E-21 | 0.2666 | -9.55847 | -2.54865 | 6419.38 | HECT and RLD domain containing E3 ubiquitin protein ligase 5 |
| IFI44 | 2.64E-14 | 9.04E-16 | 0.3173 | -8.03921 | -2.55104 | 5068.42 | interferon induced protein 44 |
| MSR1 | 9.96E-08 | 1.19E-08 | 0.4485 | -5.70116 | -2.55718 | 286.41 | macrophage scavenger receptor 1 |
| FBXO39 | 1.11E-05 | 2.09E-06 | 0.5399 | -4.74413 | -2.56117 | 20.79 | F-box protein 39 |
| GK3 | 1.36E-21 | 1.93E-23 | 0.2568 | -9.97683 | -2.56193 | 110.97 | glycerol kinase 3 |
| IFI6 | 1.43E-15 | 4.10E-17 | 0.3048 | -8.41007 | -2.56339 | 7849.26 | interferon alpha inducible protein 6 |
| AIM2 | 1.44E-29 | 7.56E-32 | 0.219 | -11.7442 | -2.57237 | 2444.82 | absent in melanoma 2 |
| TIMM10 | 8.37E-16 | 2.33E-17 | 0.3039 | -8.47596 | -2.57619 | 1904.27 | translocase of inner mitochondrial membrane 10 |
| LOC105378841 | 6.16E-31 | 2.83E-33 | 0.2148 | -12.0189 | -2.58146 | 310.03 | uncharacterized LOC105378841 |
| FBXO6 | 6.00E-35 | 1.70E-37 | 0.2023 | -12.797 | -2.58842 | 2606.82 | F-box protein 6 |
| PARP9 | 4.26E-43 | 5.82E-46 | 0.1819 | -14.2317 | -2.58941 | 20508.1 | poly(ADP-ribose) polymerase family member 9 |
| APOL6 | 1.24E-46 | 1.09E-49 | 0.1761 | -14.8201 | -2.60978 | 16227.33 | apolipoprotein L6 |
| SLC6A12 | 1.68E-19 | 2.99E-21 | 0.2764 | -9.46318 | -2.61521 | 710.58 | solute carrier family 6 member 12 |
| ISG15 | 7.60E-10 | 5.91E-11 | 0.4003 | -6.54608 | -2.62057 | 4688.75 | ISG15 ubiquitin like modifier |
| SLC6A12-AS1 | 7.94E-15 | 2.53E-16 | 0.3202 | -8.19375 | -2.62342 | 18.99 | SLC6A12 antisense RNA 1 |
| TIFA | 3.85E-24 | 3.68E-26 | 0.2485 | -10.5802 | -2.62883 | 3007.68 | TRAF interacting protein with forkhead associated domain |
| LPCAT2 | 1.07E-21 | 1.50E-23 | 0.2645 | -10.0018 | -2.6453 | 11759.24 | lysophosphatidylcholine acyltransferase 2 |
| AANAT | 3.73E-22 | 4.79E-24 | 0.2623 | -10.1139 | -2.6526 | 141.65 | aralkylamine N-acetyltransferase |
| RUFY4 | 7.80E-21 | 1.23E-22 | 0.2721 | -9.79129 | -2.66403 | 201.86 | RUN and FYVE domain containing 4 |
| GPR84 | 1.27E-24 | 1.13E-26 | 0.2502 | -10.69 | -2.67466 | 344.01 | G protein-coupled receptor 84 |
| SDC3 | 2.60E-24 | 2.43E-26 | 0.2519 | -10.619 | -2.67533 | 360.15 | syndecan 3 |
| FRMD3 | 1.30E-32 | 4.60E-35 | 0.2169 | -12.3545 | -2.67958 | 771.72 | FERM domain containing 3 |
| CLEC6A | 3.25E-10 | 2.36E-11 | 0.4058 | -6.68204 | -2.71171 | 79.11 | C-type lectin domain containing 6A |
| SAMD9L | 5.39E-38 | 1.29E-40 | 0.2037 | -13.3434 | -2.71751 | 18747.95 | sterile alpha motif domain containing 9 like |
| LOC107986193 | 2.22E-14 | 7.50E-16 | 0.3376 | -8.06213 | -2.72164 | 17.7 | uncharacterized LOC107986193 |
| LOC101926887 | 2.77E-24 | 2.60E-26 | 0.2567 | -10.6128 | -2.72419 | 53.21 | uncharacterized LOC101926887 |
| CFAP97D1 | 3.65E-11 | 2.24E-12 | 0.3893 | -7.01884 | -2.73246 | 15.76 | CFAP97 domain containing 1 |
| GK | 9.93E-26 | 7.65E-28 | 0.2526 | -10.9373 | -2.76264 | 11069.78 | glycerol kinase |
| XXYLT1-AS2 | 2.39E-18 | 4.83E-20 | 0.3021 | -9.16775 | -2.7697 | 73.39 | XXYLT1 antisense RNA 2 |
| PXT1 | 2.51E-10 | 1.78E-11 | 0.4135 | -6.72302 | -2.78023 | 19.38 | peroxisomal testis enriched protein 1 |
| STAT1 | 1.20E-61 | 6.54E-66 | 0.1639 | -17.1477 | -2.81091 | 50584.06 | signal transducer and activator of transcription 1 |
| OAS3 | 2.03E-18 | 4.01E-20 | 0.306 | -9.18768 | -2.8118 | 16541.47 | 2'-5'-oligoadenylate synthetase 3 |
| LOC105378085 | 1.05E-14 | 3.39E-16 | 0.3455 | -8.15875 | -2.81858 | 164.07 | uncharacterized LOC105378085 |
| UBE2L6 | 1.00E-46 | 8.21E-50 | 0.1918 | -14.8389 | -2.84632 | 30470.71 | ubiquitin conjugating enzyme E2 L6 |
| C1QC | 5.77E-08 | 6.56E-09 | 0.4909 | -5.80189 | -2.84819 | 127.43 | complement C1q C chain |
| FGF13 | 4.83E-09 | 4.35E-10 | 0.4566 | -6.24074 | -2.8493 | 122.05 | fibroblast growth factor 13 |
| C1QB | 2.10E-17 | 4.90E-19 | 0.3216 | -8.91443 | -2.86659 | 444.65 | complement C1q B chain |
| EPSTI1 | 3.18E-25 | 2.64E-27 | 0.2651 | -10.8244 | -2.86957 | 8928.32 | epithelial stromal interaction 1 |
| LOC105374898 | 9.01E-14 | 3.37E-15 | 0.3647 | -7.87647 | -2.87273 | 98.54 | uncharacterized LOC105374898 |
| PLSCR1 | 2.85E-31 | 1.23E-33 | 0.2382 | -12.0875 | -2.879 | 8482.44 | phospholipid scramblase 1 |
| SAMD4A | 1.71E-28 | 1.03E-30 | 0.2501 | -11.5216 | -2.88127 | 618.27 | sterile alpha motif domain containing 4A |
| SNHG28 | 2.90E-24 | 2.74E-26 | 0.2719 | -10.6078 | -2.88446 | 329.81 | small nucleolar RNA host gene 28 |
| PSTPIP2 | 4.49E-31 | 2.04E-33 | 0.2395 | -12.0459 | -2.88466 | 9064.35 | proline-serine-threonine phosphatase interacting protein 2 |
| SECTM1 | 4.79E-39 | 1.07E-41 | 0.2136 | -13.5277 | -2.88969 | 32232.31 | secreted and transmembrane 1 |
| IFI35 | 4.13E-39 | 8.81E-42 | 0.2142 | -13.5422 | -2.9003 | 8416.64 | interferon induced protein 35 |
| APOL1 | 2.59E-56 | 5.67E-60 | 0.178 | -16.3339 | -2.90743 | 8141.98 | apolipoprotein L1 |
| PARP14 | 8.03E-47 | 6.14E-50 | 0.1963 | -14.8584 | -2.91623 | 42274.33 | poly(ADP-ribose) polymerase family member 14 |
| METTL7B | 1.46E-10 | 9.87E-12 | 0.4301 | -6.80839 | -2.92831 | 38.25 | methyltransferase like 7B |
| LOC112268296 | 8.37E-43 | 1.19E-45 | 0.2076 | -14.1818 | -2.94342 | 897.36 |  |
| LHFPL2 | 2.78E-36 | 6.98E-39 | 0.2261 | -13.0428 | -2.94929 | 2724.42 | LHFPL tetraspan subfamily member 2 |
| CXCL11 | 1.10E-07 | 1.33E-08 | 0.5201 | -5.68221 | -2.95514 | 15.25 | C-X-C motif chemokine ligand 11 |
| CAPNS2 | 7.36E-17 | 1.84E-18 | 0.3402 | -8.76691 | -2.98246 | 34.28 | calpain small subunit 2 |
| CDHR5 | 4.66E-17 | 1.13E-18 | 0.3412 | -8.82184 | -3.00992 | 24.9 | cadherin related family member 5 |
| CMPK2 | 7.69E-22 | 1.05E-23 | 0.3008 | -10.0368 | -3.01914 | 3097.57 | cytidine/uridine monophosphate kinase 2 |
| IFIT3 | 3.02E-27 | 2.13E-29 | 0.2701 | -11.2575 | -3.04015 | 32073.58 | interferon induced protein with tetratricopeptide repeats 3 |
| CASP5 | 1.25E-25 | 9.78E-28 | 0.2807 | -10.9149 | -3.06393 | 2340.34 | caspase 5 |
| EXOC3L1 | 3.95E-18 | 8.24E-20 | 0.3363 | -9.10998 | -3.06396 | 71.01 | exocyst complex component 3 like 1 |
| LOC105370355 | 9.84E-23 | 1.17E-24 | 0.2993 | -10.251 | -3.0681 | 78.66 | uncharacterized LOC105370355 |
| ZDHHC19 | 1.22E-23 | 1.24E-25 | 0.2936 | -10.4656 | -3.07282 | 158.43 | zinc finger DHHC-type palmitoyltransferase 19 |
| GBP7 | 1.07E-15 | 3.02E-17 | 0.3696 | -8.44565 | -3.12182 | 89.4 | guanylate binding protein 7 |
| MYOF | 5.73E-32 | 2.26E-34 | 0.2577 | -12.2261 | -3.15076 | 4592.22 | myoferlin |
| BCL2L14 | 6.30E-12 | 3.29E-13 | 0.4359 | -7.28203 | -3.17438 | 31.77 | BCL2 like 14 |
| SCARF1 | 8.55E-42 | 1.40E-44 | 0.2267 | -14.0076 | -3.17496 | 4184.1 | scavenger receptor class F member 1 |
| LYPD5 | 8.10E-22 | 1.11E-23 | 0.3181 | -10.0313 | -3.19122 | 35 | LY6/PLAUR domain containing 5 |
| FAM225B | 3.62E-30 | 1.74E-32 | 0.269 | -11.8678 | -3.19219 | 201.03 | family with sequence similarity 225 member B |
| HCAR1 | 1.93E-10 | 1.34E-11 | 0.4725 | -6.76432 | -3.1961 | 31.85 | hydroxycarboxylic acid receptor 1 |
| LOC105369735 | 1.83E-09 | 1.53E-10 | 0.5033 | -6.40186 | -3.22203 | 12.29 | uncharacterized LOC105369735 |
| EFCAB2 | 6.49E-31 | 3.01E-33 | 0.269 | -12.0137 | -3.23178 | 1141.61 | EF-hand calcium binding domain 2 |
| TNFAIP6 | 4.23E-28 | 2.75E-30 | 0.2827 | -11.4364 | -3.23276 | 5222.56 | TNF alpha induced protein 6 |
| IFI44L | 8.66E-13 | 3.89E-14 | 0.4275 | -7.56447 | -3.23383 | 4507.9 | interferon induced protein 44 like |
| FAM225A | 6.49E-29 | 3.72E-31 | 0.2805 | -11.6087 | -3.25626 | 281.71 | family with sequence similarity 225 member A |
| LOC101929319 | 1.89E-29 | 1.01E-31 | 0.2779 | -11.7198 | -3.25638 | 383.01 | uncharacterized LOC101929319 |
| CCL2 | 1.68E-13 | 6.57E-15 | 0.4181 | -7.79249 | -3.25771 | 64.43 | C-C motif chemokine ligand 2 |
| GSDMC | 1.57E-12 | 7.38E-14 | 0.4383 | -7.48092 | -3.27862 | 17.86 | gasdermin C |
| CEACAM1 | 2.09E-34 | 6.40E-37 | 0.261 | -12.6938 | -3.31258 | 6210.13 | CEA cell adhesion molecule 1 |
| C2 | 8.89E-29 | 5.15E-31 | 0.2874 | -11.5809 | -3.32783 | 878.16 | complement C2 |
| LINC01093 | 2.69E-08 | 2.81E-09 | 0.5641 | -5.9421 | -3.35189 | 80.86 | long intergenic non-protein coding RNA 1093 |
| SLAMF8 | 1.30E-29 | 6.74E-32 | 0.2854 | -11.754 | -3.35474 | 1460.04 | SLAM family member 8 |
| LOC105374296 | 1.07E-21 | 1.49E-23 | 0.3357 | -10.0024 | -3.358 | 67.37 | uncharacterized LOC105374296 |
| NRN1 | 7.24E-12 | 3.83E-13 | 0.4661 | -7.2613 | -3.38472 | 131.53 | neuritin 1 |
| SOCS1 | 2.05E-25 | 1.66E-27 | 0.3146 | -10.8668 | -3.41859 | 728.78 | suppressor of cytokine signaling 1 |
| NECTIN2 | 3.67E-12 | 1.86E-13 | 0.4679 | -7.35846 | -3.44289 | 1082.74 | nectin cell adhesion molecule 2 |
| LOC105371082 | 4.35E-30 | 2.12E-32 | 0.2915 | -11.8514 | -3.45489 | 260.07 | uncharacterized LOC105371082 |
| CFB | 2.06E-18 | 4.08E-20 | 0.3783 | -9.18589 | -3.47462 | 41.91 | complement factor B |
| VAMP5 | 1.45E-33 | 4.84E-36 | 0.2782 | -12.5344 | -3.48727 | 4746.33 | vesicle associated membrane protein 5 |
| GBP4 | 3.71E-53 | 1.01E-56 | 0.2197 | -15.8705 | -3.48744 | 21444.92 | guanylate binding protein 4 |
| RSAD2 | 1.47E-14 | 4.85E-16 | 0.4359 | -8.11522 | -3.53707 | 9517.51 | radical S-adenosyl methionine domain containing 2 |
| BNIP5 | 2.79E-16 | 7.41E-18 | 0.4162 | -8.60841 | -3.58302 | 30.5 | BCL2 interacting protein 5 |
| LINC02471 | 1.85E-17 | 4.24E-19 | 0.4037 | -8.93052 | -3.60497 | 39.49 | long intergenic non-protein coding RNA 2471 |
| LAP3 | 6.60E-42 | 1.05E-44 | 0.2571 | -14.0283 | -3.60636 | 18122.54 | leucine aminopeptidase 3 |
| LOC105369593 | 2.94E-09 | 2.55E-10 | 0.5704 | -6.32383 | -3.60687 | 23.77 | uncharacterized LOC105369593 |
| PANDAR | 1.19E-37 | 2.92E-40 | 0.272 | -13.2825 | -3.61257 | 252.47 | promoter of CDKN1A antisense DNA damage activated RNA |
| WARS1 | 2.43E-44 | 2.92E-47 | 0.2508 | -14.4395 | -3.62194 | 89275.6 | tryptophanyl-tRNA synthetase 1 |
| SEPTIN4-AS1 | 5.68E-23 | 6.52E-25 | 0.3583 | -10.3075 | -3.69314 | 70.89 | SEPTIN4 antisense RNA 1 |
| LRRK2-DT | 4.43E-17 | 1.07E-18 | 0.4192 | -8.82784 | -3.7007 | 67.91 | LRRK2 divergent transcript |
| LINC02555 | 3.71E-27 | 2.63E-29 | 0.3296 | -11.2387 | -3.70428 | 1023.86 | long intergenic non-protein coding RNA 2555 |
| IDO2 | 1.38E-22 | 1.68E-24 | 0.3662 | -10.2158 | -3.74153 | 69.93 | indoleamine 2,3-dioxygenase 2 |
| H2BC18 | 1.99E-41 | 3.47E-44 | 0.2725 | -13.943 | -3.79945 | 1200.99 | H2B clustered histone 18 |
| LOC100996318 | 1.51E-35 | 4.04E-38 | 0.2966 | -12.9083 | -3.82894 | 770.33 | uncharacterized LOC100996318 |
| SMTNL1 | 7.35E-22 | 1.00E-23 | 0.3841 | -10.0417 | -3.8571 | 673.85 | smoothelin like 1 |
| CALHM6 | 2.20E-41 | 3.96E-44 | 0.2795 | -13.9336 | -3.89501 | 3691.8 | calcium homeostasis modulator family member 6 |
| LOC112268237 | 7.61E-32 | 3.12E-34 | 0.3207 | -12.1997 | -3.91243 | 192.79 |  |
| LOC105373645 | 9.33E-22 | 1.29E-23 | 0.395 | -10.0166 | -3.95701 | 21.69 | uncharacterized LOC105373645 |
| CXCL9 | 4.19E-22 | 5.44E-24 | 0.3933 | -10.1014 | -3.97292 | 159.39 | C-X-C motif chemokine ligand 9 |
| FCGR1BP | 3.27E-47 | 2.15E-50 | 0.2721 | -14.9286 | -4.06186 | 10112.66 | Fc gamma receptor Ib, pseudogene |
| H2BP2 | 3.25E-42 | 4.98E-45 | 0.2898 | -14.0809 | -4.08053 | 716.18 | H2B histone pseudogene 2 |
| GBP1 | 1.68E-56 | 2.76E-60 | 0.252 | -16.3777 | -4.12748 | 43911.17 | guanylate binding protein 1 |
| GBP5 | 5.24E-51 | 2.29E-54 | 0.2661 | -15.5267 | -4.13089 | 98659.83 | guanylate binding protein 5 |
| P2RY14 | 1.03E-32 | 3.61E-35 | 0.3341 | -12.3741 | -4.13469 | 5903.67 | purinergic receptor P2Y14 |
| SEPTIN4 | 3.30E-34 | 1.03E-36 | 0.3297 | -12.6567 | -4.17248 | 888.41 | septin 4 |
| IL31RA | 8.38E-35 | 2.43E-37 | 0.3335 | -12.7695 | -4.25868 | 403.32 | interleukin 31 receptor A |
| ATF3 | 6.66E-47 | 4.73E-50 | 0.2917 | -14.8759 | -4.33861 | 893.54 | activating transcription factor 3 |
| CD274 | 2.43E-44 | 2.90E-47 | 0.3047 | -14.4401 | -4.39944 | 6334.84 | CD274 molecule |
| FCGR1CP | 4.57E-48 | 2.50E-51 | 0.2922 | -15.0714 | -4.40415 | 7012.5 | Fc gamma receptor Ic, pseudogene |
| PDCD1LG2 | 6.64E-36 | 1.74E-38 | 0.3397 | -12.9729 | -4.40635 | 483.88 | programmed cell death 1 ligand 2 |
| FCGR1A | 1.93E-45 | 1.80E-48 | 0.304 | -14.6305 | -4.44746 | 18382.24 | Fc gamma receptor Ia |
| GBP6 | 4.18E-28 | 2.70E-30 | 0.3965 | -11.4382 | -4.53512 | 833.05 | guanylate binding protein family member 6 |
| LAMP3 | 2.58E-45 | 2.54E-48 | 0.3137 | -14.6069 | -4.58148 | 1424.58 | lysosomal associated membrane protein 3 |
| LOC105373582 | 2.17E-52 | 7.12E-56 | 0.2964 | -15.7477 | -4.66741 | 414.15 | uncharacterized LOC105373582 |
| ACOD1 | 8.32E-24 | 8.36E-26 | 0.4516 | -10.503 | -4.74336 | 53.56 | aconitate decarboxylase 1 |
| IDO1 | 1.38E-22 | 1.70E-24 | 0.4755 | -10.215 | -4.85722 | 11220.82 | indoleamine 2,3-dioxygenase 1 |
| SERPING1 | 1.22E-51 | 4.68E-55 | 0.3186 | -15.6281 | -4.97964 | 17364.47 | serpin family G member 1 |
| BATF2 | 1.48E-48 | 7.26E-52 | 0.3376 | -15.1528 | -5.11582 | 5500.53 | basic leucine zipper ATF-like transcription factor 2 |
| ETV7 | 2.31E-38 | 5.42E-41 | 0.3844 | -13.4081 | -5.15429 | 3271.3 | ETS variant transcription factor 7 |
| LINC02528 | 3.96E-27 | 2.83E-29 | 0.4697 | -11.2323 | -5.27603 | 62.76 | long intergenic non-protein coding RNA 2528 |
| CXCL10 | 3.84E-32 | 1.45E-34 | 0.4381 | -12.2621 | -5.37243 | 978.13 | C-X-C motif chemokine ligand 10 |
| GBP1P1 | 4.77E-60 | 5.21E-64 | 0.3329 | -16.8914 | -5.62317 | 1053.98 | guanylate binding protein 1 pseudogene 1 |
| APOL4 | 6.90E-44 | 9.04E-47 | 0.4153 | -14.3614 | -5.96454 | 1204.48 | apolipoprotein L4 |
| LIPM | 3.33E-44 | 4.19E-47 | 0.4275 | -14.4146 | -6.16283 | 801.55 | lipase family member M |
| ANKRD22 | 1.63E-47 | 9.78E-51 | 0.4136 | -14.981 | -6.1967 | 5254.01 | ankyrin repeat domain 22 |
